# Aberrant STAT signaling drives T cell dysregulation in a targetable pediatric sepsis endotype

**DOI:** 10.1101/2024.06.11.24308709

**Authors:** Robert B. Lindell, Samir Sayed, Jose S. Campos, Sydney A. Sheetz, Apoorva Babu, Montana S. Knight, Andrea A. Mauracher, Ceire A. Hay, Peyton E. Conrey, Julie C. Fitzgerald, Nadir Yehya, Stephen T. Famularo, Teresa Arroyo, Richard Tustin, Hossein Fazelinia, Edward M. Behrens, David T. Teachey, Alexandra F. Freeman, Jenna R. E. Bergerson, Steven M. Holland, Jennifer W. Leiding, Scott L. Weiss, Mark W. Hall, Deanne M. Taylor, Rui Feng, E. John Wherry, Nuala J. Meyer, Sarah E. Henrickson

## Abstract

Sepsis is a leading cause of morbidity and mortality in critically ill children, yet heterogeneity in immune responses complicates the development of targeted therapies. Although immune dysregulation is associated with poor outcomes in sepsis, it remains unclear which host immune factors contribute causally to sepsis morbidity and mortality. To address this gap, we integrated deep immune phenotyping, plasma proteomics, single-cell transcriptomics, and phosphoflow cytometry in a prospective cohort of 88 critically ill children to elucidate the immunologic mechanisms which underly disease heterogeneity. Unsupervised clustering of plasma cytokines identified three immunologic subgroups, including a high-severity group (“Group C”) characterized by marked hypercytokinemia, driven primarily by IL-6 and IFN-γ. Group C exhibited distinct alterations in immune cell frequency and activation status, along with a strong association between hyperinflammatory signaling and lymphocyte dysfunction. Single-cell RNA sequencing revealed transcriptional signatures of T cell activation and metabolic stress, and identified widespread suppression of a lymphoid protective gene program across CD8⁺ T cell subsets. In the setting of increased expression of activation markers, T cell receptor repertoire analysis revealed no dominant clonotypes, consistent with a bystander mechanism of T cell activation. Using phosphoflow cytometry, we demonstrated baseline hyperactivation of STAT1 and STAT3 in CD8⁺ T cells from patients in Group C, and these cells failed to respond to aCD3/aCD28 stimulation. Together, these findings define IL-6/IFN-γ–driven T-cell dysfunction as a distinct endotype of immune dysregulation in pediatric sepsis, highlighting the JAK/STAT axis as a potential future target for immunomodulatory therapy.

## INTRODUCTION

Sepsis is defined as life-threatening organ dysfunction that develops in the setting of a dysregulated immune response to infection (*1*) and represents a leading cause of both adult and pediatric mortality worldwide (*2*). Pediatric sepsis is associated with distinct epidemiology and outcomes compared to adult sepsis (*3*), with the majority of pediatric sepsis deaths occurring in immunocompromised children (*4–6*) and in the setting of persistent multiple organ dysfunction syndrome (MODS) (*7, 8*). MODS represents a “final common pathway” by which severe, systemic inflammation arising from diverse clinical insults (e.g. sepsis, trauma, shock) leads to progressive organ failure due to a combination of endothelial, epithelial, mitochondrial, and immunologic dysfunction (*9–11*); in children, sepsis is the most common cause of MODS (*12–14*).

Identifying modifiable molecular or physiological mechanisms that drive disease is an essential first step for translating fundamental biologic insights into precision therapeutics (*15*). In pediatric sepsis, both innate and adaptive immune dysfunction (*16–18*) are associated with secondary infection (*19*), persistent organ dysfunction (*20*) and mortality (*21*). Mitochondrial dysfunction, a hallmark of this sepsis-associated immune suppression (*22, 23*), is also associated with organ failure in pediatric sepsis (*24–26*). Yet despite the link between immune dysregulation and both morbidity and mortality in pediatric sepsis, successful immune modulation in sepsis has been elusive (*27*). Multiple trials of targeted immunomodulation (e.g. IL-1, TNF) have failed to improve survival in adults (*28–31*), and it remains unclear which molecular mechanisms contribute causally to adverse sepsis outcomes (*32*).

A successful precision medicine approach to pediatric sepsis will require an understanding of the molecular events which underlie the development of organ failure, a knowledge of which events are reversible, and an ability to identify high-risk patients in real time (*33*). Existing approaches to identify pediatric sepsis endotypes based on clinical and laboratory features have identified several high-risk patient subgroups (*34–37*), but these subgroups lack defined mechanisms of immune dysregulation and thus cannot inform targeted treatment decisions. Variants in a wide range of genes associated with inborn errors of immunity (IEI) have been identified in a minority of pediatric patients with sepsis (*38–40*), suggesting that diverse genetic factors may also contribute to the dysregulated host immune phenotypes that develop in the setting of critical illness. Recent attempts to define this heterogeneous host response in adults with sepsis using molecular approaches (*41–47*) have yielded new insights into sepsis pathobiology, suggesting that deep immune phenotyping may be able to clarify the heterogeneity within key immunologic pathways that has previously hampered precision sepsis efforts. In support of this concept, recent reanalysis of the negative PROWESS-SHOCK trial (*31*) demonstrated varied response to treatment with activated protein C across biomarker-defined sepsis subgroups, with benefit in hyperinflammatory patients and harm in hypoinflammatory patients (*48*). The lower burden of chronic comorbidities in children affords a clearer biologic signal, positioning pediatric sepsis as an optimal cohort for discovering and targeting treatable endotypes in sepsis.

Given the role of immune dysregulation in sepsis pathobiology and the lack of novel targeted therapies for these high-risk patients, we designed a longitudinal multi-omics cohort study of pediatric MODS patients with and without sepsis with the goal of identifying targetable endotypes. We hypothesized that severity-associated subgroups would be associated with targetable mechanisms of immune dysregulation that could improve outcomes. Using longitudinal proteomics, cytometry, and transcriptional analysis, we defined three prognostic subgroups in critically ill children with and without sepsis and identified an association between dysregulated STAT1 and STAT3 signaling and CD8^+^ T cell hyperactivation in the subset of pediatric MODS patients with both the highest severity of illness and worst prognosis.

## RESULTS

### Proteomic heterogeneity of pediatric critical illness

To identify endotypes in patients with and without sepsis, we collected peripheral whole blood samples in a prospective, observational cohort of 88 pediatric patients with MODS. Blood was collected within 48 hours of MODS onset and then twice weekly through death or resolution of MODS. At each timepoint, peripheral blood mononuclear cells (PBMC) and heparinized plasma biospecimens were cryopreserved for later analysis, as shown in **Fig. 1A**. We compared these patients to a separate cohort of 25 pediatric healthy control (HC) participants. Our initial analyses included a 1536-marker proteomics panel from Olink Proteomics selected for patient stratification and identification of candidate therapeutic targets (*49*) (**Table S1**) and a 35-marker spectral flow cytometry-based immune phenotyping panel optimized to define immune cell subset abundance and activation state (**Table S2**).

**Fig 1.**
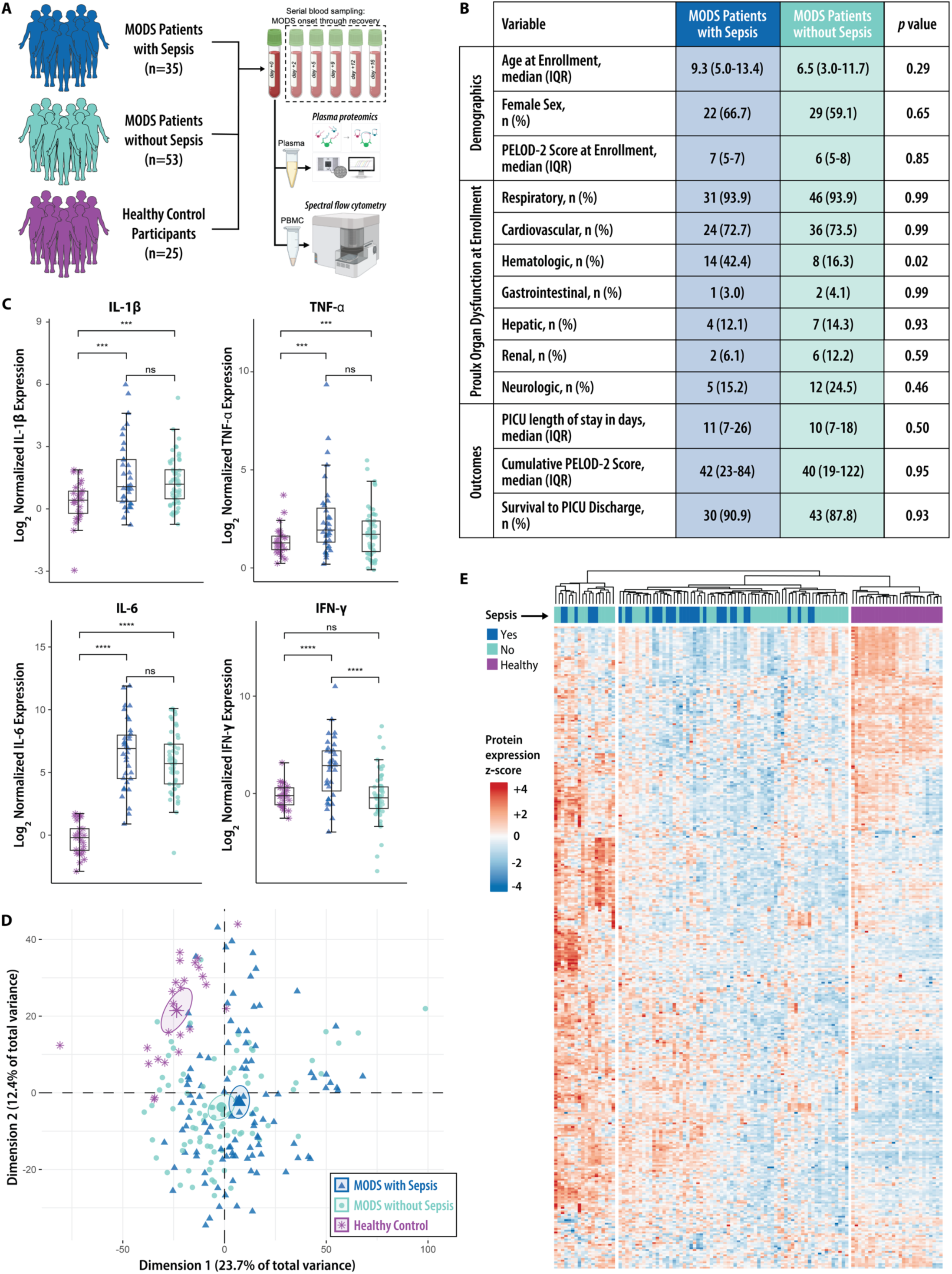
Shared proteomic features in pediatric MODS patients with and without sepsis. **(A)** Schematic of participant cohorts and sample processing pipeline using plasma proteomics and spectral flow cytometry. **(B)** Cohort demographics, exposures, and clinical outcomes, stratified by MODS etiology (i.e. sepsis vs non-sepsis). Comparisons between groups using chi-squared and Wilcoxon rank sum test, as appropriate. **(C)** Proinflammatory cytokines are increased in patients with MODS but do not differ between patients with and without sepsis, except for IFN-γ which is increased in patients with sepsis compared to patients without sepsis. Comparisons were made between groups as shown and determined by Wilcoxon rank sum test. Box-and-whiskers plots include a box indicating median and interquartile range, and whiskers extending to the data point not further than the 1.5x interquartile range. **(D)** Principal component analysis based on expression of 1,448 proteins analyzed by proximity extension assay in 186 samples from 88 patients with MODS and 25 pediatric healthy control (HC) participants reveals substantial heterogeneity and overlap between MODS patients with and without sepsis. **(E)** Clustered heatmap showing row-normalized protein expression among 88 patients at MODS onset and 25 HC participants. Colored annotation bar indicates MODS subgroup.

Demographics and clinical outcomes were similar between our 35 MODS patients with sepsis and 53 MODS patients without sepsis (**Fig. 1B**), and the only organ dysfunction category that differed between groups was hematologic dysfunction, which was driven by increased thrombocytopenia among patients with sepsis at MODS onset (platelet count median [IQR]: 119 [99-187] vs 199 [131-278], *p*<0.001; normal range 150-450 x10^3^/μl). Primary and secondary patient outcomes did not differ between MODS subgroups, including cumulative PELOD-2 (Pediatric Logistic Organ Dysfunction-2) (*13*) organ dysfunction score, survival to PICU (pediatric intensive care unit) discharge, and duration of PICU stay.

Sepsis is characterized by a surge in pro-inflammatory cytokines, and we hypothesized that expression of these cytokines would differ between patients with and without sepsis. While we noted the expected increased proinflammatory cytokines (interleukin (IL)-1β, IL-6, tumor necrosis factor (TNF), IL-18, monocyte chemoattractant protein (MCP)-1, interferon (IFN)-γ, all p<0.0001) in patients with sepsis compared to HC participants, many of these cytokines were also markedly elevated in MODS patients without sepsis. We found that median IL-1β, TNF-α, and IL-6 levels were elevated in MODS patients and did not differ between patients with and without sepsis, while IFN-γ was higher in patients with sepsis (*p*<0.0001) (**Fig. 1C**). We then used principal component analysis (PCA) to visualize the proteomic heterogeneity within the MODS cohort and overlap between patients with and without sepsis. We noted separation between HC participants and MODS patients but substantial overlap between MODS patients with and without sepsis (**Fig. 1D**). Patients with MODS also exhibited substantial heterogeneity along principal component 1 and principal component 2. These dimensions were shaped by proteins involved in biologic processes such as growth regulation (e.g. MAD1L1, HDGF), inflammatory signaling (e.g. CASP3, MAP3K5), and endothelial dysfunction (e.g. PLAUR, SRC), as illustrated by the top PCA loadings (**Table S3**).

To better understand the shared proteomic landscape of MODS patients with and without sepsis, we constructed a heatmap of row-normalized protein expression with the full proteomics dataset and used hierarchical clustering to visualize heterogeneity within the cohort at MODS onset. As shown in **Fig. 1E**, hierarchical clustering separates HC participants from patients with MODS but does not discriminate between MODS patients with sepsis and without sepsis, findings which are complementary to the PCA. This biologic overlap suggests that MODS develops in the setting of overwhelming proinflammatory signals and may represent a final common pathway of critical illness, independent of the inciting diagnosis and presence of infection.

### Identification of three severity-associated subgroups

To identify targetable immune dysregulation in children with MODS, we first sought to identify severity-associated subgroups based on plasma protein expression. To begin, we used linear mixed-effects models to identify plasma proteins associated with severity of illness, defined by the PELOD-2 organ dysfunction score, after adjustment for age and sex as fixed effects and day from MODS onset as a random effect. This yielded a set of 214 plasma proteins which were significantly associated with illness severity (**Table S4**). We then employed consensus clustering (*50*) to identify distinct subgroups of patients based on protein expression (**Fig. 2A**). Optimal cluster number (*k*=3) was identified via Monte Carlo bootstrapping and confirmed by elbow method and gap statistic (**Fig. S1**). To understand association between subgroups and clinical outcomes, we tested the effect of subgroup membership on the cumulative incidence of both mortality and survival to PICU discharge with the Fine-Gray subdistribution hazard model (*51*). In this survival analysis, patients in Group C have higher cumulative incidence of death (*p*=0.03) and lower cumulative incidence of survival to PICU discharge (*p*=0.04) compared to patients in Groups A and B, with separation occurring in the first week and persisting to day +28 (**Fig. 2B**).

**Fig. 2.**
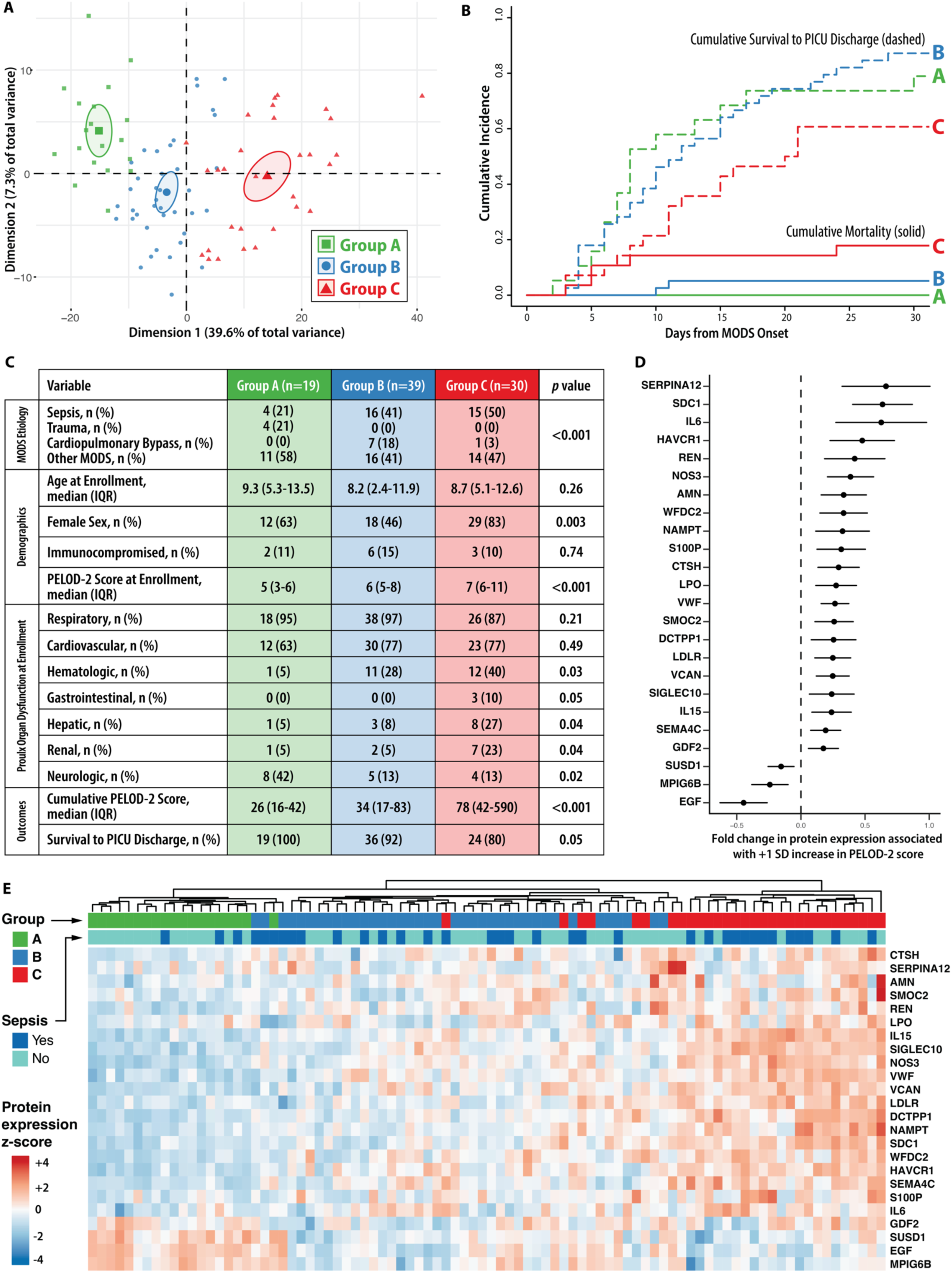
Three subgroups based on protein expression at MODS onset. **(A)** Principal component analysis based on expression of 214 severity-associated proteins in 88 patients at MODS onset reveals separation among three subgroups defined through Monte Carlo consensus clustering (*50*). **(B)** Cumulative incidence of survival to PICU discharge (dashed lines) and PICU mortality (solid lines) by MODS day among three subgroups. **(C)** MODS etiology, cohort demographics, exposures, and clinical outcomes, stratified by subgroup. Comparisons between groups by chi-squared and Wilcoxon rank sum test, as appropriate. **(D)** Forest plot showing estimated fold-change in normalized protein expression associated with a +1 standard deviation increase in PELOD-2 score for 24 proteins identified through ordinal elastic net regression (*53*) which differ across subgroups. Forest plot includes a point estimate of fold-change and whiskers which represent the 95% confidence interval around this estimate. **(E)** Heatmap showing normalized protein expression of the 24 proteins identified by ordinal elastic net regression among 88 patients at MODS onset. Colored annotation bars indicate subgroup and sepsis status.

Though defined by protein expression alone, these identified subgroups also differ by clinical features and outcomes (**Fig. 2C**). MODS etiology varies by Group, with similar proportions of sepsis patients in Group B and C and all trauma patients in Group A. Group C patients have higher severity of illness, more non-cardiopulmonary organ failures, and increased cumulative organ dysfunction scores and mortality compared to Groups A/B. Etiology of MODS and computed subgroup for each patient is detailed in **Table S5**. Immunocompromised status, defined as active malignancy, hematopoietic cell transplantation, or primary immunodeficiency, was identified in 13% of MODS patients (11/88), and patients with immunocompromised status were not imbalanced across subgroups (p=0.74), as detailed in **Table S6**.

Noting an ordinal increase in number of organ failures and mortality across protein-derived subgroups, we hypothesized a reduced set of plasma proteins could successfully identify these subgroups and could thus be more suitable for translation to the clinical setting in order to identify and test potential underlying mechanisms of immune dysregulation. Elastic net regularization is a common approach to generate a high-performing sparse model with good predictive accuracy (*52*). To define a parsimonious protein signature, we trained an ordinal elastic net model (*53*) and generated a 24-protein signature which successfully discriminates the three subgroups (**Fig. S2**).

We next sought to evaluate the performance of this parsimonious protein model through two complementary approaches. First, we tested the association between the elastic net proteins and severity of illness. Using a linear mixed-effects model, we estimated the fold change in protein expression associated with one standard deviation change in PELOD-2 organ dysfunction score (**Fig. 2D**). This model demonstrates that modest changes in expression of these 24 proteins are associated with meaningful differences in illness severity. Second, we tested the discrimination of this 24-protein signature. As shown in the heatmap of protein expression at MODS onset in **Fig. 2E**, hierarchical clustering of elastic net proteins effectively separates subgroups but does not discriminate between patients with and without sepsis.

To quantify the discrimination of subgroups using the parsimonious elastic net protein set, we employed linear discriminant analysis to calculate the polytomous discrimination index (PDI), a measure of rank-based discrimination performance, for each subgroup. Category-specific PDI indicated excellent discrimination for each subgroup (Group A 0.98, Group B 0.96, Group C 0.99), and overall PDI for the model was 0.98. Taken together, these results suggest that our reduced 24-protein signature reflects severity of illness and successfully discriminates subgroups at MODS onset.

### Immune cell frequency and activation vary by subgroup

Having identified three severity-associated subgroups (Groups A, B, and C) from protein expression data and built a parsimonious classification model, we next sought to define underlying mechanisms of immune dysregulation. We hypothesized that protein-derived subgroups would be associated with differences in cellular immunophenotype, and that these differences would help clarify the underlying mechanisms of immune dysregulation. For this analysis, we performed high dimensional spectral flow cytometry on cryopreserved PBMCs obtained at MODS onset using a custom-designed 35 marker panel (**Table S2**) which includes both phenotypic and functional markers and compared to HC participants. After arcsinh scaling (*54*) and quality control with flowAI (*55*), we performed FlowSOM metaclustering (*56*) and identified 14 immune cell populations by surface and intracellular marker expression. **Fig. S3** presents a representative example of our manual gating strategy for this immune phenotyping panel, which we used to confirm the identity of the FlowSOM metaclusters.

To measure the immunophenotypic differences between HC participants and patients in the three subgroups (**Fig. 3A**), we subsampled the data to 100,000 cells per subgroup and applied t-distributed Stochastic Neighbor Embedding with Compute Unified Device Architecture (tSNE-CUDA) dimensionality reduction (*57*). Differences in cell populations by subgroup are quantified in stacked bar plots (**Fig. 3B**). The proportional abundance of central memory, effector memory, and Temra CD8^+^ T cells were markedly reduced in Group C patients (**Fig. 3C**), and this proportional loss of non-naïve CD8^+^ T cells was strongly associated with severity of illness by linear regression (*p*<0.001). A similar ordinal trajectory was identified across multiple cell types (**Fig. 3D**), including T cells, B cells, NK cells, and dendritic cells (Cuzik test of trend *p*<0.001 for each cell type shown), and reduction in frequency of each of these cell types was strongly associated with severity of illness by linear regression (all *p*<0.001).

**Fig. 3.**
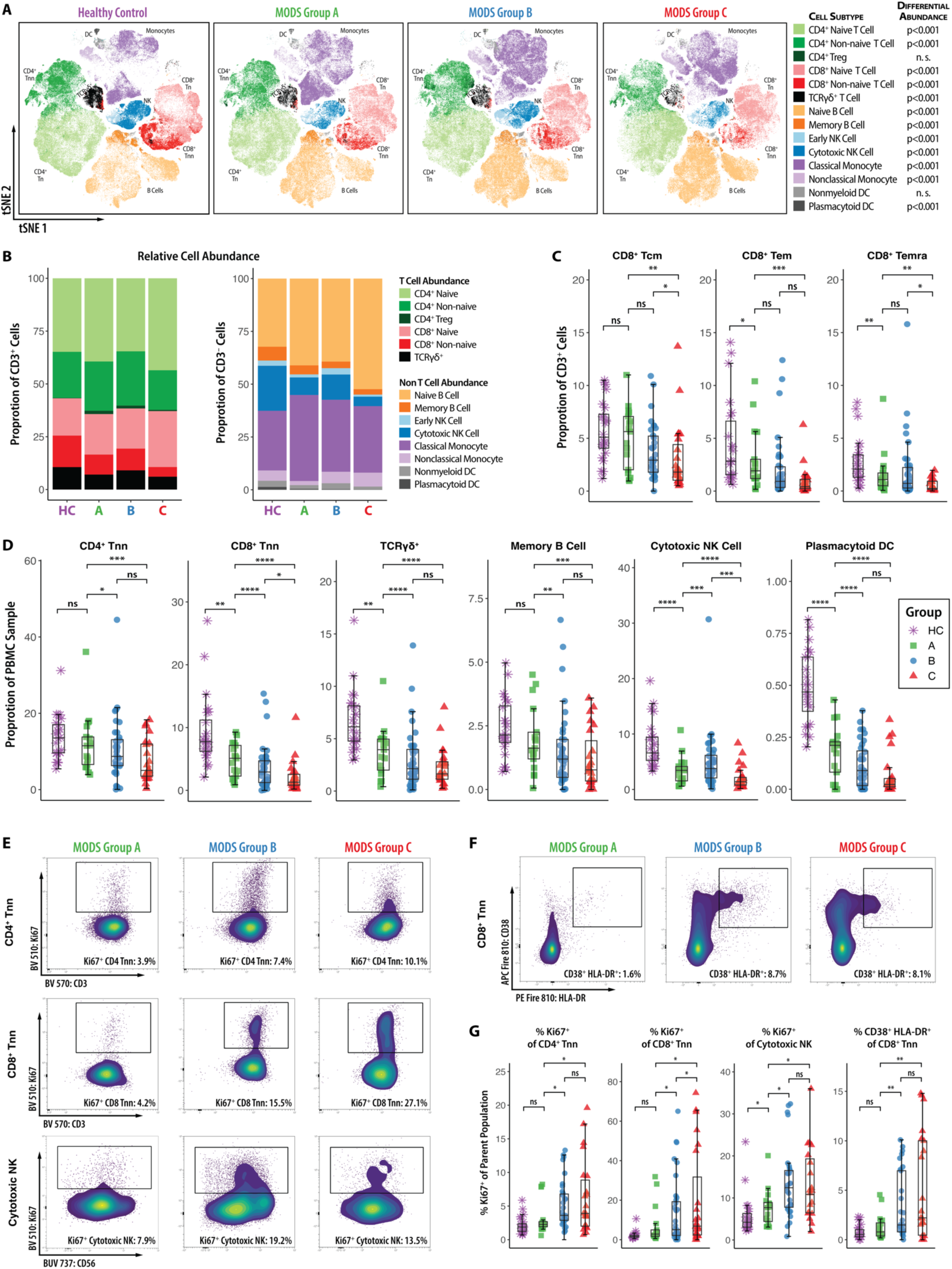
Immune cell abundance and activation state vary by plasma protein-derived subgroup. **(A)** PBMC immune phenotype tSNE-CUDA projection demonstrates marked differences in frequency of lymphoid and myeloid lineages between HC participants and patients with MODS, and among subgroups. Differential abundance across groups by cell subtype was determined by Kruskal-Wallis test. **(B)** Stacked bar plots show differences in CD3^+^ and CD3^-^ cell proportional abundance by subgroup corresponding to the FlowSOM metaclusters in the tSNE-CUDA projections at left. **(C)** Compared to HC participants, the proportional abundance of CD8^+^ T cell subsets is reduced in patients with MODS, with an ordinal trajectory across subgroups noted in CD8^+^ central memory cells, CD8^+^ effector memory cells, and CD8^+^ Temra cells. Effector CD8^+^ cells in Group C patients are significantly reduced in frequency compared to other subgroups. Comparisons were made between groups as shown and determined by Wilcoxon rank sum test. **(D)** Shifts in other lymphoid and myeloid subsets across subgroups demonstrate a proportional reduction of peripheral non-naïve CD4^+^ and CD8^+^ T cells, TCRγδ^+^ T cells, memory B cells, cytotoxic NK cells, and plasmacytoid dendritic cells, with the most marked differences in the Group C patients. Comparisons were made between groups as shown and determined by Wilcoxon rank sum test. **(E)** In non-naïve CD4^+^, non-naïve CD8^+^, and cytotoxic NK cells, Ki67 expression is increased Group B and Group C patients compared to Group A (all *p*<0.05), indicative of increased proliferation of these effector cell subsets. **(F)** In non-naïve CD8^+^ T cells, CD38 and HLA-DR co-expression is markedly increased Group B and Group C patients compared to Group A (both *p*<0.01), indicative of CD8^+^ T cell activation in these groups. **(G)** Boxplots quantifying differences in proliferation and activation by subgroup. Comparisons were made between groups as shown and determined by Wilcoxon rank sum test.

In addition to shifts in immune cell subset frequency, we hypothesized that protein-derived subgroups would be associated with differences in immune cell activation, as measured by expression of markers of proliferation (i.e. Ki67) and activation (i.e. CD38 and HLA-DR), across subgroups. Representative bivariate plots of Ki67 expression in non-naïve CD4^+^ and CD8^+^ T cells and cytotoxic NK cells (CD56^dim^ CD16^+^) demonstrate increased proliferation in Group B and Group C patients compared to Group A patients (**Fig 3E**). Similarly, CD38 and HLA-DR co-expression in non-naïve CD8^+^ T cells was markedly increased in Group B and Group C patients compared to Group A, but not different between Group B and Group C, indicative of CD8^+^ T cell activation in Groups B and C (**Fig. 3F**). Corresponding boxplots in **Fig. 3G** quantify these differences in proliferation and activation by subgroup. Taken together, these data suggest that CD8^+^ T cells have markedly reduced proportional abundance but concurrently have increased markers of proliferation and activation in Group C patients, and that these cellular phenotypes are associated with worsening severity of illness.

### Multiple concurrent mechanisms of immune dysregulation define Group C patients

Because Group C patients demonstrate immune dysregulation defined by distinct clinical, proteomic, and cellular features compared to other patients with MODS, we hypothesized that expression of key canonical inflammatory pathways would differ by subgroup. Using the proteomics dataset, we first examined differential protein expression after adjustment for patient age, sex, severity of illness, and days since MODS onset using a linear mixed-effects model. In unadjusted analysis, 1061/1448 measured proteins were differentially expressed in Group C (**Fig. 4A**), and in our adjusted model 1003/1061 proteins remained differentially expressed. For 98% (980/1003) of these differentially expressed proteins, expression was upregulated in Group C compared to Group A/B.

**Fig. 4.**
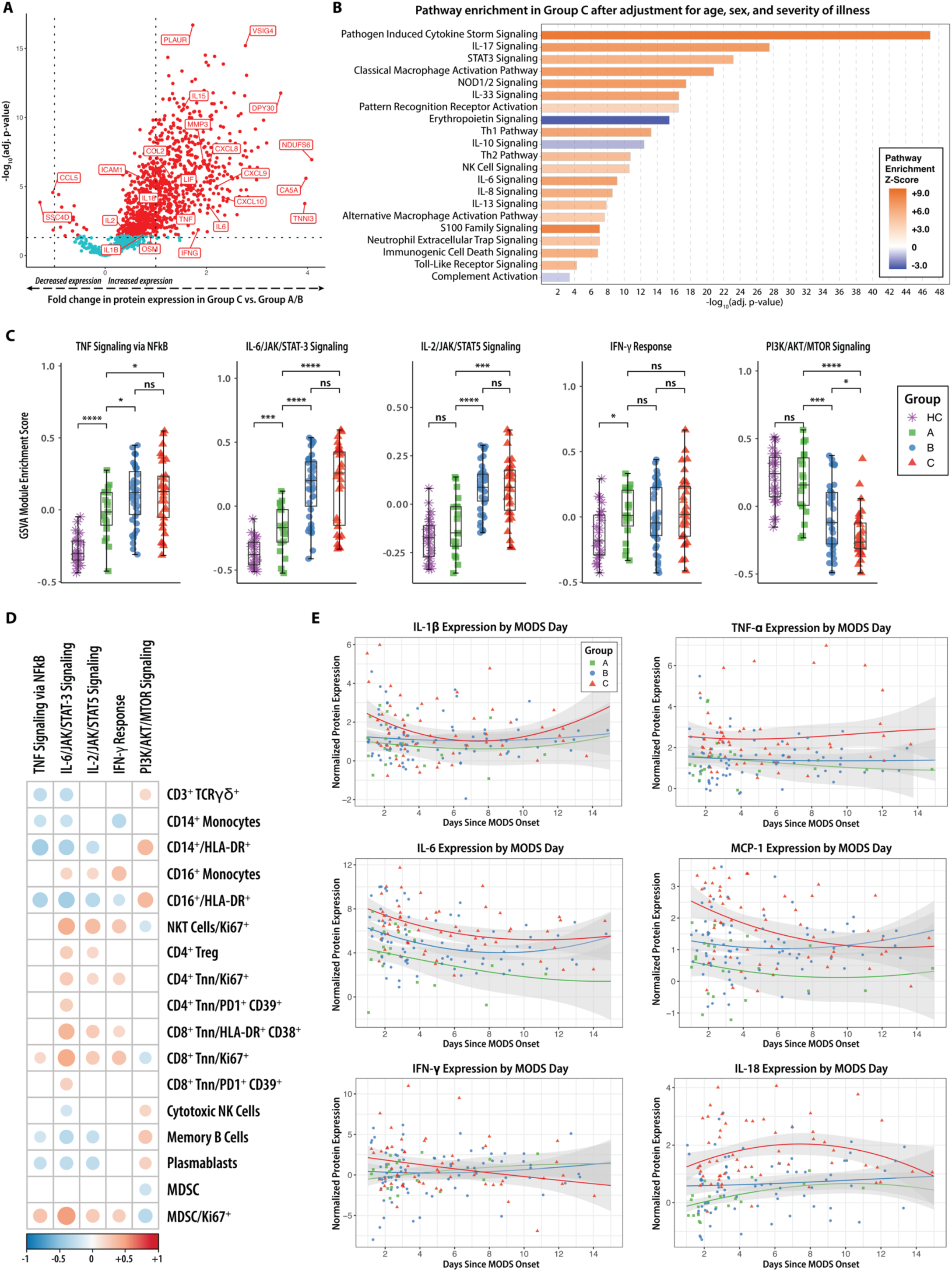
Group C patients have a hyperinflammatory plasma protein phenotype characterized by upregulation of multiple concurrent mechanisms of immune dysregulation. **(A)** Volcano plot comparing proteomic profile of Group C patients to Group A/B patients, showing that 1061/1448 measured proteins were differentially expressed in Group C. **(B)** After correction for patient age, sex, severity of illness and days since MODS onset with a linear mixed-effects model, 1003 proteins remained differentially expressed in group C. Based on these differentially expressed proteins, Ingenuity Pathway Analysis was used to identify enrichment of 22 canonical pathways in the Group C proteome after Benjamini-Hochberg correction. **(C)** Individual patient module enrichment scores calculated using GSVA and MSigDB Hallmark pathways demonstrate markedly increased IL-6/JAK/STAT3 and IL-2/JAK/STAT5 signaling in Group B and Group C patients, modest increase in TNF/NFκB signaling in Group B and Group C patients, and an ordinal reduction in PI3K/AKT/mTOR signaling across subgroups. Comparisons were made between groups as shown and determined by Wilcoxon rank sum test. **(D)** Spearman’s correlation between GSVA module enrichment score and immune cell proportional abundance, activation, and proliferation in MODS patients. Only correlations with Benjamini-Hochberg corrected *p*-value <0.05 are shown. **(E)** Longitudinal expression of canonical cytokine storm markers demonstrates persistence of immune dysregulation beyond MODS onset. For each panel, normalized protein expression is presented by days from MODS onset through day 14. Loess regression lines with 95% confidence intervals are presented for each subgroup.

To compare expression of key canonical inflammatory pathways across subgroups, we performed pathway enrichment analysis of our adjusted plasma protein expression dataset using complementary group- and patient-based strategies. First, we used Ingenuity Pathway Analysis (IPA, Qiagen) (*58*) to identify enrichment of 22 canonical pathways in the Group C proteome in comparison to Group A/B (**Fig. 4B**). Noting that 8 of the top 10 differentially expressed pathways identified by IPA were related to hyperinflammatory signaling, we then applied Gene Set Variation Analysis (GSVA) (*59*) to study enrichment of five canonical proinflammatory pathways on the individual patient level using Human Molecular Signatures Database (MSigDB) Hallmark gene sets (*60*). Using GSVA, we assigned patient-level protein module enrichment scores for each pathway of interest based on expression of proteins in the corresponding Hallmark gene set, as detailed in Methods. Module enrichment scores for each pathway are visualized in **Fig. 4C** and demonstrate markedly increased IL-6/JAK/STAT3 and IL-2/JAK/STAT5 module enrichment in Group B and Group C patients (each *p*<0.001 vs Group A), modest increase in TNF/NFκB signaling in Group B and Group C patients (each *p*<0.05 vs Group A), and an ordinal reduction in PI3K/AKT/mTOR signaling across subgroups (Cuzik test of trend *p*<0.001). IFN-γ response module enrichment scores did not vary by subgroup. We then assessed the correlation between Hallmark module enrichment and immune cell subset proportional abundance and activation in patients with MODS (**Fig. 4D**). We noted that IL-6/JAK/STAT3 module enrichment had the strongest correlation with proliferation and activation of immune cell subsets previously identified in **Fig. 3E-F**, including non-naïve CD4^+^ and CD8^+^ proliferation (Ki67 expression) and activation (HLA-DR/CD38 coexpression and PD-1/CD39 coexpression). Classical and non-classical monocyte HLA-DR expression was inversely correlated with IL-6/JAK/STAT3 module enrichment score, while immune regulatory cell populations (regulatory T cells, myeloid-derived suppressor cells) were positively correlated with IL-6/JAK/STAT3 module enrichment score.

Finally, we studied the longitudinal expression of cytokine storm markers (*61*) in patients with MODS to understand the duration of cytokinemia in Group C patients. Normalized protein expression by day since MODS onset by subgroup is shown in **Fig. 4E**. We noted that IL-6, MCP-1, and IL-18 expression were significantly higher in Group C patients for the first 7 days after MODS onset (each *p*<0.001 at day +7), while IL-1β and TNF-α expression remained different for only the first 4 days (*p*=0.05 and *p*=0.009 respectively at day +4) and IFN-γ expression did not differentiate subgroups at any timepoint.

### Immune dysregulation in MODS compared to patients with inborn errors of immunity that impact STAT1 and STAT3 signaling

The enrichment of STAT pathway signaling in Group C patients and its association with increased T cell activation by flow cytometry prompted us to ask whether this endotype reflects a physiologic adaptation to critical illness or a maladaptive, pathologic form of immune dysregulation. STAT3 is a central mediator of inflammatory signaling, implicated in endothelial dysfunction, capillary leak, emergency granulopoiesis, and disrupted lymphocyte homeostasis (*62*). Recent translational studies have highlighted its role in human sepsis, including the identification of immature CD66b⁺ neutrophils as markers of STAT3-driven emergency myelopoiesis in critically ill adults (*63*), and two recent preclinical studies have demonstrated a protective effect of selective STAT3 inhibition in mice with CLP-induced sepsis (*64, 65*). Informed by these studies, we hypothesized that the magnitude of STAT3 pathway activation in Group C patients is not merely an epiphenomenon but instead reflects pathogenic immune program that may constitute a viable target for precision immunomodulation.

To test this hypothesis, we compared plasma proteomic profiles between patients with MODS and participants with rare, monogenic inborn errors of immunity (IEI) involving constitutive activation or inactivation of the STAT3 and STAT1 signaling pathways. In this analysis, IEI offer a unique lens through which to interpret complex immune phenotypes, serving as arbiters that reveal the immunologic consequences of discrete signaling perturbations *in vivo*. Patients with four different IEI were included in this analysis: STAT1 gain-of-function (GOF) (n=9), STAT3 GOF (n=5), STAT1 autosomal dominant and dominant-negative (DN) (n=1), or STAT3 DN (n=3). We measured protein expression using a 384-marker inflammatory proteomics panel from Olink Proteomics (**Table S7**), and we analyzed pathway expression in bulk by Gene Set Enrichment Analysis (GSEA) (*66*) and at the individual patient level using GSVA (*59*).

A summary of our experimental design is shown in **Fig. 5A**, which includes 88 patients with MODS, 18 patients with IEI, and 25 HC participants. Orthogonal to our Ingenuity Pathway Analysis findings, we first confirmed population-level proteomic enrichment (normalized enrichment score 1.53, *p*-value 0.004) of the Hallmark IL-6/JAK/STAT3 signaling pathway in Group C patients at MODS onset compared to HC participants using GSEA (**Fig. 5B**). Leading edge proteins and other enriched Hallmark pathways from this analysis are shown in **Table S8**.

**Fig. 5.**
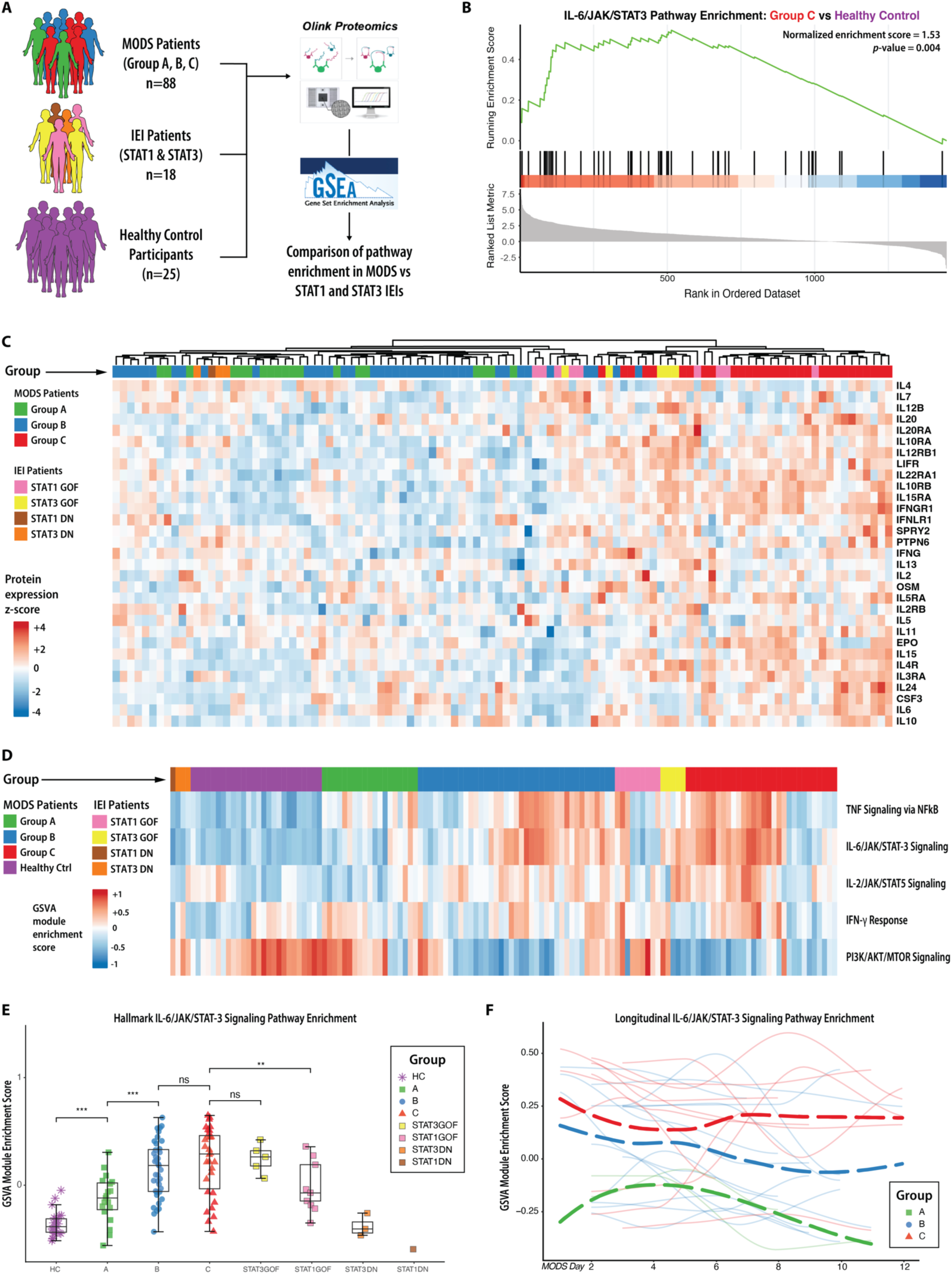
STAT3 signaling in some patients with MODS exceeds that seen in IEI patients with constitutively activated STAT3 signaling due to STAT3 GOF mutation. **(A)** Study schematic for this analysis, which includes 88 patients with MODS, 18 patients with IEI, and 25 HC participants. Normalized protein expression was analyzed using GSEA and GSVA to determine similarities in protein expression between subgroups and monogenic IEI. **(B)** GSEA analysis of population-level enrichment of the Hallmark IL-6/JAK/STAT3 signaling pathway in Group C patients at MODS onset compared to HC participants. **(C)** Heatmap showing normalized protein expression of 31 KEGG JAK/STAT target proteins among 88 patients at MODS onset and 18 patients with inborn errors of immunity (IEI) characterized by STAT1 and STAT3 gain-of-function (GOF) and dominant-negative (DN) mutations. Colored annotation bars indicate IEI diagnosis and MODS subgroup. **(D)** Clustered heatmap of patient-level enrichment of five inflammatory pathways in MODS patients and IEI patients using GSVA. Colored annotation bars indicate IEI diagnosis and MODS subgroup. **(E)** Comparison of Hallmark IL-6/JAK/STAT3 signaling module enrichment scores by IEI diagnosis and MODS subgroup. Comparisons were made between groups as shown and determined by Wilcoxon rank sum test. **(F)** Longitudinal IL-6/JAK/STAT3 module enrichment scores among 26 patients with ≥3 sample timepoints. Solid lines represent individual patient trajectories; dashed lines represent LOESS regression lines for each subgroup.

To assess expression of STAT target proteins across MODS and IEI patients, we identified 31 measured proteins from the KEGG JAK/STAT gene set and performed unsupervised hierarchical clustering based on normalized protein expression. We assessed group concordance at the first dendrogram bifurcation, which separated the cohort into two clusters. As shown in **Fig. 5C**, STAT1 GOF and STAT3 GOF patients co-localize with Group C patients (*p*<0.0001) while STAT1 DN and STAT3 DN patients co-localize with Group A patients (*p*<0.0001). This protein-level analysis also highlights the biologic heterogeneity within our severity-defined subgroups and the tested IEI.

Having demonstrated enrichment of STAT target proteins in plasma from MODS Group C patients, STAT1 GOF patients, and STAT3 GOF patients, we next used GSVA to assess patient-level enrichment of five inflammatory pathways in MODS and IEI patients. **Fig. 5D** shows a clustered heatmap of module enrichment scores for each patient in the dataset. We noted increased IL-6/JAK/STAT3 module enrichment scores in all STAT3 GOF and some STAT1 GOF patients, while STAT1 DN and STAT3 DN patients had IL-6/JAK/STAT3 module enrichment scores equivalent to healthy control patients. Compared to Group A patients, IL-6/JAK/STAT3 module enrichment scores were increased in Group C patients (*p*<0.001) and STAT3 GOF patients (*p*=0.001) (**Fig. 5E**). There was no significant difference between STAT3 GOF and Group C patients, and many patients had enrichment scores which exceeded the median score of STAT3 GOF patients. Collectively, these findings support the conclusion that Group C patients exhibit a degree and pattern of pathologic IL-6/JAK/STAT3 pathway activation that mirrors, and in some cases exceeds, that observed in individuals with monogenic STAT3 gain-of-function mutations.

Finally, we examined the temporal dynamics of STAT3 pathway activation by comparing longitudinal IL-6/JAK/STAT3 module enrichment scores among the subset of 26 patients with ≥3 serial plasma samples. Group C patients exhibited the highest enrichment scores at MODS onset, and these remained persistently elevated throughout the first two weeks of illness (**Fig. 5F**). In contrast, enrichment scores in Group B patients declined gradually over time, while scores in Group A patients showed a rapid and sustained decrease. Notably, increases in IL-6/JAK/STAT3 signaling over time were rare: only two Group C patients demonstrated a late rise in enrichment scores, with low initial activation followed by marked elevation approximately one week after MODS onset. These findings suggest that STAT3 hyperactivation in Group C is both an early and durable feature of disease, supporting its role as a driver of immune dysregulation rather than a transient host response.

### STAT1 and STAT3 signaling are associated with T cell immunometabolic dysregulation in MODS

Having demonstrated STAT3 pathway signaling in Group C patients at levels comparable to or exceeding those seen in patients with known STAT3 GOF patients, we next hypothesized that plasma STAT3 hyperactivation would result in immunometabolic dysregulation within lymphocytes from Group C patients. To test this hypothesis, we performed single cell RNA sequencing using cryopreserved PBMCs obtained at MODS onset for Group C patients (n=9) and pediatric HC participants (n=3). Plasma IL-6/JAK/STAT3 module enrichment scores were significantly different between the 9 patients with MODS and 3 HC participants (0.331 vs. −0.494, *p*<0.0001) selected for the transcriptomics experiment.

A schematic of our experimental approach is shown in **Fig. 6A**. We sorted live CD45^+^ PBMCs and then profiled 10,000 cells per patient using 5’ RNA tag single cell sequencing approach (scRNA-seq; 10X Genomics, Pleasanton, CA). Libraries were sequenced to a depth of ∼30,000 reads per cell on a Novaseq S2 (Illumina, San Diego, CA). Transcripts were aligned to the GRCh38 reference genome using Cell Ranger v8.0. After quality control using SoupX (*67*) and DoubletFinder (*68*) and integration using RPCA via Seurat v5 (*69*), cell identities were inferred using ScType (*70*) and refined using Azimuth (*71*) and T cell phenotypes from Giles et al (*72*). Using this approach, we identified 15 immune cell populations by transcriptional profile for downstream analysis. We applied UMAP dimensionality reduction (*73*) and subsampled 30,000 cells from each group (HC and Group C) for visualization of immunophenotypic differences (**Fig. 6B**). Differences in cell populations between HC participants and Group C are quantified in stacked bar plots (**Fig. 6C**) and largely mirror the differences in lineages seen in our flow cytometry analysis, redemonstrating the altered immune cell composition in Group C patients.

**Fig. 6.**
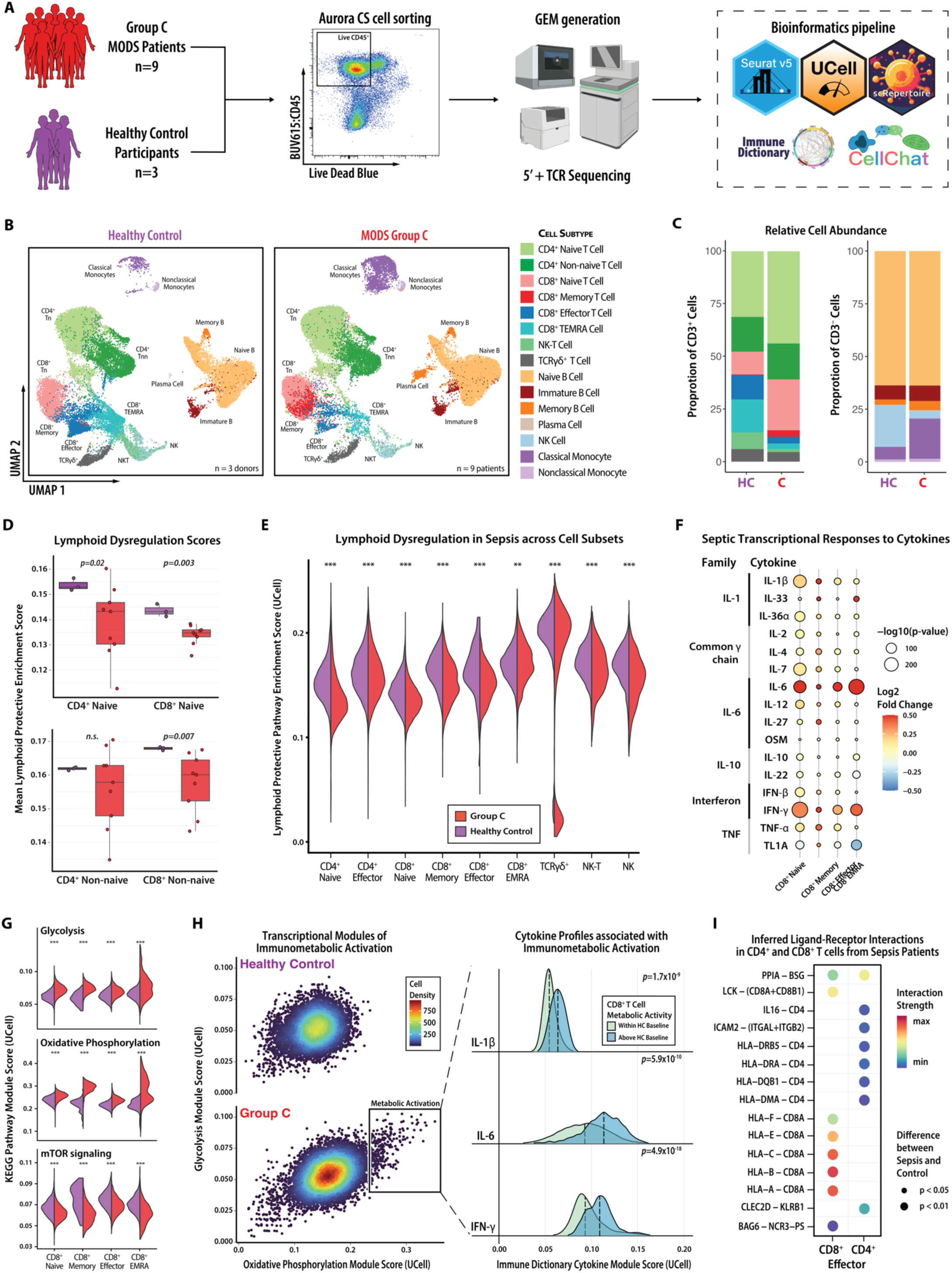
STAT1 and STAT3 signaling promote T cell immunometabolic dysregulation. **(A)** Study schematic for the scRNA-seq experiment, including 9 patients with from MODS Group C and 3 HC participants. Cells were sorted, sequenced, and analyzed using a bioinformatics pipeline to compute module enrichment scores by condition and cell type, transcriptional responses to cytokines, and inferred ligand-receptor interactions. **(B)** UMAP projection demonstrates differences in lymphoid and myeloid lineages between healthy and Group C patients. **(C)** Stacked bar plots show differences in CD3^+^ and CD3^-^ cell proportional abundance by group corresponding to the metaclusters in the UMAP projections to the left. **(D)** Pseudobulk lymphoid protective module enrichment scores are lower in CD4⁺ naïve, CD4⁺ non-naïve, and CD8⁺ non-naïve T cell populations. Module scores were calculated with UCell, and comparisons were determined by Wilcoxon rank sum test (*** reflects *p*<10^-16^). **(E)** Lymphoid protective module scores across nine lymphoid subsets demonstrate widespread transcriptional dysregulation in Group C lymphocytes. Comparisons were determined by Wilcoxon rank sum test. **(F)** Transcriptional responses to cytokines inferred using the Immune Dictionary framework reveal increased signaling activity in Group C patients in response to multiple pro-inflammatory cytokines. Module scores were calculated with UCell. Median fold change represents Group C enrichment over HC enrichment. Comparisons between C and HC were determined by Wilcoxon rank sum test. **(G)** Compared to HC participants, CD8⁺ T cells from Group C patients exhibit increased transcriptional activity in glycolysis and oxidative phosphorylation pathways and suppressed mTOR signaling across all CD8⁺ differentiation states. Comparisons were determined by Wilcoxon rank sum test (*** reflects *p*<10^-16^). **(H)** Bivariate enrichment scores for glycolysis and oxidative phosphorylation modules in CD8⁺ T cells show a distinct population of metabolically activated cells in Group C. Ridge plots demonstrate that enrichment scores for IL-6, IFN-γ, and IL-1β cytokine modules are significantly elevated in these cells. Comparisons between activated and non-activated cells were made using multivariable mixed-effects modeling with cytokine as fixed effects and patient identity incorporated as a random effect. **(I)** Ligand-receptor interaction analysis identified increased interaction strength between MHC class I molecules on monocytes and CD8A on CD8^+^ T cells in Group C patients compared to healthy controls, suggesting increased CD8^+^ activation. Color and size of dot indicate the interaction strength and *p*-value of the inferred cell-cell communication networks from between Group C and HC.

Building on our flow-cytometry evidence of T-cell depletion and activation and our plasma evidence of STAT3 hyperactivation, we first tested whether a validated transcriptomic marker of dysfunctional lymphoid immunity was similarly perturbed in Group C patients. We applied a transcriptional framework developed by the SUBSPACE consortium (*74*) which identified a “lymphoid protective” gene set derived from adult patients with sepsis. Lower scores on this module denote a “lymphoid detrimental” transcriptional program associated with T cell dysfunction and poor outcomes in adult sepsis. We calculated “lymphoid protective” module enrichment scores at the single-cell level using UCell (*75*) and generated pseudobulk profiles for statistical analysis to control Type 1 error (*76*). Consistent with our protein-level findings, Group C patients exhibited significantly lower lymphoid protective scores compared to HC participants in CD4+ naïve, CD4+ non-naïve, and CD8+ non-naïve populations (**Fig. 6D**).

To broaden our assessment of transcriptional dysregulation, we extended our analysis to include per-cell lymphoid protective module enrichment scores across 9 major lymphoid subsets (**Fig. 6E**). Group C patients exhibited significantly lower module scores across all subsets, with the most pronounced deficits observed in the CD8⁺ compartment. Together, these results suggest that lymphoid dysregulation in Group C is not restricted to numerical depletion but reflects widespread changes to immune regulatory gene programs necessary for effective immune surveillance.

To infer which cytokines may drive the immune dysregulation observed in Group C lymphocytes, we leveraged the Immune Dictionary framework (*77*), which enables cell-type specific identification of cytokine activity based on downstream gene expression patterns. As shown in **Fig. 6F**, Group C patients exhibited significantly increased transcriptional responses to multiple pro-inflammatory cytokines in comparison to HC participants, including IL-6 (which predominantly activates STAT3) and IFN-γ (which predominantly activates STAT1). These findings suggest that sustained exposure to a proinflammatory cytokine milieu, particularly IL-6 and IFN-γ, may drive the transcriptional reprogramming observed in lymphocytes from Group C patients.

To understand the effects of sepsis on T cell immunometabolism, we calculated KEGG pathway module enrichment scores for glycolysis, oxidative phosphorylation, and mTOR signaling pathways using UCell (*75*) as detailed in Methods. As shown in **Fig. 6G**, CD8⁺ T cells from Group C patients exhibited increased transcriptional activity in glycolysis and oxidative phosphorylation pathways compared to HC participants across all differentiation states. Conversely, mTOR signaling was suppressed across CD8⁺ subsets from Group C, a pattern suggesting cytokine-driven repression of the mTOR pathway, which may affect T cell function (*78–80*).

Given that IL-6 and IFN-γ exert complex effects on immunometabolic state and mTOR signaling, we next assessed whether exposure to these signals explained the metabolic phenotype observed in **Fig 6G**. To test whether specific cytokines underpinned this bioenergetic shift, we modeled single-cell Hallmark glycolysis and oxidative phosphorylation module scores jointly (**Fig. 6H**). This revealed a distinct subset of Group C CD8⁺ T cells with increased glycolysis and oxidative phosphorylation activity (labeled as “metabolic activation”). In multivariable mixed-effects logistic regression analysis, enrichment scores for IL-6, IFN-γ, and IL-1β transcriptional responses are significantly increased in metabolically activated CD8^+^ T cells as compared to cells within the HC baseline range (all *p*<0.001), as shown in the ridge plots in **Fig. 6H**.

Finally, to further characterize the amplified activation state of CD8⁺ T cells in Group C patients, we applied CellChat (*81*) to the scRNA-seq dataset to identify predicted cell-to-cell communication events via curated ligand-receptor pairs and co-receptor interactions that modulate signaling. As shown in **Fig. 6I**, Group C patients exhibited significantly stronger inferred signaling interactions between monocyte-expressed HLA class I molecules and *CD8A* co-receptor on CD8⁺ T cells, compared to healthy controls (p < 0.01). This enhanced antigen presentation signature suggests increased activation of CD8⁺ effector T cells, reinforcing our transcriptional and metabolic findings and highlighting the potential role of myeloid–lymphoid crosstalk in perpetuating immune dysregulation.

### T cell activation in MODS occurs without antigen-specific clonal expansion

Having demonstrated lymphoid dysregulation and cytokine-linked metabolic reprogramming in Group C patients, we next investigated whether these immunometabolic changes reflected antigen-specific immune responses or non-specific bystander activation. To do so, we analyzed T cell receptor (TCR) repertoires captured alongside our scRNA-seq data using 5′ tag-based single-cell V(D)J sequencing (10X Genomics, Pleasanton, CA). VDJ libraries were sequenced to a depth of ∼10,000 reads per cell on a Novaseq S2 (Illumina, San Diego, CA) and processed with Cell Ranger v8.0, aligning to the GRCh38 reference genome. TCR data were integrated with scRNA-seq analyses in Seurat using the scRepertoire package (*82*). We used this dataset to test the hypothesis that T cell activation in Group C patients represented antigen-independent “bystander” inactivation as opposed to antigen-specific activation in response to a pathogen.

To computationally assess antigen specificity, we first quantified clonal expansion across the TCR repertoire. Group C patients exhibited no evidence of dominant clones: no single clone occupied more than 1% of the repertoire, and most clones were present at frequencies below 0.01% (**Fig. 7B**). This lack of clonal expansion suggests that T cell activation in Group C is not driven by a focused antigen-specific response.

**Fig. 7.**
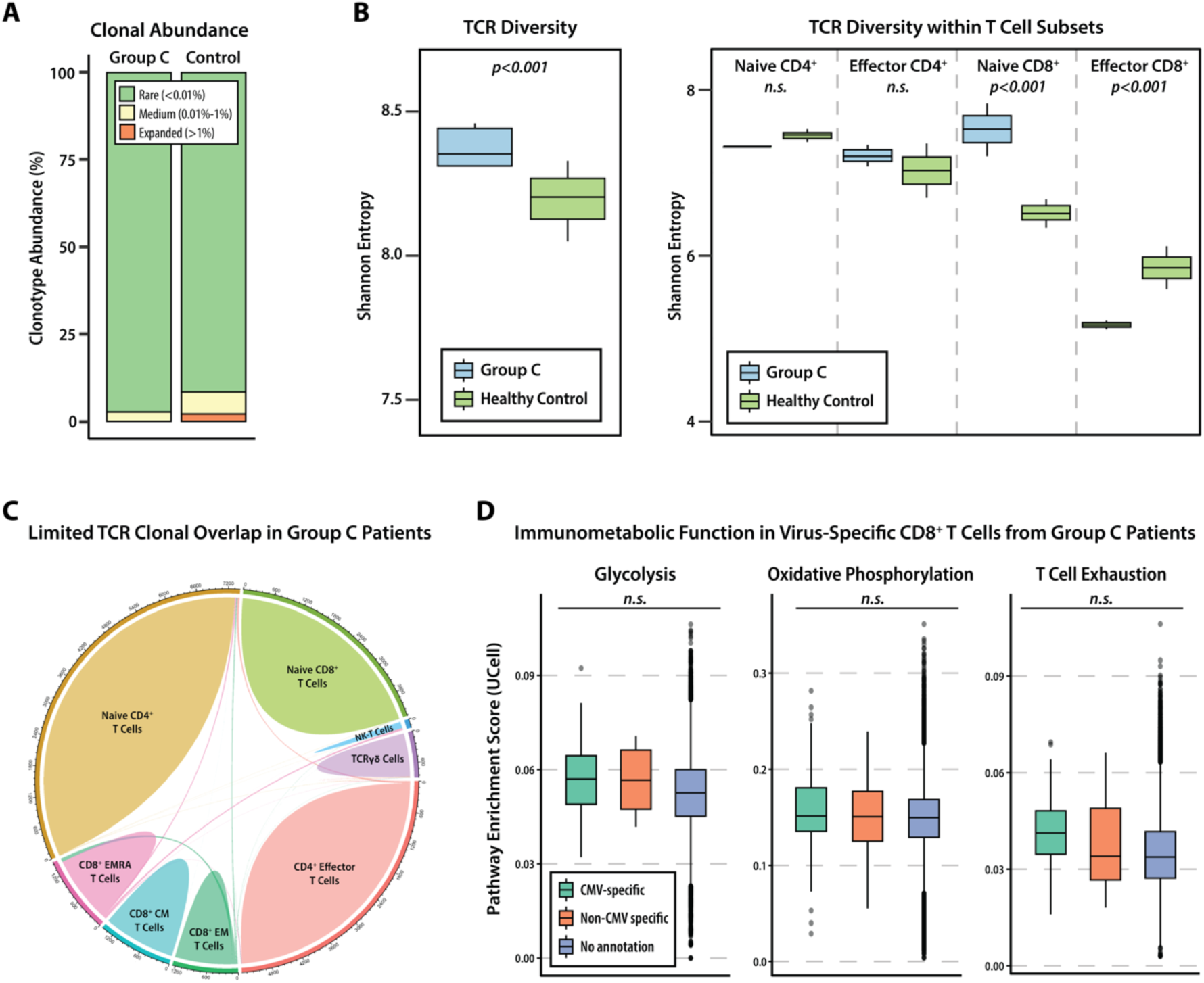
Polyclonal, antigen-independent T cell activation in Group C patients. **(A)** Analysis of clonal abundance showed no evidence of dominant clonotypes in Group C patients, with nearly all clones classified as rare (<0.01% abundance). **(B)** TCR diversity, assessed using the Shannon index, was significantly higher in Group C patients compared to healthy control participants (p<0.001), driven primarily by naïve CD8^+^ T cell. Comparisons were determined by Wilcoxon rank sum test. **(C)** Chord diagram illustrating limited TCR clonal overlap across T cell subsets in Group C patients, consistent with a lack of antigen-driven clonal expansion. **(D)** Immunometabolic pathway analysis in virus-specific CD8^+^ T cells revealed no significant differences in transcriptional signatures of glycolysis, oxidative phosphorylation, and T cell exhaustion among cells with CMV-specific, non-CMV-specific, and unannotated TCRs, suggesting that immunometabolic dysfunction in Group C patients is not driven by antigen-specific T cells. Pathway enrichment scores were calculated using UCell and compared across groups by Kruskal-Wallis test.

We next assessed paired TCR diversity using the Shannon index, which accounts for both the richness (number of unique clonotypes) and evenness (relative abundance) of clonotypes. As shown in **Fig. 7C**, Group C patients displayed significantly higher Shannon diversity compared to healthy controls (*p*<0.001), consistent with broad, non-specific T cell activation. Subset-level analysis revealed that TCR diversity remained comparable to controls in naïve and effector CD4⁺ T cells, but was increased in naïve CD8⁺ T cells and decreased in effector CD8⁺ T cells (both *p*<0.001), suggesting that alterations to the TCR repertoire are specific to CD8⁺ lineages.

To determine whether CD8^+^ T cell activation reflected antigen-specific clonal expansion, we visualized clonotype overlap across T cell subsets. As shown in **Fig. 7D**, Group C patients exhibited minimal overlap between naïve, central memory, effector memory, and EMRA CD8^+^ T cell populations, again providing evidence against expansion of antigen-specific clones and supporting a model of bystander activation.

Finally, we investigated whether immunometabolic pathway enrichment differed by antigen specificity predicted *in silico* using TReX (*83*). We stratified CD8^+^ T cells by cytomegalovirus (CMV)-specific, non-CMV-specific, and unannotated TCRs and compared glycolysis, oxidative phosphorylation, and T cell exhaustion gene set enrichment scores across these groups using UCell. No significant differences were observed across these groups (**Fig. 7E**), suggesting that virus-specific CD8⁺ T cells do not drive the immunometabolic changes observed in Group C.

Taken together, these findings suggest that T cell activation and immunometabolic dysregulation in Group C patients occurs without reliance on antigen-specific clonal expansion associated with pathogen-specific responses. These data support a model of cytokine-driven, bystander T cell activation in the setting of STAT3 and STAT1 hyperactivation rather than a conventional antigen-specific immune response.

### IL-6 and IFN-γ signaling drive aberrant STAT pathway activation and T cell dysfunction in MODS

Building on our single cell transcriptional analyses, which demonstrated antigen-independent T cell activation and marked enrichment of IL-6 and IFN-γ response modules in CD8^+^ T cells from Group C patients, we next sought to test whether these cytokines drive functional changes in signaling at the protein level. To do so, we employed an optimized a 13-marker T cell phosphoflow cytometry panel (**Table S9**) which allows for identification of T cells and includes phospho STAT (pSTAT) antibodies that recognize pSTAT1, pSTAT3, and pSTAT5 in addition to measuring total STAT proteins (*84*).

We first examined basal phosphorylation of STAT1 and STAT3, key transcription factors downstream of IFN-γ and IL-6 signaling, respectively, though their use as heterodimers and homodimers downstream of cytokine receptors is complex and includes many other cytokines. Compared to HC, Group C patients at baseline exhibited higher expression of pSTAT1 (p=0.041) and pSTAT3 (p=0.026) in non-naïve CD8+ T cells (**Fig. 8A**). This finding is consistent with the demonstrated elevated plasma cytokine levels, as well as our transcriptional cytokine response analysis, suggesting that IL-6 and IFN-γ exert a combined effect on CD8^+^ T cells in Group C patients.

**Fig. 8.**
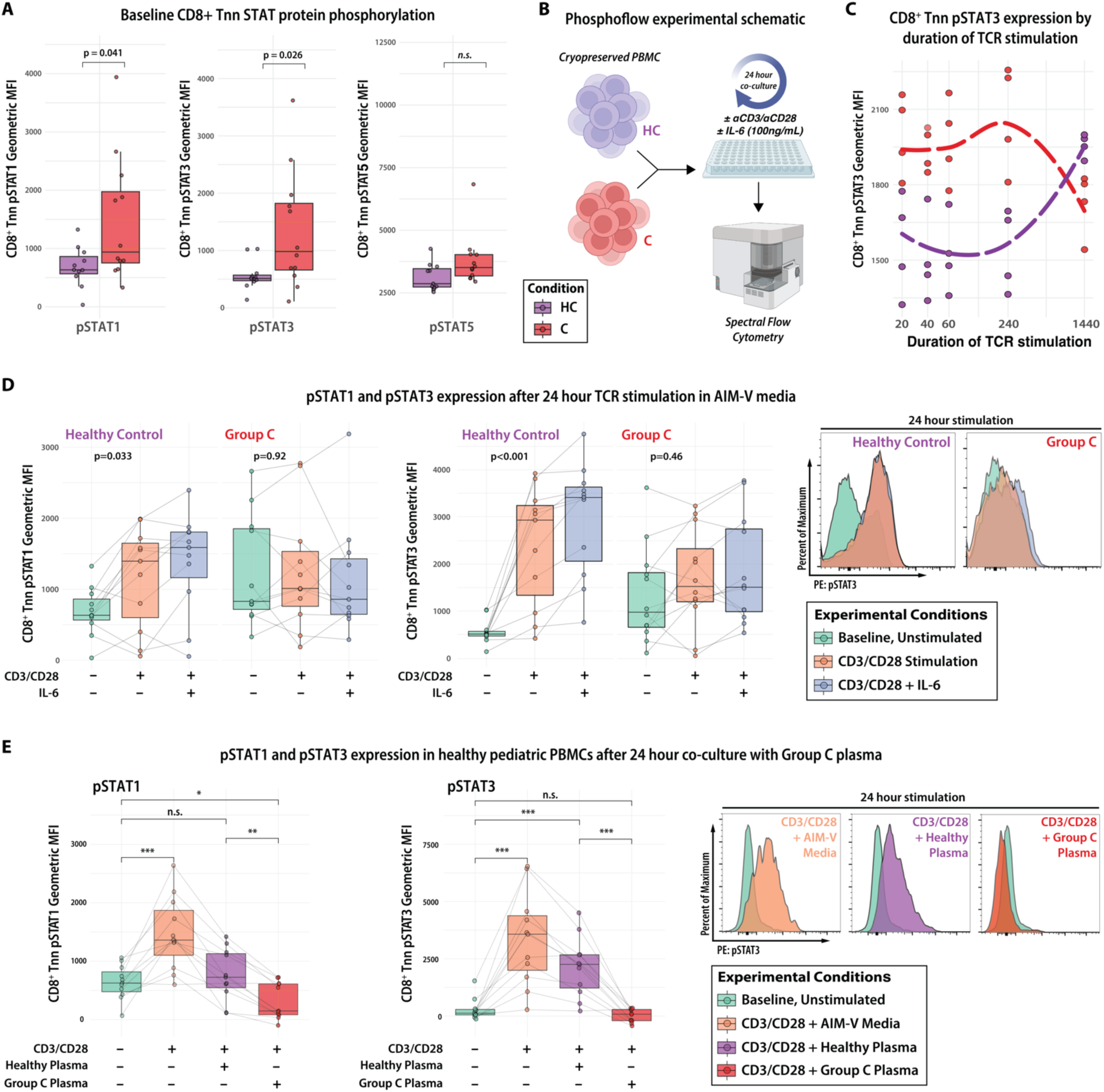
IL-6 and IFN-γ signaling drive aberrant STAT pathway activation and T cell dysfunction. **(A)** Baseline phosphorylation of STAT1 and STAT3 was elevated in non-naïve CD8⁺ T cells from Group C patients compared to healthy controls (HC), consistent with chronic cytokine stimulation (*p*=0.041 and *p*=0.026, respectively). Comparisons were determined by Wilcoxon rank sum test. **(B)** Schematic of the ex vivo TCR stimulation model. PBMCs were cultured with plate-bound anti-CD3 and soluble anti-CD28 for up to 24 hours. **(C)** Time-course of pSTAT3 expression following TCR stimulation. Group C CD8⁺ T cells showed elevated early (0-4hr) pSTAT3 expression, followed by a decline by 24 hours, consistent with cytokine-induced signaling desensitization. Conversely, HC cells increased pSTAT3 expression over time. **(D)** TCR stimulation (±IL-6) induced robust pSTAT1 and pSTAT3 responses in HC samples (*p*=0.033). In contrast, Group C CD8⁺ T cells failed to upregulate pSTATs under the same conditions, indicating functional refractoriness. Comparisons were determined using the Kruskal-Wallis test. Representative histograms illustrate impaired pSTAT3 induction in Group C. **(E)** Plasma from Group C patients suppressed TCR-induced pSTAT1 and pSTAT3 phosphorylation in healthy donor PBMCs. In contrast, culture in complete media or media with HC plasma led to increased pSTAT3 expression with TCR stimulation (both *p*<0.001). Comparisons were determined by Wilcoxon rank sum test. Representative histograms illustrate inhibition of pSTAT3 in healthy cells exposed to Group C plasma.

Given the elevated baseline STAT signaling in Group C, we next assessed how this cytokine exposure affected CD8⁺ T cell responsiveness to receptor-mediated stimulation. As illustrated in **Fig. 8B**, PBMCs from patients in Group C were thawed and cultured for up to 24 hours with plate-bound anti-CD3 and soluble anti-CD28 antibodies.

Group C CD8⁺ T cells initially exhibit higher pSTAT3 expression compared to HC during the first 4 hours of stimulation (**Fig. 8C**). However, by 24 hours pSTAT3 levels in Group C CD8⁺ T cells sharply declined, whereas healthy control cells maintained or increased STAT3 phosphorylation over the same time course. This pattern suggests an inability to amplify STAT3 signaling beyond baseline elevation in pSTAT3 in Group C patients, which could reflect reduced ongoing phosphorylation, accelerated dephosphorylation, or a combination of both mechanisms.

To quantify STAT pathway signaling capacity in Group C patients, we next compared the ability of Group C CD8⁺ T cells to respond to TCR stimulation alone or in combination with IL-6. As shown in **Fig. 8D**, HC cells upregulated both pSTAT1 and pSTAT3 following aCD3/aCD28 stimulation, and IL-6 further augmented pSTAT3 levels. In contrast, Group C cells failed to increase pSTAT1 or pSTAT3 expression in response to TCR stimulation with or without IL-6. Representative pSTAT3 histograms highlight the lack of response to stimulation in T cells from Group C patients. These results indicate that CD8^+^ T cells from Group C patients are functionally refractory to JAK/STAT signaling downstream of TCR engagement, consistent with cytokine-induced signaling desensitization (*85*).

To determine whether soluble mediators in Group C plasma were sufficient to drive the observed pSTAT1 and pSTAT3 signaling abnormalities, we performed plasma culture experiments using healthy donor PBMCs. As shown in **Fig. 8E**, healthy donor cells have low pSTAT1 and pSTAT3 expression at baseline, and pSTAT3 expression increases significantly when exposed to TCR stimulation in the setting of complete media or media supplemented with healthy control patient plasma (both *p*<0.001). In contrast, co-culture of healthy PBMCs with media supplemented with plasma from Group C patients for 24 hours completely suppressed TCR-induced pSTAT1 and pSTAT3 phosphorylation, maintaining expression at baseline levels. Representative pSTAT3 histograms highlight the inhibitory effect of Group C plasma on pSTAT3 expression in healthy donor cells. Collectively, these findings demonstrate that circulating factors present in Group C plasma are sufficient to inhibit STAT1 and STAT3 signaling in otherwise healthy CD8^+^ T cells.

Together, these findings support the hypothesis that sustained exposure to IL-6 and IFN-γ in Group C patients may drive chronic activation of the STAT1 and STAT3 signaling pathways, leading to a state of functional desensitization to further activation of those pathways in CD8⁺ T cells. Despite elevated basal pSTAT1 and pSTAT3 levels, Group C T cells fail to respond appropriately to subsequent TCR-mediated stimulation, suggesting a state of refractory responses to cytokine signals. Culture of HC cells with media supplemented by plasma from Group C patients supports the hypothesis that T cell dysfunction may be extrinsically imposed by the inflammatory milieu, rather than due to intrinsic defects in the cells themselves. These data implicate IL-6 and IFN-γ–driven STAT1 and STAT3 signaling as central mediators of T cell dysregulation in Group C, potentially defining an endotype which drives dysregulation in this subgroup, and provide a mechanistic link between systemic inflammation, transcriptional rewiring, and impaired adaptive immunity in MODS that may be therapeutically tractable and requires further study.

## DISCUSSION

In a prospective cohort of children admitted to a quaternary care PICU with MODS from diverse etiologies, we have defined a distinct immunologic endotype characterized by persistent STAT1/STAT3 hyperactivation and T cell immunometabolic dysfunction in a subset of patients with the highest severity of illness and worst prognosis. These patients can be identified by a 24-protein signature at MODS onset that is agnostic to underlying etiology, suggesting a convergent immunopathologic trajectory across diverse infectious and non-infectious insults. Because this signature relies solely on soluble biomarkers, it may be amenable to future development as a rapid clinical assay, enabling targeted studies of endotype-guided precision immunomodulation.

Multi-modal immune profiling identified that Group C is characterized by exposure to increased levels of IL-6 and IFN-γ, leading to chronic activation of STAT1 and STAT3 signaling pathways. Single-cell RNA sequencing revealed broad suppression of lymphoid protective gene programs (*74*) and transcriptional signatures of T cell exhaustion, metabolic stress, and cytokine-driven activation, particularly within CD8⁺ subsets. Despite high expression of activation markers by flow cytometry and scRNA-seq, our TCR repertoire analysis revealed no evidence of clonal expansion, consistent with antigen-independent, bystander T cell activation. Functionally, CD8^+^ T cells from Group C patients exhibited increased baseline STAT1 and STAT3 phosphorylation yet failed to respond to TCR-mediated stimulation. These findings suggest a state of cytokine-induced desensitization or functional exhaustion, which was recapitulated in healthy PBMCs following exposure to Group C plasma—confirming an extrinsic, cytokine-driven mechanism of T cell dysfunction.

Our highly granular approach to immune profiling yields new insights into the pathobiology of pediatric sepsis and MODS and identifies a potential targetable mechanism of immunopathology—IL-6/IFN-γ–mediated STAT1/STAT3 hyperactivation that drives T cell immunometabolic dysregulation—in critically ill children with and without sepsis. To our surprise, we identified features of immune dysregulation in many patients with MODS who did not have suspected infection, and all three subgroups include patients with and without sepsis. Our findings redemonstrate the association between T cell dysfunction, mitochondrial dysfunction, and clinical outcomes in pediatric sepsis (*23–26*), and suggest a potential causative molecular mechanism for sepsis-associated T cell immunometabolic dysregulation. As other investigators have shown, the highest severity patients in our cohort have innate (*17, 21*) and adaptive (*18, 20*) immune dysfunction and mitochondrial dysfunction (*25, 26, 86*). Recent human and murine data identify a role for STAT3 signaling in the pathogenesis of sepsis (*63–65*), and our results concordantly demonstrate associations between STAT3 signaling, clinical outcomes, and features of immune dysregulation implicated in pediatric sepsis pathobiology.

Pathologic STAT1 and STAT3 signaling abnormalities in this endotype are potentially targetable through approved anti-cytokine antibodies, JAK inhibitors, and experimental STAT inhibitors (*87*), and thus represent viable future targets for precision medicine studies, and potential future therapeutics, in pediatric patients with sepsis. Supporting this hypothesis, during the COVID-19 pandemic some patients requiring supplemental oxygen were noted to benefit from a combination of dexamethasone and a JAK inhibitor (*88*). In randomized controlled trials, use of the JAK inhibitors baricitinib and tofacitinib were associated with reduced mortality (*89–92*), faster recovery (*93*), and lower rates of disease progression (*89*) in multiple adult trials focusing on varying levels of illness severity. Several available JAK inhibitor agents have good bioavailability, rapid onset of action, and short half-lives, features that are favorable for use in the critical care setting. If STAT1/STAT3 hyperactivation is in fact a causal contributor to organ failure in pediatric sepsis, the safety and efficacy of baricitinib in critical COVID-19 suggests a strong rationale for targeting JAK signaling in future pediatric MODS clinical trials.

The longitudinal analysis of protein and pathway expression in this study provides evidence that immune dysregulation present at MODS onset persists for more than a week in many patients. Persistent immune dysregulation is amenable to consideration of a trial of precision therapeutics in which patients could be identified at MODS onset and then assigned to a treatment arm based on biomarker-based prognostic and predictive enrichment strategies. Equally relevant, prognostic subgroups defined at MODS onset demonstrated relatively stable proinflammatory pathway enrichment scores through time, suggesting that subgroup membership is patient/episode-specific and minimally impacted by time from MODS onset.

By incorporating comparisons to rare, monogenic IEI to contextualize complex sepsis biology, we are able to learn from the demonstrated impacts of chronic exposure to amplified signaling in key pathways. Historically identified in children with severe presentations of specific infectious diseases (*94*), IEI are known to be enriched in pediatric patients with COVID-19 (*95–98*), influenza (*99–101*), and sepsis (*38, 39, 102*). Genotype-phenotype associations between monogenic IEI and disease susceptibility may offer insights into causal pathobiology of illness which subsequently inform precision therapeutics (*103*). More recently, it has become clear that many IEI are characterized by immune dysregulation (*104*). Our comparison between patients with MODS and patients with gain-of-function and loss-of-function/dominant negative STAT disorders allows us to learn the impact of chronic amplification of these key pathways from human disease and suggests that the extent of STAT1 and STAT3 hyperactivation in the most severe MODS subgroup acutely exceeds that produced by constitutive STAT1 and STAT3 pathway activation.

The cellular correlates of immune dysfunction observed in this study – defined by persistent STAT1 and STAT3 hyperactivation, altered metabolic programming, and impaired responsiveness to TCR stimulation – were reproducible across platforms, including spectral flow cytometry and scRNA-seq. These phenotypic and functional features may also offer candidate functional biomarkers to serve as surrogate endpoints for early-phase interventional studies. For example, *ex vivo* pSTAT signaling assays or CXCL9 levels could potentially be used to evaluate pharmacodynamic responses to JAK inhibition or cytokine blockade, while immune cell metabolic profiling (using Seahorse or SCENITH (*105*) assays) could track immunometabolic recovery following treatment. Ultimately, these findings define a path toward biomarker-guided, mechanism-informed interventions for critically ill children and establish a new translational roadmap for targeting immune dysregulation in pediatric sepsis.

Building on this mechanistic framework, we used penalized regression to identify a prognostic protein signature detectable at MODS onset and demonstrated excellent discrimination using linear discriminant analysis. Elastic net regularization is a common approach applied to feature selection in high-dimensional data which generates a high-performing sparse model with good prediction accuracy (*52*) and has been employed with similar results in adult sepsis studies (*47*). Prior to potential use for prognostication or selection of targeted therapeutics, the proposed parsimonious 24-protein MODS severity model will require prospective validation. It will also benefit from validation across multiple proteomics platforms, as aptamer-based and antibody-based approaches are susceptible to both technical and genetic variation among samples (*106*). A validated severity model derived from the plasma proteome would present an opportunity for reverse translation to the bedside, as clinical labs in many centers perform rapid, quantitative diagnostic testing in-house using multiplex protein assays.

Limitations of our study include an *a priori* sample size that was only powered to resolve 3 proteomic subgroups within a heterogeneous syndrome. This reflects a pragmatic approach to study design and does not preclude the existence of other subgroups and/or endotypes. Additionally, our flow cytometry and transcriptomics experiments relied on cryopreserved PBMC samples. While this approach improved study feasibility and minimized batch effect associated with longitudinal sample collection, it is possible that rare and fragile cell populations may have been impacted by a single freeze/thaw cycle. The use of PBMCs precludes cellular analysis of granulocyte populations, particularly neutrophils, which may be of particular relevance in the setting of STAT pathway hyperactivation. Finally, we conducted our analysis on circulating blood cells and plasma protein expression based on the assumption that circulating immune cells reflect what happens in the injured tissues of patients with MODS, but the functional similarities between circulating immune cells and tissue resident immune cells are not known in this hyperinflammatory setting.

Collectively, our longitudinal multi-omics analysis defines a high-risk endotype in which sustained IL-6 and IFN-γ exposure contributes to aberrant STAT1 and STAT3 signaling, culminating in immune dysregulation and impaired T cell responsiveness. Concordant results from proteomic, cytometric, and transcriptomic assays underscore the biological coherence of this endotype, which is associated with poor prognosis and can be defined at the onset of MODS using a parsimonious plasma proteomic signature. Taken together, these findings provide strong justification for biomarker-guided, mechanism-informed precision therapies in critically ill children and lay the foundation for clinical trials targeting the JAK/STAT axis while also supporting the broader implementation of functional immune profiling as a tool for tailoring immunomodulation in pediatric critical care.

## MATERIALS AND METHODS

### Study Design and Participants

#### Experimental design

After obtaining IRB approval (IRB #19-017032), we enrolled patients with multiple organ dysfunction syndrome (MODS) into a prospective observational cohort study in the 75-bed Pediatric Intensive Care Unit and 38-bed Cardiac Intensive Care Unit at Children’s Hospital of Philadelphia. Both units are combined medical and surgical ICUs which provide comprehensive services to children outside of the neonatal period. Patients with new dysfunction of ≥2 organs defined by modified Proulx criteria (**Table S10**) (*9*) within the last three calendar days and age >40 weeks post-conceptual age and <18 years were eligible for enrollment. Proulx criteria identify MODS through objective, reproducible definitions of seven organ systems, and MODS defined by Proulx criteria is strongly associated with both short- and long-term PICU outcomes (*107*). Exclusion criteria included limitations of care orders at the time of eligibility, clinical suspicion for brain death, and prior enrollment. This study co-enrolled with the multicenter PediAtric ReseArch of Drugs, Immunoparalysis and Genetics during MODS (PARADIGM) study, with shared inclusion/exclusion criteria and case report form but independent biospecimen collection, processing, and analysis. Patient (or legal guardian) consent (and assent, if appropriate) were obtained prior to study enrollment in accordance with our IRB approved protocol. We enrolled June 2020 to December 2022, when we reached our prespecified enrollment target of 88 patients, which we expected to achieve 90% power to detect moderate differences (Cohen’s d = 0.5) among three subgroups, based on pilot flow cytometry and proteomics data. Longitudinal whole blood samples were obtained twice weekly in sodium heparin tubes from onset of MODS through death or resolution of all organ failure.

#### Clinical metadata

Clinical metadata were abstracted by a nurse research coordinator using a standardized case report form into REDCap (*108*) for the parent PARADIGM study. Data quality was verified through manual and automated queries. MODS onset was defined as the calendar day that a patient developed new dysfunction of ≥2 organs defined by modified Proulx criteria **(Table S10)** (*9*). MODS inciting diagnosis was abstracted from attending physician daily progress notes and coded as “sepsis,” “trauma,” “cardiopulmonary bypass,” or “non-infected.” Sepsis was identified as the MODS inciting diagnosis based on clinical suspicion for infection by the primary team and positive microbiologic or virologic testing. Patients with “culture negative sepsis” were classified as sepsis if they developed organ dysfunction in the setting of a clinical suspicion for infection and received sepsis therapies, in concordance with pediatric sepsis guidelines (*109, 110*). Immunocompromised status was defined by diagnosis of active malignancy, prior hematopoietic cell transplant, or primary immune deficiency syndrome. Daily data were extracted from MODS onset through day +28 to allow for daily calculation of the PELOD-2 organ dysfunction score (*13*). We defined cumulative PELOD-2 score through day +28 as our primary outcome because it incorporates both degree and duration of organ failure into a composite outcome variable which is associated with meaningful differences in long-term mortality (*111*) and health related quality of life (*112*) in pediatric patients with sepsis. For each patient and healthy control participant, a one-way hash function was used to generate a “hashed patient ID” number which cannot be linked to patient records or used to identify patients by anyone outside of the study team. These hashed patient IDs are included in supplemental tables where required.

#### Healthy control participant samples

Cryopreserved PBMC and heparin plasma samples from 25 pediatric participants without immunological disease were identified from an existing pediatric biorepository (IRB #18-015920) to serve as a healthy control group. Participant (or legal guardian) consent (and assent, if appropriate) were obtained prior to study enrollment in accordance with our IRB approved protocol. Age range and sex of healthy control participants are shown in **Table S11**.

#### Inborn errors of immunity patient samples

Heparin plasma samples from patients with STAT1 gain-of-function (9), STAT1 dominant-negative (1), STAT3 gain-of-function (5), and STAT3 dominant-negative (3) mutation were identified from collaborators and used as comparators for MODS patients with dysregulated STAT3 signaling. These participants were consented in accordance with local IRB protocols and samples were shared through collaborative research agreements (IRB #18-015920). IEI samples were collected during outpatient appointments from patients without evidence of active infection at the time of sampling.

### Study Procedures

#### Sample processing

Biospecimens were collected starting within 48 hours of MODS onset and continued twice weekly through death, recovery from all acute organ failures, or until six samples were obtained. Blood samples were obtained in heparin plasma tubes by clinical research coordinators and processed on site within one hour of acquisition. Platelet poor plasma was separated by centrifugation and snap frozen on dry ice. Peripheral blood mononuclear cells (PBMCs) were isolated using SepMate (STEMCELL Technologies, Vancouver, Canada) density gradient centrifugation using Lymphoprep media. Whole blood was mixed 1:1 with PBS and layered onto Lymphoprep gradient. SepMate tubes were centrifuged and the buffy coat suspension was spun down for cell isolation. ACK lysis was completed and cells were resuspended in complete RPMI (cRPMI; RPMI 1640 supplemented with 10% FBS, 1% L-Glutamine, 1% Pen-Strep) for counting prior to cryopreservation in 500μl of freezing media (90% FBS, 10% DMSO).

#### Plasma proteomics by proximity extension assay

For each patient and timepoint, 100μl of heparin plasma was submitted to Olink Proteomics (Uppsala, Sweden) for analysis on the Explore 1536 proximity extension assay (PEA) platform. PEA is a dual-recognition assay which uses paired antibodies labeled with DNA oligonucelotides to identify and quantify protein expression through Next Generation Sequencing (*49*). After normalization and quality control, protein concentrations are reported in log2-transformed Normalized Protein eXpression (NPX) units. In total, we analyzed 229 samples from 131 patients across three experiments using 8 shared bridging samples for between-plate-normalization. 1448 proteins met quality control thresholds in all three experiments and were included in downstream analyses (**Table S12**). For analysis of IEI samples, plasma proteins were measured using the Olink Explore 384 Inflammation panel (**Table S7**), a select set of proteins also measured on the Olink Explore 1536 panel for MODS patients.

#### Spectral flow cytometry staining and acquisition

Cryopreserved PBMCs were thawed in 10mL cRPMI and 1×10^6^ cells per sample were plated in a 96-well round-bottom plate. Cell pellets were sequentially incubated with live/dead blue with Fc block, surface antibody stain with Brilliant Stain buffer, permeabilization reagent, and intracellular antibody stain with Brilliant Stain buffer. After staining, cell pellets were resuspended in 1.6% PFA and held at 4°C until acquisition on a Cytek Aurora spectral flow cytometer (Cytek Biosciences, Fremont, CA) the following morning. In total, we analyzed 303 samples from 113 patients across eight experiments. Please see **Table S2** for details of flow cytometry antibodies and buffers and **Supplemental Methods** for complete staining protocol.

#### Flow cytometric cell sorting for scRNA-seq

Cryopreserved PBMCs were thawed in 10mL cRPMI and 1×10^6^ cells were transferred to a new tube for staining. Cell pellets were incubated with 100μl mixture of live/dead blue and CD45 antibody, then washed and passed through a 35µm nylon mesh cell strainer prior to acquisition. Using a Cytek Aurora CS cell sorter (Cytek Biosciences, Fremont, CA), live singlet CD45^+^ cells were sorted into 1.5ml Eppendorf tubes prefilled with 500μl of PBS + 20% FBS. Sorted cells were washed with 500μl PBS + 10% FBS twice to remove sheath fluid EDTA prior to 10X processing. Please see **Supplemental Methods** for complete FACS staining protocol.

#### Single cell RNA and TCR sequencing (scRNA-seq and TCR-seq)

We performed joint single-cell transcriptomic and T cell receptor (TCR) sequencing using the 10X Genomics Chromium Next GEM Single Cell 5′ Kit v2 and Single Cell Human TCR Amplification Kit (10X Genomics, Pleasanton, CA) on sorted PBMC samples (see above) from 9 patients in MODS Group C and 3 pediatric HC participants. We selected samples for this experiment at random from Group C and HC cohorts to avoid bias. Single-cell isolation and library preparation were performed in the Center for Applied Genomics at Children’s Hospital of Philadelphia. Sequencing was performed using the Illumina S2 flow cell (Illumina, San Diego, CA). Demultiplexing and alignment to the GRCh38 reference genome were carried out using the Cell Ranger v8.0 pipeline (10X Genomics), generating both transcriptomic and V(D)J clonotype data for downstream analysis.

#### Phosphoflow Cytometry

Cryopreserved PBMCs were thawed in 10mL cRPMI and 2.5×10^5^ cells per well were plated in a 96-well flat-bottom plate previously coated with anti-human CD3 antibody solution. Cell pellets were sequentially stimulated with anti-human CD28 antibody and subsequently stained with surface antibodies in Brilliant Stain buffer, fixed in 1.6% PFA, and held at −20°C in methanol overnight. Cells were then stained with intracellular antibodies in Brilliant Stain buffer, resuspended in 1.6% PFA, and held at 4°C until acquisition on a Cytek Aurora spectral flow cytometer (Cytek Biosciences, Fremont, CA) the following morning. Please see **Table S9** for details of flow cytometry antibodies and buffers and **Supplemental Methods** for complete staining protocol.

### Statistical Analyses

#### Identification of subgroups

We constructed linear mixed effects models for each protein to determine the association between normalized protein expression and cumulative PELOD-2 score, with age and sex modeled as fixed effects and day from MODS onset modeled as a random effect. Proteins which were significantly associated with cumulative PELOD-2 score after Benjamini-Hochberg correction (FDR *p*<0.05) were retained for further analysis. We used consensus clustering to define subgroups and identified optimal *k* via the Monte Carlo reference-based consensus clustering algorithm (*50*) which uses the proportion of ambiguous clustering (PAC) score to test the hypothesis that *k*=*n* clusters is more informative than *k*=1. Optimal number of clusters in the MODS data (k=3) was confirmed by Monte Carlo bootstrapping, elbow method, and gap statistic. The clinical and proteomic datasets contained no missing data elements, thus imputation was not necessary for our analyses.

#### Competing risk survival analysis

We fit a proportional subdistribution hazards regression model to assess the effect of covariates on the subdistribution of death and survival to PICU discharge in a competing risk setting and then estimated the cumulative incidence function from this model for each subgroup and outcome using the Fine and Gray model (*51*). We then tested the hypothesis that the subdistribution hazard differed between Group C and Group A/B for both death and survival to PICU discharge.

#### Model reduction via ordinal elastic net

We defined a parsimonious protein signature to classify subgroups by training an ordinal elastic net model (*53*) for feature selection using the severity-associated proteins previously identified through linear mixed effects modeling. This approach fit a semi-parallel elementwise link multinomial-ordinal regression model with elastic net penalty which defines coefficients for each protein:subgroup and then shrinks the model to a single coefficient for each protein via elastic net penalty to minimize Gini index. We assessed the performance of the parsimonious elastic net protein set to discriminate the three subgroups using principle component analysis, linear mixed effects post-estimation, and polytomous discrimination index (*113*).

#### Identification of spectral flow cytometry metaclusters

After arcsinh scaling (*54*) and quality control with flowAI (*55*), we performed FlowSOM metaclustering (*56*) to identify 14 immune cell populations by surface and intracellular marker expression. PBMCs were first clustered into k=60 FlowSOM metaclusters and then combined in a stepwise fashion to generate the 14 canonical populations identified in the figure and used for downstream analysis. Metacluster similarity was determined by surface and intracellular marker expression and tSNE-CUDA (*57*) proximity and then confirmed with manual gating. Proliferation and activation markers were analyzed using bivariate plots.

#### Pathway enrichment analysis

We first performed pathway enrichment analysis by subgroup by analyzing estimated protein expression in Group C patients (adjusted for age, sex, and PELOD-2 score) with Ingenuity Pathway Analysis (Qiagen) (*58*). We then analyzed pathway expression in the protein dataset in bulk by GSEA (*66*) and at the individual patient level using GSVA (*59*), with a focus on enrichment of five canonical proinflammatory pathways using Human Molecular Signatures Database (MSigDB) Hallmark gene sets (*60*):

“TNF Signaling via NFκB” (HALLMARK_TNFA_SIGNALING_VIA_NFKB),
“IL-6/JAK/STAT3 Signaling” (HALLMARK_IL6_JAK_STAT3_SIGNALING),
“IL-2/JAK/STAT5 Signaling” (HALLMARK_IL2_STAT5_SIGNALING),
“IFN-γ Response” (HALLMARK_INTERFERON_GAMMA_RESPONSE),
“PI3K/AKT/mTOR Signaling” (HALLMARK_PI3K_AKT_MTOR_SIGNALING).

We also applied GSVA to single cell transcriptomics data in a similar approach using UCell (*75*) and extended our analysis to include relevant immunometabolic pathways using MSigDB KEGG and Hallmark gene sets:

“KEGG Glycolysis” (KEGG_GLYCOLYSIS_GLUCONEOGENESIS)
“KEGG Oxidative Phosphorylation” (KEGG_OXIDATIVE_PHOSPHORYLATION)
“KEGG mTOR” (KEGG_MTOR_SIGNALING_PATHWAY)
“Glycolysis” (HALLMARK_GLYCOLYSIS),
“Oxidative Phosphorylation” (HALLMARK_OXIDATIVE_PHOSPHORYLATION).

A list of the gene sets used in GSEA and GSVA analysis in this manuscript are included in **Table S13**. Throughout the manuscript, we refer to Hallmark pathways as “modules” as opposed to “gene sets” because the analytic approach is applied to both proteomics data (via GSVA) and transcriptomics data (via UCell).

#### Single cell transcriptomics analytic pipeline

After sequencing, the reads were aligned to the GRCh38 reference genome using Cell Ranger pipeline from 10x Genomics. The transcriptomic data were analyzed using the Seurat 5.0 preprocessing and integration pipeline (*69*). After review of standard quality control metrics, cells with less than 200 genes, greater than 3 median absolute deviation above the median, and >10% mitochondrial RNA content were removed from downstream analysis. After normalization and scaling using the scTransform function in Seurat, we performed anchor-based RPCA integration across samples and conditions using the top 3000 genes as anchors. Following linear dimension reduction using PCA, the data was clustered using the Louvain algorithm and projected into 2D coordinates using the RunUMAP function in Seurat. The cell identities were then inferred using the scType package (*71*) followed by curated manual annotation using canonical marker genes to define 14 immune cell populations by transcriptional profile. Results of each quality control and integration step are shown in **Fig. S4**. Pseudobulk analysis was performed by condition, sample, and cell type using Seurat’s AggregateExpression function. Differential expression analysis was performed using the FindMarker function in Seurat. Cell-cell communication analysis was performed using CellChat (*81*). To assess associations between effector cell pathways and T cell immunometabolic states, we employed generalized linear mixed-effects models, treating module enrichment scores as fixed effects and patient identity as a random effect.

#### T cell receptor sequencing analytic pipeline

T-cell receptor (TCR) sequencing data were analyzed using the scRepertoire package (*82*). VDJ output from the Cell Ranger pipeline (10x Genomics) were integrated with single-cell RNA sequencing gene expression data using the combineExpression() function. Clonotypes were defined based on CDR3 amino acid sequences and visualized using clonotype homeostasis and proportional abundance plots. Diversity was quantified using Shannon diversity (*114*). To further characterize TCR sequence features, clonotype-level annotation was performed using the TCR Repertoire Explorer (TReX) package (*83*).

#### Immune Dictionary cytokine signature analysis

To assess cytokine-specific transcriptional responses in our scRNA-seq dataset, we applied the Immune Dictionary framework, which maps cytokine signaling to downstream gene expression profiles in specific immune cell types. Single-cell transcriptomic data were scored using curated cytokine-response gene signatures derived from the published experimental datasets (*77*). A list of the gene sets used in this analysis are included in **Table S14**. Module enrichment scores were computed using UCell for each cytokine response transcriptional signature, enabling cell-specific quantification of cytokine signaling activity.

#### Analysis software

All computational analysis was completed using R 4.5.0 and Bioconductor 3.20. Flow cytometry data was processed using FlowJo 10.9 and Omiq.ai. Figures were compiled in Adobe Illustrator.

## LIST OF SUPPLEMENTARY MATERIALS

The Supplementary Materials file includes the following:

- Supplemental methods
- Fig. S1. Supporting evidence for *k*=3 clusters in Fig. 2A.
- Fig. S2. PCA resolving three subgroups based on expression of 24 elastic net derived severity-associated proteins.
- Fig. S3. Representative gating strategy for spectral flow immune phenotyping panel.
- Fig. S4. Results from single cell transcriptomics quality control and integration steps.
- Table S1. 1472 proteins measured in proteomics panel used in MODS and healthy control cohorts.
- Table S2. Antibodies used in 35-marker spectral flow immune phenotyping panel.
- Table S3. PCA loadings in Fig. 1D, using the full proteomics dataset.
- Table S4. Severity-associated proteins identified through linear mixed-effects model after adjustment for age, sex, and day from MODS onset.
- Table S5. Etiology of MODS and computed subgroup for each patient with MODS.
- Table S6. Immunocompromised diagnoses by subgroup.
- Table S7. 368 proteins measured in proteomics panel used in IEI cohort.
- Table S8. Leading edge analysis of protein expression in Group C patients compared to HC participants.
- Table S9. Antibodies used in 13-marker T cell phosphoflow cytometry panel.
- Table S10. Modified Proulx criteria used for screening and enrollment in the MODS cohort.
- Table S11. Age and sex for each healthy control participant.
- Table S12. Final set of 1448 proteins which met quality control thresholds in all three experiments and were included in downstream analyses.
- Table S13. Gene sets used in GSEA and GSVA analysis.
- Table S14. Gene sets used in Immune Dictionary cytokine signature analysis.

## Supporting information

Supplemental Digital Content

Supplemental Large Tables

## ACKNOWLEDGEMENTS

We owe a debt of gratitude to all the patients and families who contributed clinical data and blood samples to this study, and to all involved clinical teams. We thank the many groups throughout CHOP and the University of Pennsylvania who contributed to this study, including the Pediatric Sepsis Program, the Dysregulated Immune Response Team, the Institute for Immunology & Immune Health, and the Department of Anesthesia and Critical Care. In particular, we thank the CHOP clinical research coordinator team (Nola Juste, Stephen Famularo, Teresa Arroyo, Barrington Bucknor, Ryan Burnett, Jenny Bush, Noah Buzinkai, Amanda Bwint, Amanda Cornetta, Shawn Dickey, Mary Ann DiLiberto, Rebecca Douglas, Emily Duffey, Glory Edioma, Emily Kirwin, Cindee Levow, Carly Machon, Alanah McKelvey, Erin Nfonoyim, Kathlyn Phengchomphet, Myooran Sivarupan, Taylor Slocumb, Bryn Spaide, Julia Surow, Samantha Switzer, Derrick Tam, Wandave Tizhe, Krithika Tupil, and Jia Yuan) for screening and enrolling eligible families in the study, the CHOP BioRC Specimen Processing Unit (Richard Tustin III, Annemarie Butler, Kate Kearns, Nicole Sundo, and Molly Tancini) for processing longitudinal human biospecimens, and the CHOP Center for Applied Genomics (Diana Slater, Cuiping Hou, and Mikaela Hufnell) for their assistance with single cell processing and sequencing. We are grateful to the PediAtric ReseArch of Drugs, Immunoparalysis and Genetics during MODS (PARADIGM) study team (Mark Hall and Athena Zuppa) for allowing this study to proceed as a single-center ancillary to their multicenter study, and for generously sharing curated metadata from the parent study. Special thanks to members of the Collaborative Pediatric Critical Care Research Network (CPCCRN), Pediatric Acute Lung Injury and Sepsis Investigators (PALISI) Network, and Pediatric Critical Care and Trauma Scientist Development Program (PCCTSDP) for their advice during the development and conduct of this study.

## Author contributions

Conceptualization: RBL, AFZ, EJW, NJM, SEH

Methodology: RBL, JCF, NY, MK, SLW, MWH, AFZ, EJW, NJM, SEH

Investigation: RBL, SS, JSC, SAS, AB, AAM, CAH, PC, STF, TA, RT, AFF, JREB, SMH, JWL

Data Curation: RBL, STF, TA

Formal Analysis: RBL, SAS, AB, MK, HF, DMT, RF, NJM, SEH

Writing – Original Draft: RBL, NJM, SEH

Writing – Reviewing & Editing: all authors Visualization: RBL, AB

Funding Acquisition: RBL, JCF, EMB, DTT, EJW, NJM, SEH

Supervision: NJM, SEH

## Data and materials availability

Code and de-identified study data will be made available at https://github.com/Lindell-Lab/STAT-Hyperactivation-in-Sepsis at the time of publication. MINSEQE-compliant transcriptional data will also be made available in the GEO repository at the time of publication.

