## Supplemental Digital Content for "Aberrant STAT signaling drives T cell dysregulation in a targetable pediatric sepsis endotype"

**This PDF file includes:**

- Supplemental methods

- Fig. S1. Supporting evidence for *k*=3 clusters in Fig. 2A.
- Fig. S2. PCA resolving three subgroups based on expression of 24 elastic net derived severity-associated proteins.
- Fig. S3. Representative gating strategy for spectral flow immune phenotyping panel.
- Fig. S4. Results from single cell transcriptomics quality control and integration steps.

- Table S1. 1472 proteins measured in proteomics panel used in MODS and healthy control cohorts.
- Table S2. Antibodies used in 35-marker spectral flow immune phenotyping panel.
- Table S3. PCA loadings in Fig. 1D, using the full proteomics dataset.
- Table S4. Severity-associated proteins identified through linear mixed-effects model after adjustment for age, sex, and day from MODS onset.
- Table S5. Etiology of MODS and computed subgroup for each patient with MODS.
- Table S6. Immunocompromised diagnoses by subgroup.
- Table S7. 368 proteins measured in proteomics panel used in IEI cohort.
- Table S8. Leading edge analysis of protein expression in Group C patients compared to HC participants.
- Table S9. Antibodies used in 13-marker T cell phosphoflow cytometry panel.
- Table S10. Modified Proulx criteria used for screening and enrollment in the MODS cohort.
- Table S11. Age and sex for each healthy control participant.
- Table S12. Final set of 1448 proteins which met quality control thresholds in all three experiments and were included in downstream analyses.
- Table S13. Gene sets used in GSEA and GSVA analysis.
- Table S14. Gene sets used in Immune Dictionary cytokine signature analysis.

### SUPPLEMENTAL METHODS

#### PROTOCOL FOR THAWING AND STAINING HUMAN PBMCS FOR IMMUNE PROFILING ANALYSIS

<u>PBMC Thawing Protocol</u>

1. Prepare 15mL conical tubes with 10mL cRPMI (warmed 37°C water bath).
2. Obtain cryovials from LN and keep on dry ice until immediately before thaw.
3. Swirl 2 vials in 37°C water bath for ~30 seconds. Partially thawed pellet should remain.
4. Add 1mL warm cRPMI to cryovial and mix. Transfer all contents into 15mL tube.
5. Wash cryovial with 2mL warm cRPMI and transfer contents to 15mL tube.
6. Spin @ 800xg for 4 mins at RT.
7. Decant supernatant into waste and tap vial ridge to remove excess media.
8. Resuspend cells in 1mL warm cRPMI.
9. Count cells: 10μL Trypan blue + 10μL Cell suspension.
10. Spin @ 800xg for 4 mins at RT.
11. Decant supernatant into waste and tap vial ridge to remove excess media.
12. Resuspend cells in warm cRPMI at a concentration of $5x10^6$ cells/mL.
13. Plate 200μL/well, or $1x10^6$ cells/well, in a 96-well round bottom plate.

<u>PBMC Staining Protocol</u>

1. Spin at 800xg for 4 mins, RT. Flick plate to discard supernatant.
2. Wash cells with 150μL of PBS.
3. Spin at 800xg for 4 mins, RT. Flick plate to discard supernatant.
4. Resuspend cells in 50μL **LD/Fc stain mixture**.
5. Incubate at 4°C for <u>15 minutes</u>.
6. Wash cells with 150μL of PBS.
7. Spin at 800xg for 4 mins, RT. Flick plate to discard supernatant.
8. Resuspend cells in 50μL **SS antibody mixture** in FACS Buffer + Brilliant Stain Buffer.
9. Incubate at 4°C for <u>30 minutes</u> in the dark.
10. Wash with 150μL FACS buffer.
11. Spin @ 800xg for 4 mins, RT. Flick plate to discard supernatant.
12. Resuspend cells in 50μL of **Fix/Perm reagent**.
13. Incubate at RT for <u>20 minutes</u> in the dark.
14. Wash with 150μL Perm buffer.
15. Spin @ 800xg for 4 mins, RT. Flick plate to discard supernatant.
16. Resuspend cells in 50μL **ICS antibody mixture** in Perm Buffer + Brilliant Stain Buffer.
17. Incubate at 4°C for <u>60 minutes</u> in the dark.
18. Wash with 150μL of Perm Buffer.
19. Spin @ 800xg for 4 mins, RT. Flick plate to discard supernatant.
20. Resuspend pellets in 100μL **1.6% PFA fixative** and transfer to bullet tubes.
21. Cells can be held at 4°C overnight prior to acquisition.

#### Protocol for Thawing and Staining Human PBMCs for Aurora CS Sorting

PBMC Thawing Protocol

1. Prepare 15mL conical tubes with 10mL cRPMI (warmed 37°C water bath).
2. Obtain PBMC cryovials from LN and keep on dry ice until immediately before thaw.
3. Swirl 2 vials in 37°C water bath for ~30 seconds. Partially thawed pellet should remain.
4. Add 1mL warm cRPMI to cryovial and mix. Transfer all contents into 15mL tube.
5. Wash cryovial with 2mL warm cRPMI and transfer contents to 15mL tube.
6. Spin @ 800xg for 4 mins at RT.
7. Decant supernatant into waste and tap vial ridge to remove excess media.
8. Resuspend cells in 1mL warm cRPMI.
9. Count cells with 10µL trypan blue + 10µL cell suspension.
10. Lift $1x10^6$ live cells into a new labeled tube for staining.

PBMC Staining Protocol

1. Spin at 800xg for 4 mins, RT. Decant/tap to remove excess media.
2. Wash cells with 1mL of FACS buffer.
3. Spin at 800xg for 4 mins, RT. Decant/tap to remove excess media.
4. Resuspend cells in 100µL **Stain Mixture**.
5. Incubate at RT for 30 minutes in the dark.
6. Wash cells with 1mL of FACS buffer.
7. Spin at 800xg for 4 mins, RT. Decant/tap to remove excess media.
8. Resuspend cells in 500µL FACS Buffer and pass through blue cell strainer into FACS tube.
9. Cover and keep on wet ice. Cells are ready for sorting.

Prepare Sort Tubes and Resuspension Media

1. Prepare three 1.5ml eppendorf tubes with 500µL of PBS + 20% FBS for each sample.
2. Prepare 10% FBS Resuspension Media.

Cell Washing and Resuspension

1. After sorting, spin down cells in microcentrifuge 800xg for 5 mins, 4 degrees.
2. Aspirate supernatant and wash with 500µL PBS + 10% FBS.
3. Spin down cells in microcentrifuge 800xg for 5 mins, 4 degrees.
4. Aspirate supernatant and wash with 500µL PBS + 10% FBS.
5. Spin down cells in microcentrifuge 800xg for 5 mins, 4 degrees.
6. Goal concentration is 1200 cells per microliter. Calculate resuspension volume.
7. Aspirate supernatant and resuspend in PBS + 10% FBS.
8. Cover and keep on wet ice. Cells are ready for 10x processing.

**PROTOCOL FOR THAWING AND STAINING HUMAN PBMCS FOR PHOSPHOFLOW CYTOMETRY**

PBMC Stimulation Protocol

1. Prepare a 2µg/mL solution of anti-CD3 in sterile PBS.
   - InVivoMAb Anti-Human CD3, BioXCell, UCHT1 (Leu-4) (T3), BE0231
2. Add 200µL of diluted anti-CD3 solution to stimulation wells of a flat bottom 96 well plate.
3. Add 200µL of PBS to unstimulated wells in the same flat bottom 96 well plate.
4. Cover the plate with a plate sealer and store at 4°C overnight.

PBMC Thawing Protocol

5. Prepare 15mL conical tubes with 10mL cRPMI (warmed 37°C water bath).
6. Obtain PBMC cryovials from LN and keep on dry ice until immediately before thaw.
7. Swirl 2 vials in 37°C water bath for ~30 seconds. Partially thawed pellet should remain.
8. Add 1mL warm cRPMI to cryovial and mix. Transfer all contents into 15mL tube.
9. Wash cryovial with 2mL warm cRPMI and transfer contents to 15mL tube.
10. Spin @ 800xg for 4 mins at RT.
11. Decant supernatant into waste and tap vial ridge to remove excess media.
12. Resuspend cells in 1mL warm cRPMI.
13. Count cells with 10µL trypan blue + 10µL cell suspension.
14. Spin @ 800xg for 4 mins at RT.
15. Decant supernatant into waste and tap vial ridge to remove excess media.
16. Resuspend cells in warm cRPMI at a concentration of $2.5x10^6$ cells/mL.

PBMC Stimulation Protocol Continued

17. Remove anti-CD3 flat bottom plate from 4°C and carefully aspirate coating solution and PBS from wells.
18. Plate 100µL/well, or $2.5x10^5$ cells/well, in the 96-well flat bottom plate coated with anti-human CD3 antibody and PBS.
19. Prepare a 2µg/mL of an anti-human CD28 in AIM-V media. *Add cytokine if desired here.*
    - BD FastImmune Anti-Human CD28/CD49d, BD, L293/L25, 347690
20. Add 100µL of stimulation media to each stimulated well.
21. Add 100µL of AIM-V media to each unstimulated well.
22. Incubate at 37°C, 5% $CO_2$ for 24 hours.

PBMC Staining Protocol

23. Spin at 800xg for 4 mins, RT. Flick plate to discard supernatant.
24. Resuspend cells in 50µL of **pFlow surface stain mixture** and incubate at RT for 30 minutes.
25. Wash cells with 150µL of FACS Buffer.
26. Spin at 800xg for 4 minutes at RT. Flick plate to discard supernatant.
27. Resuspend cells in 100µL of **1.6% PFA** and incubate at RT for 15 minutes.
28. Spin at 800xg for 4 minutes at RT. Flick plate to discard supernatant.
29. Add 150µL/well of **cold MeOH**.
30. Wrap plate in foil and incubate at -20°C overnight.
31. Spin at 800xg for 5 minutes at RT. Flick plate to discard supernatant.
32. Wash cells with 200µL of Perm Buffer.

33. Spin at 800xg for 4 minutes at RT. Flick plate to discard supernatant.
34. Wash cells with 200µL of Perm Buffer.
35. Spin at 800xg for 4 minutes at RT. Flick plate to discard supernatant.
36. Resuspend cells in 50µL of **pFlow ICS stain mixture** and incubate at RT for <u>60 minutes</u>.
37. Wash cells with 150µL of Perm Buffer.
38. Spin at 800xg for 4 minutes at RT. Flick plate to discard supernatant.
39. Resuspend cells in 125µL of **1.6% PFA**.
40. Cover and keep on wet ice. Cells are ready for acquisition.

**Fig. S1. Supporting evidence for *k*=3 clusters in Fig. 2A.**

(A) This cumulative distribution function (CDF) plot demonstrates increased cluster stability up to *k*=3 (as indicated by flatter line), with decreasing cluster stability with finer clustering.

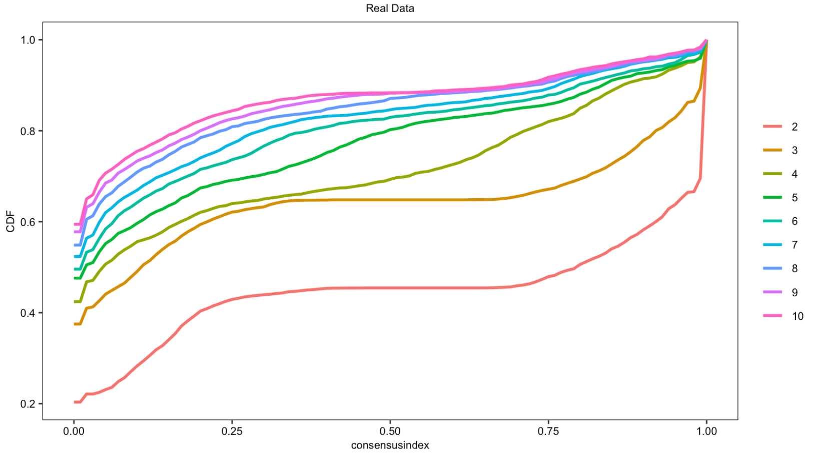


(B) Relative Cluster Stability Index (RCSI) is highest at *k*=3.

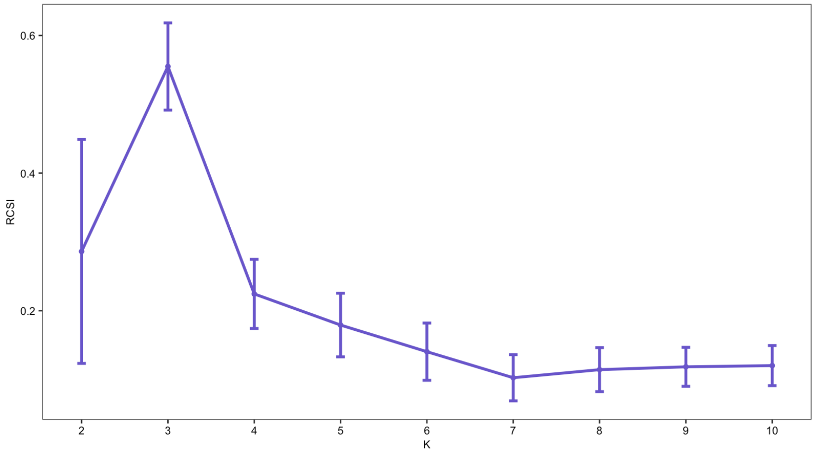


(C) M3C calculates empirical *p*-values from reference distributions to test the null hypothesis that *k*=1. In our analysis, at *k*=3 the null hypothesis is rejected (p=0.038).

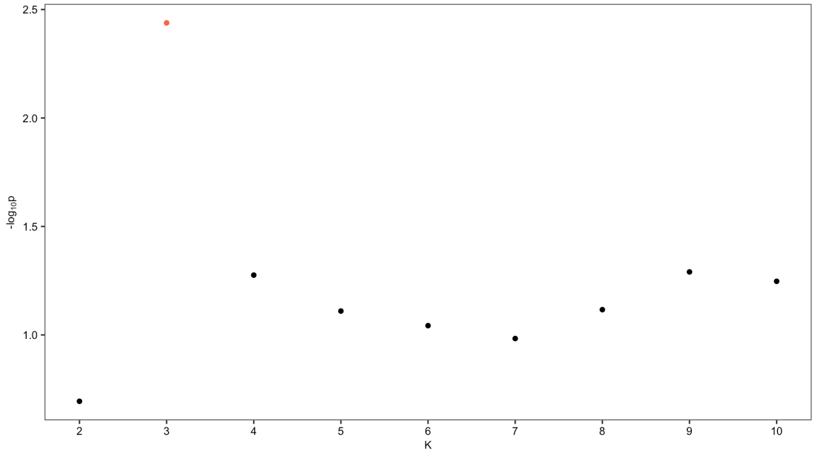

**Fig. S2. PCA resolving three subgroups based on expression of 24 elastic net derived severity-associated proteins.**

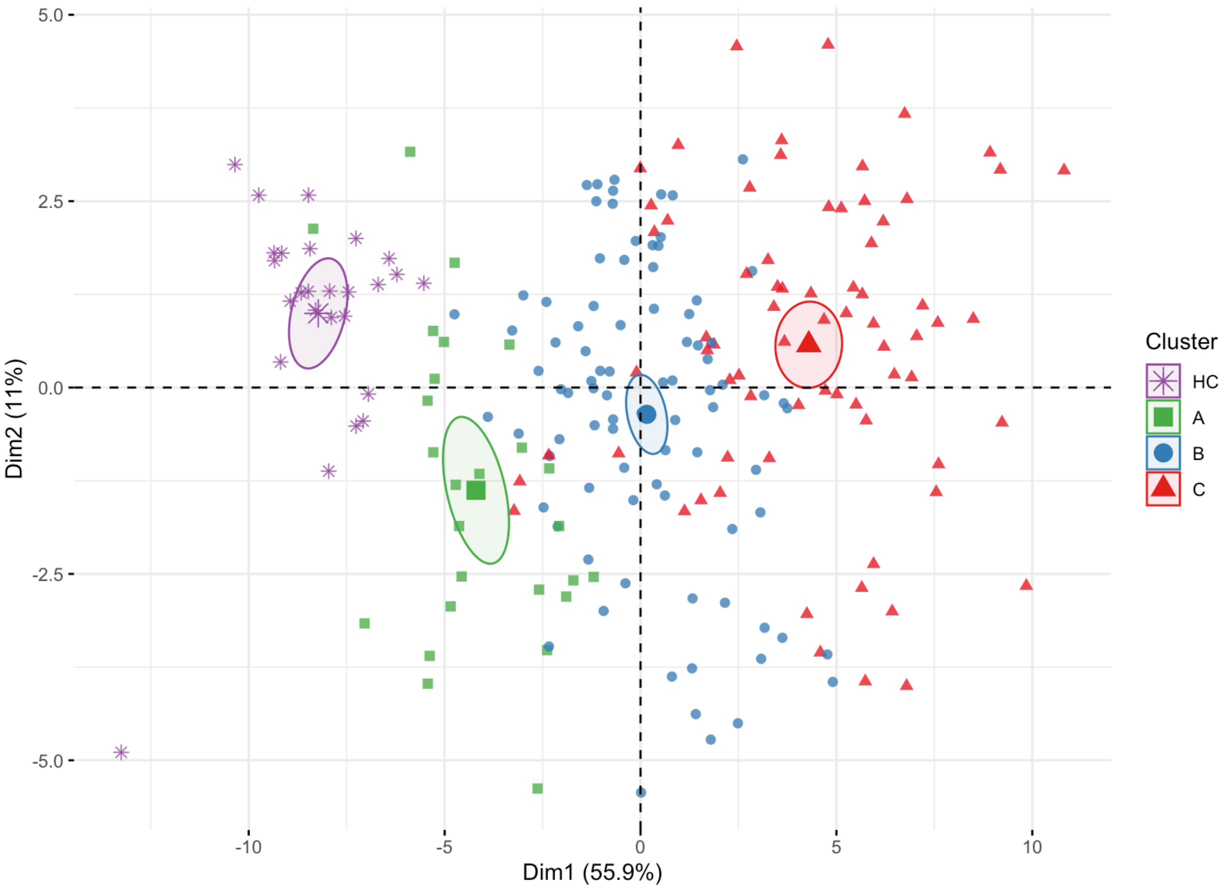


PCA dimensionality reduction demonstrates effective resolution of three MODS subgroups based on expression of 24 elastic net derived severity-associated proteins, with the majority of variance within the dataset defined by Dimension 1 as expected.

**Fig. S3. Representative gating strategy for spectral flow immune phenotyping panel.**

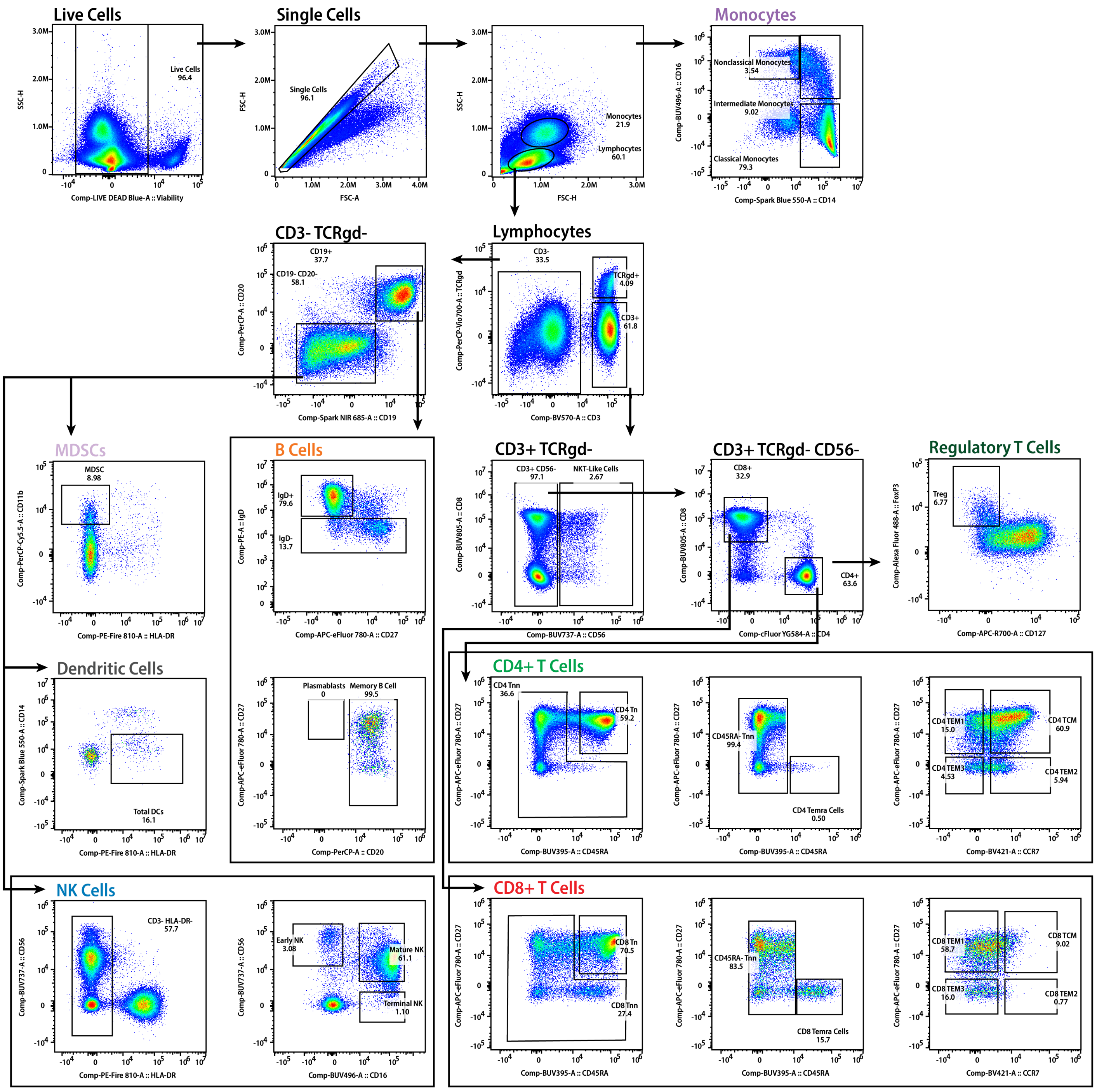

**Fig. S4. Results from single cell transcriptomics quality control and integration steps.**

**(A) Quality Control**

After review of standard quality control metrics, cells with less than 200 genes, greater than 3 median absolute deviation above the median, and >10% mitochondrial RNA content were removed from downstream analysis.

After applying these quality control steps, sample quality was consistent across groups.

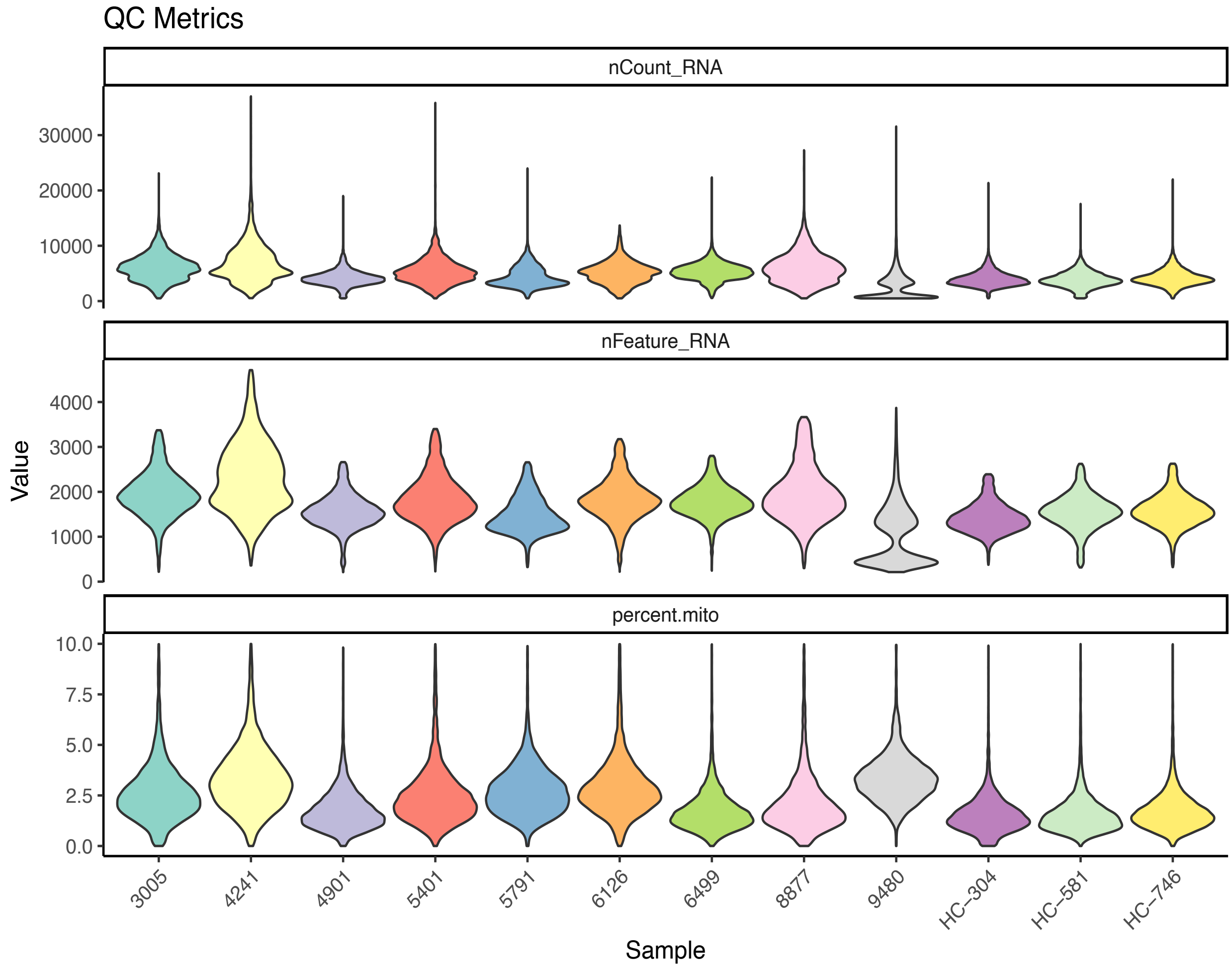

**(B) RPCA Integration**

Unintegrated data demonstrates differences between Group C Patients and HC Participants.

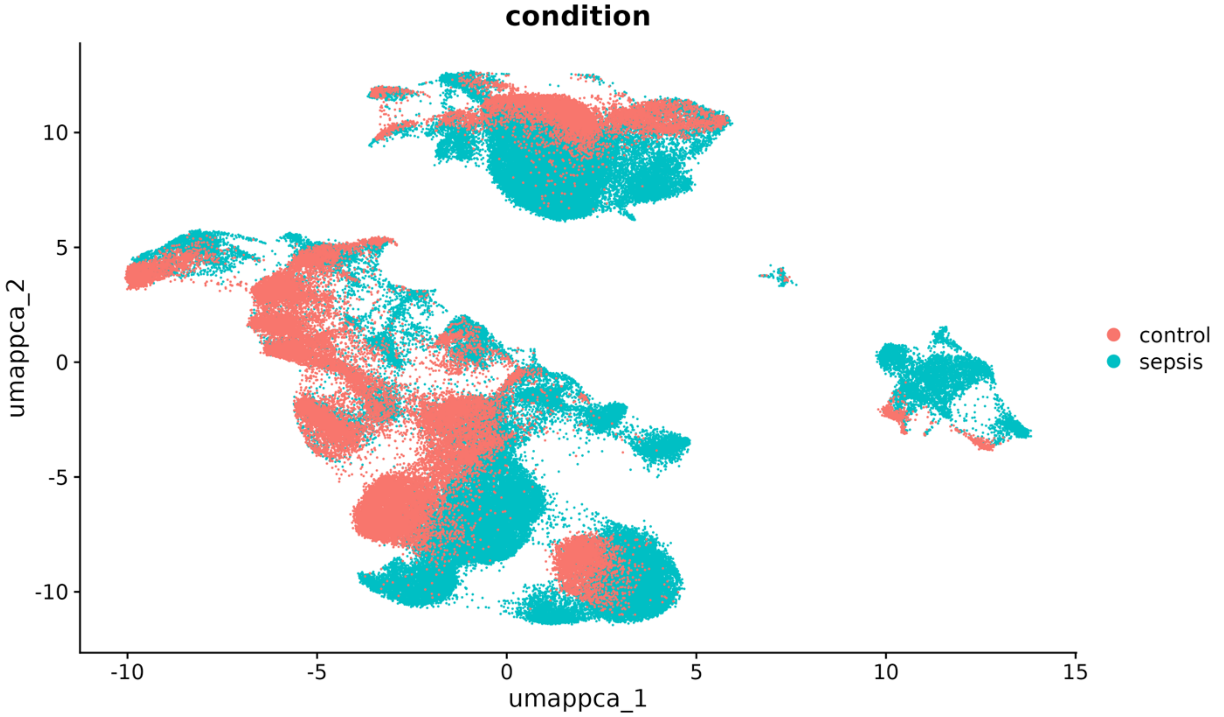


Integration with RPCA resolves discrepancies between Group C Patients and HC Participants.

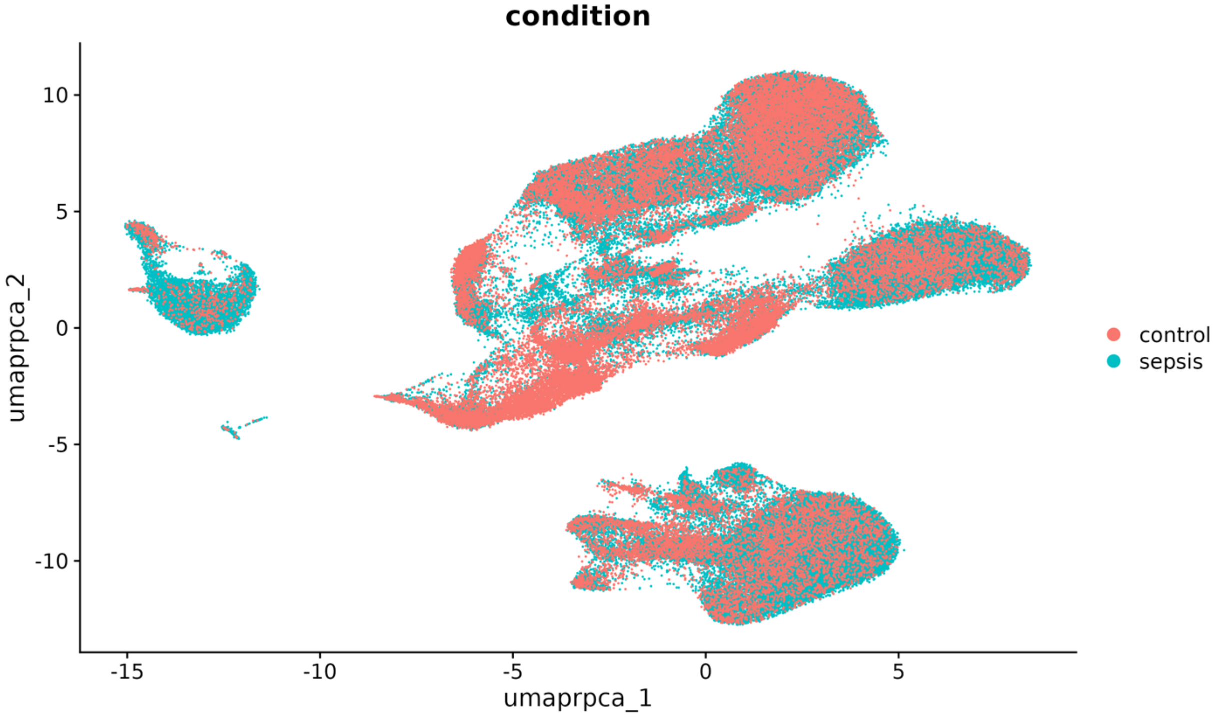

After integration, each RPCA cluster contains cells from MODS patients and HC participants.

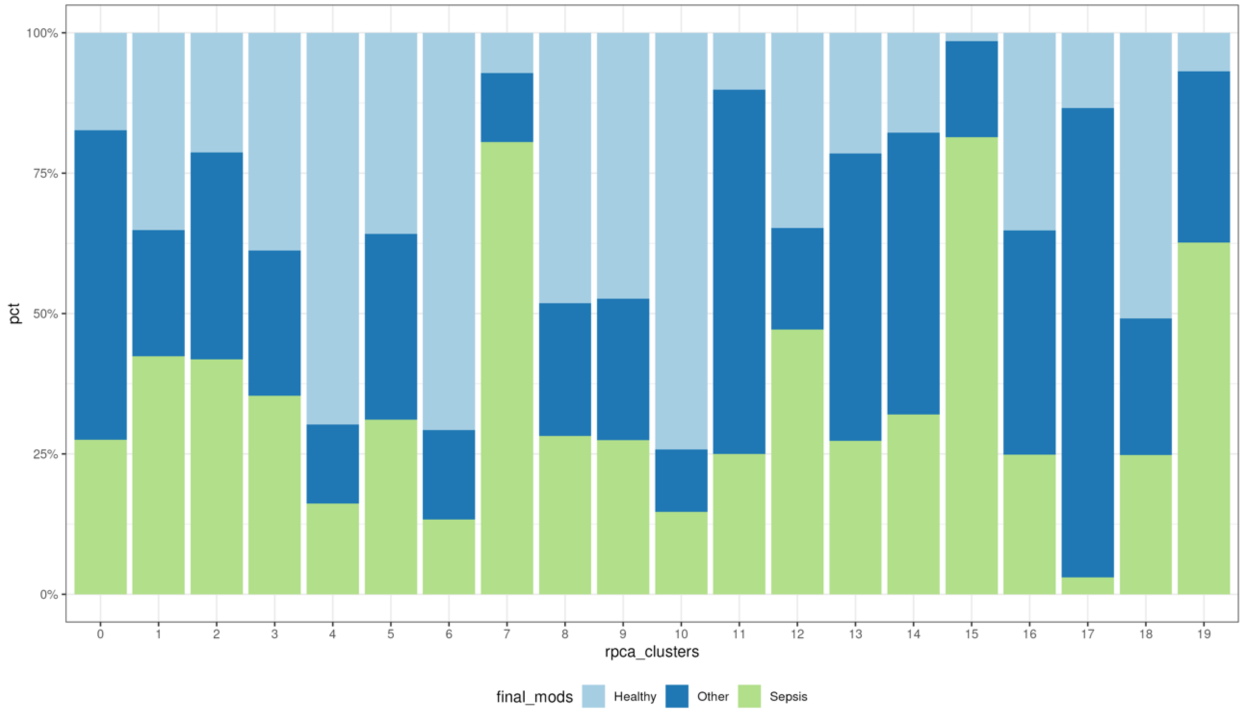

**Table S1. 1472 proteins measured in proteomics panel used in MODS and healthy control cohorts.**

*Due to its size, Table S1 is available in the attached supplemental spreadsheet.*

#### Table S2. Antibodies used in 35-marker spectral flow immune phenotyping panel.

| MARKER | FLUORORPHORE | VENDOR | CLONE | CATALOG NO. |
|---|---|---|---|---|
| CD45RA | BUV395 | BD Biosciences | H100 | 740298 |
| Live/Dead | LD Blue | Thermo Fisher | N/A | L34961 |
| CD16 | BUV496 | BD Biosciences | 3G8 | 612944 |
| CD26 | BUV563 | BD Biosciences | L272 | 749181 |
| CD39 | BUV661 | BD Biosciences | TU66 | 749967 |
| CD56 | BUV737 | BD Biosciences | NCAM16.2 | 612767 |
| CD8 | BUV805 | BD Biosciences | SK1 | 612889 |
| CCR7 | BV421 | BioLegend | G043H7 | 353208 |
| CD123 | Super Bright 436 | Thermo Fisher | 6H6 | 62-1239-42 |
| CD161 | eFluor 450 | Thermo Fisher | HP-3G10 | 48-1619-41 |
| ICOS | BV480 | BD Biosciences | DX29 | 746248 |
| Ki67 | BV510 | BD Biosciences | B56 | 563462 |
| CD3 | BV570 | BioLegend | UCHT1 | 300436 |
| CCR4 | BV605 | BioLegend | L291H4 | 359418 |
| CD95 | BV650 | BioLegend | DX2 | 305642 |
| CCR6 | BV711 | BioLegend | G034E3 | 353436 |
| CXCR5 | BV750 | BD Biosciences | RF8B2 | 747111 |
| PD-1 | BV785 | BioLegend | EH12.2H7 | 329930 |
| Foxp3 | Alexa Fluor 488 | BioLegend | 259D | 320212 |
| CD14 | Spark Blue 550 | BioLegend | 63D3 | 367148 |
| CD20 | PerCP | BioLegend | 2H7 | 302324 |
| CD11b | PerCP-Cy5.5 | BioLegend | ICRF44 | 301328 |
| TCRγδ | PerCP-eFluor 710 | Thermo Fisher | B1.1 | 46-9959-42 |
| IgD | PE | BioLegend | IA6-2 | 348204 |
| CD4 | cFluor YG584 | CYTEK | SK3 | R7-20041-100T |
| Tbet | PE-Cy5 | Thermo Fisher | 4B10 | 15-5825-80 |
| CD25 | PE-Alexa Fluor 700 | Thermo Fisher | CD25-3G10 | MHCD2524 |
| CXCR3 | PE-Cy7 | BioLegend | G025H7 | 353720 |
| HLA-DR | PE-Fire 810 | BioLegend | L243 | 307683 |
| CRTH2 | APC | BioLegend | BM16 | 350110 |
| CD11c | Alexa Fluor 647 | BioLegend | 3.9 | 301620 |
| CD19 | Spark NIR 685 | BioLegend | HIB19 | 302270 |
| CD127 | APC-R700 | BD Biosciences | HIL-7R-M21 | 565185 |
| CD27 | APC-eFluor780 | Thermo Fisher | O323 | 47-0279-42 |
| CD38 | APC/Fire 810 | BioLegend | HIT2 | 303550 |

#### Table S3. PCA loadings in Fig. 1D, using the full proteomics dataset.

| DIMENSION 1<br>(TOP 25 PROTEINS) | |
|---|---|
| **PROTEIN NAME** | **CONTRIBUTION** |
| POLR2F | 0.92969158 |
| MAD1L1 | 0.91356503 |
| TXLNA | 0.89474736 |
| ELOA | 0.86811405 |
| DPY30 | 0.86400025 |
| EGLN1 | 0.85809847 |
| ZBTB17 | 0.85543741 |
| APBB1IP | 0.85335406 |
| PXN | 0.85301337 |
| GPKOW | 0.85281899 |
| DFFA | 0.85090827 |
| OGFR | 0.84328415 |
| HDGF | 0.841057 |
| PLAUR | 0.84001585 |
| NUDC | 0.83925242 |
| INPP1 | 0.83863173 |
| GFER | 0.83484888 |
| CKAP4 | 0.8330877 |
| RAD23B | 0.82862877 |
| SRP14 | 0.82675401 |
| NMNAT1 | 0.82477831 |
| NBN | 0.82470521 |
| EZR | 0.82439488 |
| DCTPP1 | 0.82416125 |
| HEXIM1 | 0.82383476 |

| DIMENSION 2<br>(TOP 25 PROTEINS) | |
|---|---|
| **PROTEIN NAME** | **CONTRIBUTION** |
| ARHGEF12 | 0.92927293 |
| DBNL | 0.91717031 |
| CASP3 | 0.91624786 |
| PPP1R12A | 0.90434238 |
| SRC | 0.90335717 |
| CRKL | 0.90060822 |
| NT5C3A | 0.89589151 |
| PLXNA4 | 0.89309739 |
| MAP2K6 | 0.88974514 |
| CA13 | 0.88624933 |
| TBC1D23 | 0.88578306 |
| CLIP2 | 0.88413467 |
| TJAP1 | 0.88285037 |
| SKAP2 | 0.87773049 |
| AXIN1 | 0.87715071 |
| PDLIM7 | 0.87685547 |
| MANF | 0.87614294 |
| CIAPIN1 | 0.87457034 |
| MYO9B | 0.87420508 |
| SNAP29 | 0.87219403 |
| MAP3K5 | 0.87165308 |
| DNAJA2 | 0.86881494 |
| CORO1A | 0.86234331 |
| CRACR2A | 0.86216539 |
| SNAP23 | 0.85969907 |

**Table S4. Severity-associated proteins identified through linear mixed-effects model after adjustment for age, sex, and day from MODS onset.**

*Due to its size, Table S4 is available in the attached supplemental spreadsheet.*

#### Table S5. Etiology of MODS and computed subgroup for each MODS patient.

| HASHED PATIENT ID | MODS ETIOLOGY | CAUSE OF SEPSIS | SUBGROUP |
|---|---|---|---|
| 1018 | Cardiopulmonary Bypass | n/a | B |
| 1168 | Sepsis | Culture Negative Sepsis | B |
| 1234 | Sepsis | Culture Negative Sepsis | C |
| 1344 | Cardiopulmonary Bypass | n/a | B |
| 1426 | Sepsis | Culture Negative Sepsis | B |
| 1501 | Non-infected | n/a | B |
| 1521 | Sepsis | Bacterial Sepsis | B |
| 1523 | Non-infected | n/a | B |
| 1568 | Non-infected | n/a | B |
| 1620 | Non-infected | n/a | B |
| 1670 | Sepsis | Culture Negative Sepsis | B |
| 1677 | Non-infected | n/a | C |
| 1845 | Sepsis | Viral Sepsis | A |
| 1947 | Non-infected | n/a | B |
| 1973 | Non-infected | n/a | A |
| 2040 | Non-infected | n/a | C |
| 2056 | Cardiopulmonary Bypass | n/a | B |
| 2166 | Non-infected | n/a | A |
| 2243 | Cardiopulmonary Bypass | n/a | B |
| 2274 | Non-infected | n/a | C |
| 2309 | Cardiopulmonary Bypass | n/a | B |
| 2314 | Non-infected | n/a | C |
| 2393 | Sepsis | Culture Negative Sepsis | B |
| 2439 | Cardiopulmonary Bypass | n/a | B |
| 2726 | Non-infected | n/a | C |
| 2997 | Non-infected | n/a | A |
| 3005 | Sepsis | Bacterial Sepsis | C |
| 3134 | Sepsis | Bacterial Sepsis | C |
| 3227 | Sepsis | Viral Sepsis | C |
| 3365 | Sepsis | Culture Negative Sepsis | C |
| 3502 | Cardiopulmonary Bypass | n/a | B |
| 3596 | Non-infected | n/a | B |
| 3638 | Sepsis | Bacterial Sepsis | B |
| 3645 | Non-infected | n/a | B |
| 3783 | Non-infected | n/a | C |
| 3986 | Sepsis | Culture Negative Sepsis | B |
| 4130 | Sepsis | Viral Sepsis | B |
| 4241 | Sepsis | Culture Negative Sepsis | C |
| 4464 | Non-infected | n/a | B |
| 4638 | Non-infected | n/a | B |
| 4798 | Sepsis | Viral Sepsis | B |
| 4807 | Sepsis | Culture Negative Sepsis | B |
| 4868 | Sepsis | Viral Sepsis | B |
| 4901 | Sepsis | Culture Negative Sepsis | C |
| 5017 | Non-infected | n/a | A |
| 5368 | Sepsis | Culture Negative Sepsis | A |
| 5401 | Non-infected | n/a | C |
| 5522 | Sepsis | Culture Negative Sepsis | B |
| 5565 | Sepsis | Bacterial Sepsis | B |
| 5700 | Sepsis | Viral Sepsis | B |
| 5734 | Sepsis | Viral Sepsis | A |
| 5791 | Non-infected | n/a | C |
| 5842 | Sepsis | Culture Negative Sepsis | A |
| 5859 | Cardiopulmonary Bypass | n/a | C |
| 5909 | Non-infected | n/a | C |
| 5931 | Sepsis | Viral Sepsis | C |
| 6126 | Sepsis | Bacterial Sepsis | C |
| 6134 | Non-infected | n/a | A |
| 6468 | Sepsis | Culture Negative Sepsis | C |
| 6499 | Sepsis | Bacterial Sepsis | C |
| 6564 | Non-infected | n/a | A |

| HASHED PATIENT ID | MODS ETIOLOGY | CAUSE OF SEPSIS | SUBGROUP |
|---|---|---|---|
| 6606 | Sepsis | Viral Sepsis | B |
| 6698 | Non-infected | n/a | B |
| 7184 | Non-infected | n/a | A |
| 7331 | Sepsis | Culture Negative Sepsis | C |
| 7343 | Non-infected | n/a | C |
| 7411 | Non-infected | n/a | C |
| 7433 | Non-infected | n/a | B |
| 7632 | Non-infected | n/a | B |
| 7639 | Non-infected | n/a | B |
| 7791 | Trauma | n/a | A |
| 7810 | Non-infected | n/a | A |
| 7919 | Non-infected | n/a | A |
| 7977 | Trauma | n/a | A |
| 8047 | Non-infected | n/a | B |
| 8823 | Non-infected | n/a | C |
| 8825 | Sepsis | Bacterial Sepsis | B |
| 8877 | Non-infected | n/a | C |
| 8893 | Trauma | n/a | A |
| 9129 | Trauma | n/a | A |
| 9222 | Non-infected | n/a | A |
| 9328 | Non-infected | n/a | B |
| 9345 | Non-infected | n/a | A |
| 9480 | Sepsis | Culture Negative Sepsis | C |
| 9484 | Non-infected | n/a | B |
| 9630 | Sepsis | Culture Negative Sepsis | C |
| 9712 | Sepsis | Viral Sepsis | C |
| 9948 | Non-infected | n/a | C |

#### Table S6. Immunocompromised diagnoses by subgroup.

| HASHED PATIENT ID | IMMUNOCOMPROMISED DIAGNOSIS | COMPUTED SUBGROUP | SURVIVAL TO PICU DISCHARGE |
|---|---|---|---|
| 5368 | Cancer, solid tumor | A | Yes |
| 6564 | Cancer, leukemia | A | Yes |
| 1521 | Very early onset inflammatory bowel disease | B | Yes |
| 3596 | Hematopoietic cell transplant | B | Yes |
| 5522 | Cancer, solid tumor | B | No |
| 5565 | Cancer, solid tumor | B | Yes |
| 7433 | Cancer, solid tumor | B | No |
| 9484 | Cancer, leukemia | B | Yes |
| 3227 | Hematopoietic cell transplant | C | Yes |
| 6499 | Common variable immunodeficiency | C | No |
| 9630 | Cancer, solid tumor | C | Yes |

**Table S7. 368 proteins measured in proteomics panel used in IEI cohort.**

*Due to its size, Table S7 is available in the attached supplemental spreadsheet.*

**Table S8. Leading edge analysis of protein expression in Group C patients compared to HC participants.**

| HALLMARK PATHWAY | FDR ADJUSTED P-VALUE | LEADING EDGE ENRICHMENT |
|---|---|---|
| HALLMARK_IL6_JAK_ STAT3_SIGNALING | 0.004779285 | IL6, CCL7, PLA2G2A, REG1A, JUN, HMOX1, IL4R, TNFRSF12A, TNFRSF1A, TNFRSF1B, CXCL10, CXCL9, IL2RA, IL15RA, IL18R1, LTBR, CSF1, CD38, CXCL11, ACVRL1, IL1B, CD14, TNF, CXCL13, IL12RB1, TGFB1, CSF2RA, FAS, IFNGR1 |
| HALLMARK_XENOBIOTIC_ METABOLISM | 0.005617107 | PTS, FABP1, IGFBP1, FBP1, REG1A, HMOX1, TNFRSF1A, NINJ1, AHCY, CES1, ARG1, DDC, IGFBP4, EPHA2, GSTA3, DCXR |
| HALLMARK_ INFLAMMATORY_ RESPONSE | 0.008012074 | IL6, CCL7, CXCL8, CSF3, CCL20, LIF, TIMP1, IL10, BST2, IL4R, OSM, TNFRSF1B, CXCL10, NAMPT, CXCL9, CCL2, IL15, PLAUR, CLEC5A, LDLR, IL15RA, PVR, IL18R1, ADM, LAMP3, CSF1, MSR1, LYN, CXCL11, IL1B, CX3CL1, SLAMF1, CD14 |
| HALLMARK_INTERFERON_ ALPHA_RESPONSE | 0.016318021 | BST2, IL4R, GBP2, CXCL10, IL15, CASP1, LAMP3, CSF1, CD74, OGFR, PSME2, LAP3, CXCL11 |
| HALLMARK_TNFA_ SIGNALING_VIA_NFKB | 0.030735353 | IL6, AREG, CCL20, LIF, SERPINB8, JUN, CXCL10, NAMPT, FOSB, NINJ1, CCL2, PLAUR, CEBPB, LDLR, IL15RA, TNC, SOD2, CSF1, CCN1, B4GALT1, PTX3, CXCL11, KYNU, IL1B, CCL4, TNF, NFKBIE, VEGFA, TANK, F3, IL18, ICAM1 |
| HALLMARK_INTERFERON_ GAMMA_RESPONSE | 0.031250000 | IL6, CCL7, BST2, IL4R, CXCL10, NAMPT, CXCL9, CCL2, IL15, CASP1, IL15RA, CD274, SOD2, IL18BP, CD38, CD74, OGFR, PSME2, VAMP5, LAP3, CXCL11 |
| HALLMARK_OXIDATIVE_ PHOSPHORYLATION | 0.035126235 | NDUFS6, GRPEL1, ACAA1, POLR2F, BAX, COX5B |
| HALLMARK_UNFOLDED_ PROTEIN_RESPONSE | 0.035144814 | IGFBP1, ZBTB17, EIF4EBP1, CCL2, CEBPB, NPM1, FUS, PREB, BAG3, VEGFA |
| HALLMARK_EPITHELIAL_ MESENCHYMAL_ TRANSITION | 0.038038038 | IL6, AREG, CXCL8, SDC1, PRSS2, TIMP1, JUN, FSTL3, TNFRSF12A, TFPI2, SFRP1, TNFRSF11B, THBS2, MMP3, IL15, ACTA2, TGM2, PLAUR, COL4A1, TNC, BGN, PVR, CAPG, IGFBP4, VCAN, IGFBP2 |
| HALLMARK_ESTROGEN_ RESPONSE_LATE | 0.044102564 | AREG, KRT19, AGR2, TFPI2, SORD, FKBP4, MDK, TFF1, IGFBP4, CA12, DCXR, SLC16A1, TFF3, BLVRB, MAPT, FABP5, FKBP5 |

#### Table S9. Antibodies used in 13-marker T cell phosphoflow cytometry panel.

| MARKER | FLUORORPHORE | VENDOR | CLONE | CATALOG NO. |
|---|---|---|---|---|
| CD4 | BUV395 | BD | RPA-T4 | 564724 |
| Live/Dead Aqua | V500 | Thermo Scientific | N/A | L34966 |
| CD14 | V500 | BD | M5E2 | 561391 |
| CD16 | V500 | BD | 3G8 | 561394 |
| CD19 | V500 | BD | HIB19 | 561121 |
| CD8a | BV605 | BioLegend | RPA-T8 | 301040 |
| CD45RA | BV650 | BD | HI100 | 563963 |
| CD27 | BV785 | BioLegend | O323 | 302832 |
| pSTAT1 | AF488 | BD | 4A | 612596 |
| pSTAT3 | PE | BD | 4/P STAT3 | 612569 |
| pSTAT5 | PE-Cy7 | BD | 47 | 560117 |
| Total STAT3 | APC | BD | M59-50 | 560392 |
| CD3 | APC-R700 | BD | UCHT1 | 565119 |

**Table S10. Modified Proulx criteria used for screening and enrollment in the MODS cohort.**

| ORGAN SYSTEM | CLINICAL AND LABORATORY CRITERIA FOR ORGAN DYSFUNCTION[1] |
|---|---|
| Respiratory Dysfunction | i) Respiratory rate > 90 breaths per min (age < 1yr) or > 70 breaths per min (age ≥ 1yr) **OR**<br>ii) PaO2 < 40 mmHg (5.3kPa) in absence of cyanotic congenital heart disease **OR**<br>iii) PaCO2 ≥ 65 mmHg (8.7kPa) **OR**<br>iv) PaO2/FiO2 < 200 in absence of cyanotic congenital heart disease (where P/F ratio is based on simultaneous measurements) **OR**<br>v) Mechanical ventilation<br>(1) Must be for > 24hrs post-operative if intubated just for surgery/procedure<br>(2) Includes new HFNC ≥ 6L/min, non-invasive ventilation (e.g. CPAP, BiPAP), and VV ECMO<br>(3) Includes patients with tracheostomy who do NOT require mechanical ventilation at baseline<br>(4) For subjects with non-invasive ventilation at baseline, intubation would qualify as Respiratory Dysfunction |
| Cardiovascular Dysfunction | i) Systolic blood pressure < 40 mmHg (age < 1yr) or < 50 mmHg (age ≥ 1yr) **OR**<br>ii) Heart rate < 50 or > 220 (age < 1yr), < 40 or > 200 (age ≥1yr) **OR**<br>iii) Cardiac arrest **OR**<br>iv) pH < 7.2 with normal PaCO2 **OR**<br>v) Continuous vasoactive drug infusion for hemodynamic support (excluding dopamine infusion ≤5ug/kg/min) **OR**<br>vi) VA ECMO **OR**<br>vii) Ventricular assist device |
| Hematologic Dysfunction | i) Hemoglobin < 5g/dL (50g/L) **OR**<br>ii) White blood cell count < 3,000/mm3 (3x10*9/L) **OR**<br>iii) Platelet count < 20,000/mm3 (20x10*9) **OR**<br>iv) PT > 20 seconds OR aPTT > 60 seconds (in absence of anticoagulation therapy) |
| Gastrointestinal Dysfunction | i) Gastrointestinal bleeding **AND** one of the following believed to be the result of gastroduodenal bleeding:<br>(1) Drop in hemoglobin ≥ 2g/dL (20g/L) over 24 hours **OR**<br>(2) Blood transfusion **OR**<br>(3) Hypotension with blood pressure < 5th percentile of age **OR**<br>(4) Gastric or duodenal surgery |
| Renal Dysfunction | i) Serum BUN ≥ 100mg/dL (36 mmol/L) **OR**<br>ii) Serum creatinine ≥ 2 mg/dL (177 umol/L) without pre-existing renal disease **OR**<br>iii) Dialysis and/or hemofiltration |
| Neurological Dysfunction | i) Fixed, dilated pupils **OR**<br>ii) Glasgow coma score (GCS) < 5 in the absence of neuromuscular blocking drugs |
| Hepatic Dysfunction | i) Total bilirubin > 3mg/dL (60 umol/L) |

- [1] MODS criteria derived from: F. Proulx *et al.*, The pediatric multiple organ dysfunction syndrome. *Pediatr Crit Care Med* **10**, 12-22 (2009).

#### Table S11. Age range and sex for each healthy control participant.

| HASHED PARTICIPANT ID | AGE RANGE (YEARS) | SEX |
|---|---|---|
| HC-105 | 6-10 | M |
| HC-175 | 11-15 | M |
| HC-182 | 0-5 | F |
| HC-304 | 6-10 | M |
| HC-335 | 11-15 | M |
| HC-411 | 11-15 | M |
| HC-417 | 16-20 | M |
| HC-439 | 11-15 | M |
| HC-500 | 6-10 | M |
| HC-534 | 11-15 | F |
| HC-549 | 0-5 | F |
| HC-581 | 11-15 | F |
| HC-598 | 11-15 | F |
| HC-665 | 16-20 | M |
| HC-679 | 6-10 | F |
| HC-698 | 6-10 | F |
| HC-730 | 6-10 | M |
| HC-746 | 6-10 | F |
| HC-778 | 6-10 | M |
| HC-782 | 0-5 | F |
| HC-841 | 0-5 | F |
| HC-919 | 16-20 | M |
| HC-937 | 0-5 | M |
| HC-963 | 11-15 | F |
| HC-979 | 6-10 | M |

**Table S12. Final set of 1448 proteins which met quality control thresholds in all three experiments and were included in downstream analyses.**

*Due to its size, Table S12 is available in the attached supplemental spreadsheet.*

#### Table S13. Gene sets used in GSEA and GSVA analysis.

| GENE SET | GENE SYMBOLS |
|---|---|
| HALLMARK_ TNFA_ SIGNALING_ VIA_NFKB | ABCA1, ACKR3, AREG, ATF3, ATP2B1, B4GALT1, B4GALT5, BCL2A1, BCL3, BCL6, BHLHE40, BIRC2, BIRC3, BMP2, BTG1, BTG2, BTG3, CCL2, CCL20, CCL4, CCL5, CCN1, CCND1, CCNL1, CCRL2, CD44, CD69, CD80, CD83, CDKN1A, CEBPB, CEBPD, CFLAR, CLCF1, CSF1, CSF2, CXCL1, CXCL10, CXCL11, CXCL2, CXCL3, CXCL6, DENND5A, DNAJB4, DRAM1, DUSP1, DUSP2, DUSP4, DUSP5, EDN1, EFNA1, EGR1, EGR2, EGR3, EHD1, EIF1, ETS2, F2RL1, F3, FJX1, FOS, FOSB, FOSL1, FOSL2, FUT4, G0S2, GADD45A, GADD45B, GCH1, GEM, GFPT2, GPR183, HBEGF, HES1, ICAM1, ICOSLG, ID2, IER2, IER3, IER5, IFIH1, IFIT2, IFNGR2, IL12B, IL15RA, IL18, IL1A, IL1B, IL23A, IL6, IL6ST, IL7R, INHBA, IRF1, IRS2, JAG1, JUN, JUNB, KDM6B, KLF10, KLF2, KLF4, KLF6, KLF9, KYNU, LAMB3, LDLR, LIF, LITAF, MAFF, MAP2K3, MAP3K8, MARCKS, MCL1, MSC, MXD1, MYC, NAMPT, NFAT5, NFE2L2, NFIL3, NFKB1, NFKB2, NFKBIA, NFKBIE, NINJ1, NR4A1, NR4A2, NR4A3, OLR1, PANX1, PDE4B, PDLIM5, PER1, PFKFB3, PHLDA1, PHLDA2, PLAU, PLAUR, PLEK, PLK2, PLPP3, PMEPA1, PNRC1, PPP1R15A, PTGER4, PTGS2, PTPRE, PTX3, RCAN1, REL, RELA, RELB, RHOB, RIGI, RIPK2, RNF19B, SAT1, SDC4, SERPINB2, SERPINB8, SERPINE1, SGK1, SIK1, SLC16A6, SLC2A3, SLC2A6, SMAD3, SNN, SOCS3, SOD2, SPHK1, SPSB1, SQSTM1, STAT5A, TANK, TAP1, TGIF1, TIPARP, TLR2, TNC, TNF, TNFAIP2, TNFAIP3, TNFAIP6, TNFAIP8, TNFRSF9, TNFSF9, TNIP1, TNIP2, TRAF1, TRIB1, TRIP10, TSC22D1, TUBB2A, VEGFA, YRDC, ZBTB10, ZC3H12A, ZFP36 |
| HALLMARK_ IL6_ JAK_STAT3_ SIGNALING | A2M, ACVR1B, ACVRL1, BAK1, CBL, CCL7, CCR1, CD14, CD36, CD38, CD44, CD9, CNTFR, CRLF2, CSF1, CSF2, CSF2RA, CSF2RB, CSF3R, CXCL1, CXCL10, CXCL11, CXCL13, CXCL3, CXCL9, DNTT, EBI3, FAS, GRB2, HAX1, HMOX1, IFNAR1, IFNGR1, IFNGR2, IL10RB, IL12RB1, IL13RA1, IL15RA, IL17RA, IL17RB, IL18R1, IL1B, IL1R1, IL1R2, IL2RA, IL2RG, IL3RA, IL4R, IL6, IL6ST, IL7, IL9R, INHBE, IRF1, IRF9, ITGA4, ITGB3, JUN, LEPR, LTB, LTBR, MAP3K8, MYD88, OSMR, PDGFC, PF4, PIK3R5, PIM1, PLA2G2A, PTPN1, PTPN11, PTPN2, REG1A, SOCS1, SOCS3, STAM2, STAT1, STAT2, STAT3, TGFB1, TLR2, TNF, TNFRSF12A, TNFRSF1A, TNFRSF1B, TNFRSF21, TYK2 |
| HALLMARK_ IL2_STAT5_ SIGNALING | ABCB1, ADAM19, AGER, AHCY, AHNAK, AHR, ALCAM, AMACR, ANXA4, APLP1, ARL4A, BATF, BATF3, BCL2, BCL2L1, BHLHE40, BMP2, BMPR2, CA2, CAPG, CAPN3, CASP3, CCND2, CCND3, CCNE1, CCR4, CD44, CD48, CD79B, CD81, CD83, CD86, CDC42SE2, CDC6, CDCP1, CDKN1C, CISH, CKAP4, COCH, COL6A1, CSF1, CSF2, CST7, CTLA4, CTSZ, CXCL10, CYFIP1, DCPS, DENND5A, DHRS3, DRC1, ECM1, EEF1AKMT1, EMP1, ENO3, ENPP1, EOMES, ETFBKMT, ETV4, F2RL2, FAH, FGL2, FLT3LG, FURIN, GABARAPL1, GADD45B, GALM, GATA1, GBP4, GLIPR2, GPR65, GPR83, GPX4, GSTO1, GUCY1B1, HIPK2, HK2, HOPX, HUWE1, HYCC2, ICOS, IFITM3, IFNGR1, IGF1R, IGF2R, IKZF2, IKZF4, IL10, IL10RA, IL13, IL18R1, IL1R2, IL1RL1, IL2RA, IL2RB, IL3RA, IL4R, IRF4, IRF6, IRF8, ITGA6, ITGAE, ITGAV, ITIH5, KLF6, LCLAT1, LIF, LRIG1, LRRC8C, LTB, MAFF, MAP3K8, MAP6, MAPKAPK2, MUC1, MXD1, MYC, MYO1C, MYO1E, NCOA3, NCS1, NDRG1, NFIL3, NFKBIZ, NOP2, NRP1, NT5E, ODC1, P2RX4, P4HA1, PDCD2L, PENK, PHLDA1, PHTF2, PIM1, PLAGL1, PLEC, PLIN2, PLPP1, PLSCR1, PNP, POU2F1, PRAF2, PRKCH, PRNP, PTCH1, PTGER2, PTH1R, PTRH2, PUS1, RABGAP1L, RGS16, RHOB, RHOH, RNH1, RORA, RRAGD, S100A1, SCN9A, SELL, SELP, SERPINB6, SERPINC1, SH3BGRL2, SHE, SLC1A5, SLC29A2, SLC2A3, SLC39A8, SMPDL3A, SNX14, SNX9, SOCS1, SOCS2, SPP1, SPRED2, SPRY4, ST3GAL4, SWAP70, SYNGR2, SYT11, TGM2, TIAM1, TLR7, TNFRSF18, TNFRSF1B, TNFRSF21, TNFRSF4, TNFRSF8, TNFRSF9, TNFSF10, TNFSF11, TRAF1, TTC39B, TWSG1, UCK2, UMPS, WLS, XBP1 |
| HALLMARK_ INTERFERON_ GAMMA_ RESPONSE | ADAR, APOL6, ARID5B, ARL4A, AUTS2, B2M, BANK1, BATF2, BPGM, BST2, BTG1, C1R, C1S, CASP1, CASP3, CASP4, CASP7, CASP8, CCL2, CCL5, CCL7, CD274, CD38, CD40, CD69, CD74, CD86, CDKN1A, CFB, CFH, CIITA, CMKLR1, CMPK2, CMTR1, CSF2RB, CXCL10, CXCL11, CXCL9, DDX60, DHX58, EIF2AK2, EIF4E3, EPSTI1, FAS, FCGR1A, FGL2, FPR1, GBP4, GBP6, GCH1, GPR18, GZMA, HELZ2, HERC6, HIF1A, HLA-A, HLA-B, HLA-DMA, HLA-DQA1, HLA-DRB1, HLA-G, ICAM1, IDO1, IFI27, IFI30, IFI35, IFI44, IFI44L, IFIH1, IFIT1, IFIT2, IFIT3, IFITM2, IFITM3, IFNAR2, IL10RA, IL15, IL15RA, IL18BP, IL2RB, IL4R, IL6, IL7, IRF1, IRF2, IRF4, IRF5, IRF7, IRF8, IRF9, ISG15, ISG20, ISOC1, ITGB7, JAK2, KLRK1, LAP3, LATS2, LCP2, LGALS3BP, LY6E, LYSMD2, MARCHF1, METTL7B, MT2A, MTHFD2, MVP, MX1, MX2, MYD88, NAMPT, NCOA3, NFKB1, NFKBIA, NLRC5, NMI, NOD1, NUP93, OAS2, OAS3, OASL, OGFR, P2RY14, PARP12, PARP14, PDE4B, PELI1, PFKP, PIM1, PLA2G4A, PLSCR1, PML, PNP, PNPT1, PSMA2, PSMA3, PSMB10, PSMB2, PSMB8, PSMB9, PSME1, PSME2, PTGS2, PTPN1, PTPN2, PTPN6, RAPGEF6, RBCK1, RIGI, RIPK1, RIPK2, RNF213, RNF31, RSAD2, RTP4, SAMD9L, SAMHD1, SECTM1, SELP, SERPING1, SLAMF7, SLC25A28, SOCS1, SOCS3, SOD2, SP110, SPPL2A, SRI, SSPN, ST3GAL5, ST8SIA4, STAT1, STAT2, STAT3, STAT4, TAP1, TAPBP, TDRD7, TNFAIP2, TNFAIP3, TNFAIP6, TNFSF10, TOR1B, TRAFD1, TRIM14, TRIM21, TRIM25, TRIM26, TXNIP, UBE2L6, UPP1, USP18, VAMP5, VAMP8, VCAM1, WARS1, XAF1, XCL1, ZBP1, ZNFX1 |
| HALLMARK_ PI3K_AKT_ MTOR_ SIGNALING | ACACA, ACTR2, ACTR3, ADCY2, AKT1, AKT1S1, AP2M1, ARF1, ARHGDIA, ARPC3, ATF1, CAB39, CAB39L, CALR, CAMK4, CDK1, CDK2, CDK4, CDKN1A, CDKN1B, CFL1, CLTC, CSNK2B, CXCR4, DAPP1, DDIT3, DUSP3, E2F1, ECSIT, EGFR, EIF4E, FASLG, FGF17, FGF22, FGF6, GNA14, GNGT1, GRB2, GRK2, GSK3B, HRAS, HSP90B1, IL2RG, IL4, IRAK4, ITPR2, LCK, MAP2K3, MAP2K6, MAP3K7, MAPK1, MAPK10, MAPK8, MAPK9, MAPKAP1, MKNK1, MKNK2, MYD88, NCK1, NFKBIB, NGF, NOD1, PAK4, PDK1, PFN1, PIK3R3, PIKFYVE, PIN1, PITX2, PLA2G12A, PLCB1, PLCG1, PPP1CA, PPP2R1B, PRKAA2, PRKAG1, PRKAR2A, PRKCB, PTEN, PTPN11, RAC1, RAF1, RALB, RIPK1, RIT1, RPS6KA1, RPS6KA3, RPTOR, SFN, SLA, SLC2A1, |

| GENE SET | GENE SYMBOLS |
|---|---|
| | SMAD2, SQSTM1, STAT2, TBK1, THEM4, TIAM1, TNFRSF1A, TRAF2, TRIB3, TSC2, UBE2D3, UBE2N, VAV3, YWHAB |
| HALLMARK_ OXIDATIVE_ PHOSPHOR YLATION | ABCB7, ACAA1, ACAA2, ACADM, ACADSB, ACADVL, ACAT1, ACO2, AFG3L2, AIFM1, ALAS1, ALDH6A1, ATP1B1, ATP5F1A, ATP5F1B, ATP5F1C, ATP5F1D, ATP5F1E, ATP5MC1, ATP5MC2, ATP5MC3, ATP5ME, ATP5MF, ATP5MG, ATP5PB, ATP5PD, ATP5PF, ATP5PO, ATP6AP1, ATP6V0B, ATP6V0C, ATP6V0E1, ATP6V1C1, ATP6V1D, ATP6V1E1, ATP6V1F, ATP6V1G1, ATP6V1H, BAX, BCKDHA, BDH2, CASP7, COX10, COX11, COX15, COX17, COX4I1, COX5A, COX5B, COX6A1, COX6B1, COX6C, COX7A2, COX7A2L, COX7B, COX7C, COX8A, CPT1A, CS, CYB5A, CYB5R3, CYC1, CYCS, DECR1, DLAT, DLD, DLST, ECH1, ECHS1, ECI1, ETFA, ETFB, ETFDH, FDX1, FH, FXN, GLUD1, GOT2, GPI, GPX4, GRPEL1, HADHA, HADHB, HCCS, HSD17B10, HSPA9, HTRA2, IDH1, IDH2, IDH3A, IDH3B, IDH3G, IMMT, ISCA1, ISCU, LDHA, LDHB, LRPPRC, MAOB, MDH1, MDH2, MFN2, MGST3, MPC1, MRPL11, MRPL15, MRPL34, MRPL35, MRPS11, MRPS12, MRPS15, MRPS22, MRPS30, MTRF1, MTRR, MTX2, NDUFA1, NDUFA2, NDUFA3, NDUFA4, NDUFA5, NDUFA6, NDUFA7, NDUFA8, NDUFA9, NDUFAB1, NDUFB1, NDUFB2, NDUFB3, NDUFB4, NDUFB5, NDUFB6, NDUFB7, NDUFB8, NDUFC1, NDUFC2, NDUFS1, NDUFS2, NDUFS3, NDUFS4, NDUFS6, NDUFS7, NDUFS8, NDUFV1, NDUFV2, NNT, NQO2, OAT, OGDH, OPA1, OXA1L, PDHA1, PDHB, PDHX, PDK4, PDP1, PHB2, PHYH, PMPCA, POLR2F, POR, PRDX3, RETSAT, RHOT1, RHOT2, SDHA, SDHB, SDHC, SDHD, SLC25A11, SLC25A12, SLC25A20, SLC25A3, SLC25A4, SLC25A5, SLC25A6, SUCLA2, SUCLG1, SUPV3L1, SURF1, TCIRG1, TIMM10, TIMM13, TIMM17A, TIMM50, TIMM8B, TIMM9, TOMM22, TOMM70, UQCR10, UQCR11, UQCRB, UQCRC1, UQCRC2, UQCRFS1, UQCRH, UQCRQ, VDAC1, VDAC2, VDAC3 |
| HALLMARK_ GLYCOLYSIS | ABCB6, ADORA2B, AGL, AGRN, AK3, AK4, AKR1A1, ALDH7A1, ALDH9A1, ALDOA, ALDOB, ALG1, ANG, ANGPTL4, ANKZF1, ARPP19, ARTN, AURKA, B3GALT6, B3GAT1, B3GAT3, B3GNT3, B4GALT1, B4GALT2, B4GALT4, B4GALT7, BIK, BPNT1, CACNA1H, CAPN5, CASP6, CD44, CDK1, CENPA, CHPF, CHPF2, CHST1, CHST12, CHST2, CHST4, CHST6, CITED2, CLDN3, CLDN9, CLN6, COG2, COL5A1, COPB2, CTH, CXCR4, CYB5A, DCN, DDIT4, DEPDC1, DLD, DPYSL4, DSC2, ECD, EFNA3, EGFR, EGLN3, ELF3, ENO1, ENO2, ERO1A, EXT1, EXT2, FAM162A, FBP2, FKBP4, FUT8, G6PD, GAL3ST1, GALE, GALK1, GALK2, GAPDHS, GCLC, GFPT1, GFUS, GLCE, GLRX, GMPPA, GMPPB, GNE, GNPDA1, GOT1, GOT2, GPC1, GPC3, GPC4, GPR87, GUSB, GYS1, GYS2, HAX1, HDLBP, HK2, HMMR, HOMER1, HS2ST1, HS6ST2, HSPA5, IDH1, IDUA, IER3, IGFBP3, IL13RA1, IRS2, ISG20, KDELR3, KIF20A, KIF2A, LCT, LDHA, LDHC, LHPP, LHX9, MDH1, MDH2, ME1, ME2, MED24, MERTK, MET, MIF, MIOX, MPI, MXI1, NANP, NASP, NDST3, NDUFV3, NOL3, NSDHL, NT5E, P4HA1, P4HA2, PAM, PAXIP1, PC, PDK3, PFKFB1, PFKP, PGAM1, PGAM2, PGK1, PGLS, PGM2, PHKA2, PKM, PKP2, PLOD1, PLOD2, PMM2, POLR3K, PPFIA4, PPIA, PPP2CB, PRPS1, PSMC4, PYGB, PYGL, QSOX1, RARS1, RBCK1, RPE, RRAGD, SAP30, SDC1, SDC2, SDC3, SDHC, SLC16A3, SLC25A10, SLC25A13, SLC35A3, SLC37A4, SOD1, SOX9, SPAG4, SRD5A3, STC1, STC2, STMN1, TALDO1, TFF3, TGFA, TGFBI, TKTL1, TPBG, TPI1, TPST1, TXN, UGP2, VCAN, VEGFA, VLDLR, XYLT2, ZNF292 |
| KEGG_ GLYCOLYSIS_ GLUCONEO GENESIS | ACSS1, ACSS2, ADH1A, ADH1B, ADH1C, ADH4, ADH5, ADH6, ADH7, AKR1A1, ALDH1A3, ALDH1B1, ALDH2, ALDH3A1, ALDH3A2, ALDH3B1, ALDH3B2, ALDH7A1, ALDH9A1, ALDOA, ALDOB, ALDOC, BPGM, DLAT, DLD, ENO1, ENO2, ENO3, FBP1, FBP2, G6PC1, G6PC2, GALM, GAPDH, GCK, GPI, HK1, HK2, HK3, LDHA, LDHAL6A, LDHAL6B, LDHB, LDHC, PCK1, PCK2, PDHA1, PDHA2, PDHB, PFKL, PFKM, PFKP, PGAM1, PGAM2, PGAM4, PGK1, PGK2, PGM1, PGM2, PKLR, PKM, TPI1 |
| KEGG_ OXIDATIVE_ PHOSPHOR YLATION | ATP12A, ATP4A, ATP4B, ATP5F1A, ATP5F1B, ATP5F1C, ATP5F1D, ATP5F1E, ATP5MC1, ATP5MC1P5, ATP5MC2, ATP5MC3, ATP5ME, ATP5MF, ATP5MG, ATP5PB, ATP5PD, ATP5PF, ATP5PO, ATP6AP1, ATP6V0A1, ATP6V0A2, ATP6V0A4, ATP6V0B, ATP6V0C, ATP6V0D1, ATP6V0D2, ATP6V0E1, ATP6V0E2, ATP6V1A, ATP6V1B1, ATP6V1B2, ATP6V1C1, ATP6V1C2, ATP6V1D, ATP6V1E1, ATP6V1E2, ATP6V1F, ATP6V1G1, ATP6V1G2, ATP6V1G3, ATP6V1H, COX10, COX11, COX15, COX17, COX4I1, COX4I2, COX5A, COX5B, COX6A1, COX6A2, COX6B1, COX6B2, COX6C, COX6CP3, COX7A1, COX7A2, COX7A2L, COX7B, COX7B2, COX7C, COX8A, COX8C, CYC1, LHPP, MT-ATP6, MT-ATP8, MT-CO1, MT-CO2, MT-CO3, MT-CYB, MT-ND1, MT-ND2, MT-ND3, MT-ND4, MT-ND4L, MT-ND5, MT-ND6, NDUFA1, NDUFA10, NDUFA11, NDUFA2, NDUFA3, NDUFA4, NDUFA4L2, NDUFA5, NDUFA6, NDUFA7, NDUFA8, NDUFA9, NDUFAB1, NDUFB1, NDUFB10, NDUFB2, NDUFB3, NDUFB4, NDUFB5, NDUFB6, NDUFB7, NDUFB8, NDUFB9, NDUFC1, NDUFC2, NDUFS1, NDUFS2, NDUFS3, NDUFS4, NDUFS5, NDUFS6, NDUFS7, NDUFS8, NDUFV1, NDUFV2, NDUFV3, PPA1, PPA2, SDHA, SDHB, SDHC, SDHD, TCIRG1, UQCR10, UQCR10P1, UQCR11, UQCRB, UQCRC1, UQCRC2, UQCRFS1, UQCRH, UQCRHL, UQCRQ |
| KEGG_ MTOR_ SIGNALING_ PATHWAY | AKT1, AKT2, AKT3, BRAF, CAB39, CAB39L, DDIT4, EIF4B, EIF4E, EIF4E1B, EIF4E2, EIF4EBP1, HIF1A, IGF1, INS, MAPK1, MAPK3, MLST8, MTOR, PDPK1, PGF, PIK3CA, PIK3CB, PIK3CD, PIK3CG, PIK3R1, PIK3R2, PIK3R3, PIK3R5, PRKAA1, PRKAA2, RHEB, RICTOR, RPS6, RPS6KA1, RPS6KA2, RPS6KA3, RPS6KA6, RPS6KB1, RPS6KB2, RPTOR, STK11, STRADA, TSC1, TSC2, ULK1, ULK2, ULK3, VEGFA, VEGFB, VEGFC, VEGFD |
| SUBSPACE_ LYMPHOID_ PROTECTIVE | ARL14EP, BPGM, BTN3A2, BUB3, CAMK4, CASP8, CCNB1IP1, CD247, CD3E, CD3G, DBT, DDX6, DYRK2, JAK1, KLRB1, MAP4K1, NCR3, PIK3R1, PLCG1, PPP2R5C, SEMA4F, SIDT1, SMAD4, SMYD2, TP53BP1, TRIB2, ZAP70, ZCCHC4, ZNF831 |

**Table S14. Gene sets used in Immune Dictionary cytokine signature analysis.**

| GENE SET | GENE SYMBOLS |
|---|---|
| IMMUNE_ DICTIONARY_ CD8_IL1B | SOCS3, BCL3, GADD45G, IGFBP4, GZMB, CRIP1, SOCS1, ARID5B, EMP3, SERPINB1, STAT3, SSH2, IFNGR1, GZMA, SBNO2, RGS10, ARID5A, FRMD4B, CYSLTR2, FAM241A, VARS, RGCC, NDRG3, HSD11B1, MXD1, DAPL1, CABLES1, ELOVL6, CARS, PLAC8, SKAP2, GBP2, C15orf48, FAM46A, ZBP1, AARS, MTHFD2, GPR171, CDKN2D, GPR18, NEB, RASGRP2, PIM1, FAM78A, FLOT1, THEMIS, ETV6, FAM102A, PPA1, PRELID2, ADGRE5, SMPDL3A, NSG2, ZNF281, BATF, BIN1, PPM1H, PJA1, ARL5C, ID3, CLIC4, RAMP1, CEBPB, KLHL6, TDRP, NARS, CD226, STAP1, SEMA4D, CRLF2, MAP3K8, SERPINB9, CCR9, MAPKAPK2, PPP1R16B, ICAM1, TREML2, CHST11, TSPAN32, RASGRP1, RRAS2, FKBP5, SLC41A1, CD96, SETBP1, USP6NL, GNGT2, RPAIN, S100A9, TRIB3, ITGAE, S100A8, LAMP2, YARS, IARS, LPP, RNF19A, MFSD6, DTX3L, C5orf30, SLC30A4, KIF1B, SEMA7A, ATP2B4, JAK3, SLC7A1, ACTN2, LARS, RCN3, RAB37, AHR, IDH2, SLC7A5, CLTB, MYD88, BCL2L11, PIM2, SPEF2, ABI3, SLC1A4, KIF23, CREM, TIMP2, ZFP1, LILRB4, SHMT2, GGT1, AKT2, SLPI, IRF8, ABCA2, CD55, CNDP2, TTC39B, RAB27A, TEX2, PDCD1, CKS2, MAP3K5, XPOT, GBP5, CPNE3, TRNP1, TESC, CXXC5, EPOP, MLXIP, EMC9, ANKRD28, RAPGEF3, RRP1B, GPR68, BTBD11, KCNA2, RRAGD, ALDH18A1, GPR146, PLEKHO1, HSPBAP1, LYPD6B, PLSCR1, MEX3A, CCR5, EIF4EBP1, GPRIN3, ACSS2, HSPA1A, HEXB, SLC15A2, TNIP2, AUTS2, BIRC3, MUC1, GADD45B, KCTD12, DBP, ABCB1, ODC1, GALNT10, ZC3H12D, MYO1F, TRPS1, RNF157, CEP164, KSR1, NTRK3, FGFR1, PSAT1, IL12RB2, CSRP1, SERPINI1, CCL17, FAM184A, IER3, FAS, EPHB2, AGFG2, SNX10, DGAT1, DLG3, JCHAIN, MUS81, BAHD1, RUNX2, IFITM10, ART4, FCGRT, SMPD5, HIST1H1D, METRNL, FRY, SLC15A1, CIART |
| IMMUNE_ DICTIONARY_ CD8_IL33 | CD79A, HSPA1A, BCL3, DHX58, FCMR, MS4A1, EBF1, SERPINB1 |
| IMMUNE_ DICTIONARY_ CD8_IL36A | PPA1, BCL3, ZBP1, ISG15, BST2, GBP2, EIF5A, GZMB, PSME2, ISG20, GADD45G, RNF213, EIF2S2, NARS, SHMT2, MTHFD2, NCL, RAN, GBP6, PHGDH, PA2G4, RTP4, SOCS3, ERH, RANBP1, CLIC4, CRIP1, STAT1, IGFBP4, CARS, XAF1, PSAT1, S100A10, BATF, CYCS, GBP6, EPRS, SOCS1, AARS, CCT3, UBA52, SRM, IFIT3, PARP14, IFI35, ST6GALNAC4, OAS3, HSP90AA1, VARS, IRF7, STAT3, NOP56, CEBPB, GADD45B, IFIT1B, GBP7, NOP58, LARS, HSPD1, PHB, EIF3C, LGALS3BP, CD82, SNRPD1, MAPKAPK2, TIMM8A, POLR2L, DDX39A, PCBP1, GBP6, RGS10, EMP3, KSR1, ODC1, C1QBP, GZMA, UCHL3, PARP9, TARS, MRTO4, LARP1, HDGF, MDN1, YARS, IMPDH2, STAT2, MYBBP1A, CHMP4B, IARS, SRSF7, ETV6, GARS, TAPBPL, DKC1, MARS, C15orf48, IRF8, EBNA1BP2, PRMT1, RRP1B, HSPA9, PSMA6, ZNF593, NMI, SARS, LAP3, NOLC1, TOMM40, IL7R, IFRD2, EIF4EBP1, RCC2, OAS1, LSP1, EIF1AX, SLC29A1, SLC7A1, TRAFD1, NAMPT, DHX58, RUVBL1, FAM162A, SNRPA1, WDR12, KLRD1, PRPF31, GBP5, NSUN2, IFITM3, ATAD3A, CALHM6, SLC7A5, NAA20, NOP16, KARS, GGT1, GBP6, MRPL17, ACP5, APEX1, OAS2, LMNA, FRMD4B, WARS, YPEL3, DDIT3, RUVBL2, ALDH18A1, MPP6, DCTPP1, PDCD4, SHMT1, CLTB, DTX3L, TRIB3, NOP2, TNFRSF9, EPOP, RRS1, AIMP2, IPO5, PPP1R16B, YBX3, XPOT, RASGRP2, GSTP1, NOP14, ARID5B, TIMM50, CD274, CKS2, AEN, NME2, ICAM1, USP18, PIM1, EIF2AK2, BIRC3, SLC19A1, IPO4, GALK1, C7orf50, HELZ2, HERC6, FAM136A, PUSL1, MTHFD1, TECPR1, KLHDC4, MUC1, FABP5, RRP9, LIPG, PIM2, KIAA0040, LAMP2, SKAP2, CXCL10, NFKBIA, POLR1A, DDX1, DAXX, NFKB2, AK6, SBNO2, CDK6, PLA2G12A, BYSL, PML, ZNF281, UMPS, TMA16, ITGB7, HIRIP3, GCSH, OR2V1, CD86, CLUH, RCC1, PITRM1, ANKRD44, PHF11, PABPC4, PLSCR3, IFIT3, STAT5A, MOV10, PYCARD, CASTOR1, LAD1, TNFRSF25, TMEM238, MECR, MTHFD1L, PISD, SLFN5, WDR4, POLE3, CAD, ADCY7, ADGRE5, TMEM97, RELB, DENND5A, IL18BP, SMPDL3A, PUS7, ZFP36L2, EEF1AKMT4, IFIH1, NOL10, SMYD2, THOP1, PARP12, PCGF6, HIF1A, MXD4, ADD3, IRGM, OASL, MACF1, CISH, NEFH, BCAT1, TFDP1, ARL5C, BIN1, ASNS, ACTN1, APP, RANGRF, GNG12, HSD11B1, IL2RA, PITPNC1, FEN1, TSC22D3, KIF21B, CDCA7, DOCK9, PNPT1, SLC30A4, HMGA1, GBP4, WDR36, RMI2, BNIP3L, CLPB, RMDN3, GBP6, TXNRD3, CTPS1, POP1, SESN2, FAM46A, POLR1B, TEX2, IFNG, PCK2, SLC25A33, ARHGEF18, SLC6A9, SLC1A4, DAP, JAKMIP1, ITGA4, TOP1MT, PHF10, HSH2D, SMIM13, MAP3K8, TSEN54, SLC7A6, CD96, DCTD, KLF3, PDF, SELENOP, SERPINB1, MS4A6A, TNIK, MID1IP1, POLR3H, FAM78A, CCR9, NSG2, PLEC, RAMP1, PLCG2, TDRP, USP6NL, TADA2A, HK2, HDAC7, SEMA7A, PREX1, PLAGL1, UBE2L6, CIART, FAM105A, TRIM37, ATP8A2, SIPA1, AHNAK, FURIN, SLC12A7, MARCKSL1, CD55, OSM, MXD1, LY6K, HMGN3, C19orf38, TLR7, KPNA2, TWF2, TMEM108, POLD4, ACTN2, UTF1, CHAC1, ATF6, CYSLTR2, RASGRP1, RAPGEF4, ACAA2, AHR, AEBP1, TNNT2, RNF157, RAVER1, BCL6, GM2A, STIM1, TPST1, TSPAN32, HVCN1, MGST2, KIT, EVA1B, CMTR1, IL12RB2, SLC25A15, CXCR4, SETX, RCN3, IL12RB1, PSMC3IP, CREM, SORCS2, USP31, EPB41L4B, TCP11L2, RETREG1, AHCY, TM6SF1, SYNE1, COQ4, HADH, ITGA6, CD40LG, ADD1, TNF, TTC28, MDK, SPATS2, C11orf98, FAAH, TNKS1BP1, BCL9L, MRPL53, HOOK2, LCN2, TIMM23, MEX3A, IFI44, FRMD8, STX11, TIMP2, PDCD1LG2, C17orf80, GOLM1, GSDME, ATP2A3, DHX37, HEXB, CSPG5, DGAT1, SGSM3, CCRL2, RAB37, ARRB2, S100A9, MMACHC, RASA3, SYCE2, MTA3, GPT2, CABLES1, CHST11, ACSS2, FAM185A, EPHX1, GRIN2C, MLKL, RACGAP1, MYO5A, PDZK1IP1, PCMTD2, PIPOX, ITGAE, PER2, NR6A1, FRY, MAP4K2, ZMIZ1, MICAL1, SUV39H2, LETM2, ACSS1, NLRC3, PINK1, IDH2, MTBP, MYO10, TANC1, AAED1, PHF11, KIF23, SLC28A2, TLCD1, OXCT1, NTRK3, AKAP1, CDC14B, LIF, BBC3, EMC9, BAG2, IGKC, ZNF428, FCGRT, GSTT2, MFSD2A, TNFAIP8L1, BSPRY, IFITM2, ST3GAL6, TMEM69, MYO1F, KLHL11, CENPA, NUTF2, LYPD6B, CXXC5, COX4I2, RFX3, KCNA2, IZUMO4, AK7, FKBP11, SGSH, RNF144A, STK35, IKZF4, AGRN, RHOB, COL9A3, PRNP, ABCA2, METTL27, MTURN, RFX1, CDC42EP3, TBXA2R, GAB3 |

| GENE SET | GENE SYMBOLS |
|---|---|
| IMMUNE_ DICTIONARY_ CD8_IL2 | GZMB, PPA1, EIF5A, NME1, NCL, SRM, PPP1R14B, PHGDH, RAN, RANBP1, CDK6, HSP90AA1, SHMT2, CYCS, EIF4A1, REXO2, YBX3, MIF, ANP32B, SOCS1, NPM3, PA2G4, SLC29A1, SLC25A5, TUBA1B, LDHA, MTHFD2, CDCA7, SET, CCND2, HSPD1, ATP5G1, MYBBP1A, SHMT1, ODC1, YBX1, DDX21, NOLC1, TUBA4A, TUBB4B, LARP1, IRF8, KLF2, C1QBP, HSPA8, DDX39A, GALK1, CCT3, SRSF2, ENO1, NOP56, MTHFD1, NHP2, MRTO4, NSUN2, PSAT1, PABPC4, TIMM8A, HSPA5, ST6GALNAC4, MAT2A, CISH, CAD, IARS, RPS27L, FABP5, EPRS, IPO5, EIF3A, EBNA1BP2, NOP58, MDN1, HSPA9, EIF2S2, SNRPD1, FBL, HNRNPAB, DCTPP1, IL7R, BCL3, NOC2L, SNU13, SRSF7, TOMM5, NOP16, G3BP1, MCM2, KARS, NARS, CACYBP, APEX1, ERH, GNL3, CANX, PCBP1, MPP6, DKC1, LTA, KPNB1, EIF3C, EIF3B, FKBP2, APRT, TMEM238, HSPA4, MRPS28, BANF1, IL2RB, YARS, EIF4G1, BZW1, ZNF593, SLC19A1, RRP15, AARS, CLUH, PRMT1, TOMM40, RRP1B, UQCC2, TSPAN4, RCC2, ACOT7, VARS, TPI1, LARS, RSL1D1, BYSL, UNG, TXN, LAP3, NAP1L1, SLC7A1, HDGF, WARS, CORO2A, RMI2, RGS10, NOP2, TIMM10, PUS7, LYAR, IPO4, TARS, RRS1, FASN, RPN1, SYNCRIP, PSME3, MRPL20, FAM162A, TIMM9, STAT3, BOP1, PHB, PPRC1, UCHL3, ATAD3A, RRP9, NDUFAB1, UTP20, PPAN, DTX1, ABCE1, EEF1E1, GCSH, PGK1, CLIC4, TSC22D3, PGAM1, UCK2, GAR1, HSP90B1, YPEL3, THOP1, ETF1, GART, IFRD2, MAGOHB, SLAMF7, PPID, METTL1, NEFH, EIF1AY, SAR1A, SMYD5, PUM3, JUN, RUVBL2, AIMP2, BZW2, BTG2, NIP7, SNRPA1, EIF2S1, FTSJ3, RUVBL1, CHEK1, MARS, MRPL12, CDK4, GBP6, CARS, RARS, BSPRY, NUDC, GADD45G, DNAJC2, MRPL17, XPOT, SFXN1, PDIA6, SMYD2, WDR12, SDF2L1, STIP1, CDV3, PTGES3, MYDGF, MTHFD1L, POLR1B, IL2RA, LILRB4, TMEM97, MTAP, STAT1, EPOP, SETBP1, LARP4, EIF4E, MANF, RPF2, HEATR1, RANGRF, IFNG, MCM5, CCDC86, ATIC, TCERG1, CDH1, AGPAT5, GNL1, NIFK, CNDP2, RRP12, POLR2H, MCM3, WDR77, PRPF31, CTPS1, PFKP, SSSCA1, LIF, PNO1, TUBA1C, PPAT, JAML, NOMO1, TMA16, GPATCH4, PIK3IP1, SEPT11, EIF1AX, HSPBP1, YWHAG, GARS, PUSL1, CD3EAP, TFDP1, DCTD, TBX21, RCL1, CINP, RCC1, UBL4A, USP36, AKT1, SLC35A4, ERAP1, NOL10, GBP2, FIGNL1, EIF2B3, GEMIN5, WDR46, PREP, PRPF19, EFHD2, ATP2A2, NOP14, PFAS, SMC4, CDC34, NAT10, SMARCC1, PCNA, TOMM70, TEX2, AGPAT3, NUP62, MFSD2A, AEN, KLF6, WDR18, S1PR1, DIS3, PITRM1, GBP7, TNK2, CHSY1, DAPL1, XBP1, AGFG1, CHCHD4, CAMKK2, SNRPA, NASP, POLR1A, ASNS, LRPPRC, POLR3D, GRWD1, UMPS, CAPG, YRDC, TSR1, UTP18, HYOU1, ACSL5, HELLS, COMTD1, LGALS3BP, PELP1, IRGM, KMT5A, LTV1, ID3, NOC4L, LRRC59, POLR2L, ZBP1, TIPIN, SMCO4, SYPL1, F2R, NEK6, TRMT61A, SYCE2, WDR3, P4HB, HK2, ZNHIT6, TFRC, MCM6, FEN1, FAM46A, SLC16A1, PPIF, DNAJC11, PLA2G12A, FAM136A, CRELD2, HIRIP3, CD160, DYNLL2, WEE1, DDX1, SZRD1, NLE1, SMPDL3A, IRF1, PDXK, NAMPT, ALDH18A1, KLRC1, ZMYND19, PRAG1, HBEGF, ARMC6, ACTN1, CDC25A, C7orf50, SERPINB9, FLT3LG, WDR4, WDR5, YDJC, NUP205, SLC35B1, WDR36, NUP93, BCAT1, TBL3, CCR5, CA12, NRARP, EIF4EBP1, UBQLN4, FOS, UHRF1, SLC7A5, DHX33, SCO2, SAMD1, ARL5C, EEF1AKMT4, ALG8, PWP1, SEMA7A, CD7, DTL, MEMO1, SPOUT1, TAF5L, PCGF6, CDCA7L, PWP2, CKS1B, PUS7L, CCDC58, NAF1, ERGIC1, RCN1, BHLHE40, SLAMF6, NME2, SOCS3, NAA25, BNIP3L, ISG20L2, GINS2, SLC25A33, ANKRD13B, PIM3, GSTO1, BAK1, ELAC2, RUNDC3B, PACSIN2, CCDC85C, UXS1, MAPKAPK2, PIM2, ACACA, RNF157, SLC25A22, CRLS1, SLC25A15, MTRR, NOL6, KAT2A, AMZ1, TRIM37, NCOA5, PCSK4, MOV10, IL12RB1, NOC3L, CHAF1A, S100A8, CD320, PTPRS, KIF21B, B4GALT5, FKBP5, FASLG, NOL9, MXD4, SPATA5, TMEM71, MAPK6, RCC1L, S100A9, SLC1A4, FCER1G, OAS3, KLHL24, LEO1, RIPK3, FAM185A, FXN, PRELID2, LRRC41, RBM19, MEX3A, CDKN1B, E2F6, FOXK2, TDRP, PPP1R15A, TMEM163, NSD2, AKAP1, MECR, CD274, CIART, PDCD1LG2, SLC35C2, RB1CC1, CARM1, ABI3, AEBP1, POLD4, LPCAT4, JAKMIP1, JPT2, NUDCD1, CXCR4, LMNB2, TDG, MID1, TYROBP, URB2, POP1, AMPD2, BSN, POLR3H, KLRB1, SENP3, IL15RA, GBP6, HIST1H1C, MGST2, GFM1, TTLL4, HMGA1, TADA2A, FPGS, NDC1, STYK1, TXNRD3, KPNA2, PHF10, PER2, PPP2R1B, XCL1, RMDN3, MAFK, HGH1, SUPV3L1, CCL2, CCDC102A, ABCC1, DAP, TP53RK, NUP188, NT5DC2, MAFF, TOP1MT, RAPGEF4, TNFSF10, ALPL, CENPS, TGM2, WDR74, CCR9, SHQ1, FAM129B, CASTOR1, AQP9, TSEN54, TNFRSF21, ATP23, ENOPH1, USP31, ABCB1, PRDX4, MCRIP2, BAG2, SLC29A2, TSPAN32, NUTF2, CYP51A1, SELENOP, CD55, LCN2, FKBP11, PRIM1, RCN3, GZMA, TMEM158, BBC3, TGIF2, F2RL2, SMPD4, TTLL12, EVA1B, JUNB, PCMTD1, EDARADD, LETM2, FNIP2, EXO1, NBN, TRDC, BEND3, HSPA1A, SUV39H2, CRYBG2, SOCS2, OSM, CD244, U2AF1L4, SCARB1, RHOQ, IZUMO4, ANKS1A, TXNRD2, SPIRE1, TPST1, IFT80, PLCXD2, TLE6, MTFP1, ACY1, SPRED1, NDST1, CCDC137, E2F1, DPAGT1, DNAJC27, PLK3, ADRM1, EMC9, MIER2, SMG8, NCR1, TNNT1, SLC25A13, TLCD1, IL12RB2, IL1R2, MS4A6A, STK39, DHRS7, TIMM23, NETO2, NR6A1, TNFSF11, GPRIN3, TNFRSF9, CENPA, ZNF367, DNA2, KCNQ5, CCL22, EPCAM, DSCC1, HIC1, FRAT2, KLHDC1, SGO1, PINK1, TBKBP1, AAED1, KLRB1, PTRH1, SLC5A6, PLP2, IFITM3, PTGER3, HBP1, FRMD4B, GBP4, ACSS2, ZNF566, LYPD6B, FAM46C, ART4, KCNA2, DHX37, RSAD1, GINS1, PLCG2, SRRD, TMEM150A, MCM10, TYW3, EEA1, LITAF, RACGAP1, PALB2, L2HGDH, LRP8, HEXB, MYO19, NDOR1, SPATA6, GCAT, LPL |
| IMMUNE_ DICTIONARY_ CD8_IL4 | HLA-DMA, DUSP10, IL4R, BCL2, SOCS1, PPA1, PHGDH, SHMT2, EIF5A, CDK6, CD8A, ENO1, MTHFD2, NOP56, EIF2S2, NCL, SRM, PLAC8, S100A10, NME1, GLIPR2, C1QBP, NCOA7, PPP3CC, AK2, NARS, PA2G4, CDCA7, CYCS, MBD2, FABP5, PPP1R14B, TUBA4A, METTL22, MARS, RANBP1, METTL1, CNP, CRIP1, TSPAN9, LYST, CARS, TCOF1, GNL3, EMP3, ERH, KLRD1, APEX1, NOP58, OTULIN, GALK1, CXCR6, LARS, TIMM8A, MRTO4, EPRS, DKC1, DDX21, PRPF31, PHB, AARS, KLF2, MYBBP1A, PRELID2, RMI2, EIF4EBP1, MRPS28, SLC7A1, GZMA, NCOA3, TMIE, GARS, PSAT1, NOLC1, SLC29A1, YARS, RRP15, SLC39A6, UCHL3, WDR43, RGS1, TIMM10, ST6GALNAC4, FOXN3, GAR1, GMFG, NOC2L, DDIT3, LAP3, PLA2G12A, EBNA1BP2, TOMM40, HS6ST1, RUVBL1, VIM, CD3EAP, UBL3, MTHFD1, WDR46, EEF1E1, IFRD2, DCTPP1, ATAD3A, ALDH18A1, NSUN2, RRS1, NIFK, NOP2, ZNF593, C12orf73, BYSL, MYC, VARS, TIMM9, NOP14, XBP1, UNG, NEFH, SMYD2, IARS, EOMES, UTP18, GRWD1, RRP9, PPAT, TPI1, RUVBL2, ALS2CL, IL7R, CDCA7L, SHMT1, ADK, RPF2, TMA16, GCSH, RNF19B, SCO2, RANGRF, GGACT, TARS, TRIB3, ZBP1, CLIC4, LSP1, WDR12, WDR74, PRPS1, CINP, S1PR1, HMBS, NOP16, SFT2D2, ZNHIT6, CTPS1, PUS7, DCTD, CISH, MCM2, CA12, NUP43, SUCLG2, SLC19A1, POLR3D, TRMT61A, ACTN1, ITGB7, EPOP, COMTD1, KIAA0040, PINX1, YDJC, |

| GENE SET | GENE SYMBOLS |
| --- | --- |
| | SPOUT1, SLC1A4, TRMT11, SLC16A2, UCK2, KLF6, RASGRP2, AHNAK, PUSL1, HIST1H4A, BTG2, RCN1, XPOT, GEMIN6, PROS1, XAF1, ZFP36L2, TECPR1, ACP5, TNP2, PCSK4, TMEM97, CAD, POLR3H, NME2, SMYD5, NFIL3, DYRK4, WARS, IPO4, YBX3, FAM46A, HLADMB, BIN1, F2R, TFDP1, ARHGAP26, DGKA, SLC7A5, PDXK, ISG20, SPECC1, PITRM1, TRAT1, YPEL3, TMTC4, PLXNA1, POLR1B, GEMIN5, THOP1, ETFBKMT, JUN, CDH1, CD7, CLUH, S100A11, TSC22D3, SCRN2, PECR, SLC25A33, AR, FOS, HGH1, MCRIP2, NEK1, TSPAN4, ABHD14A, CHAC1, SYCE2, OSBPL6, ANGPTL4, GADD45G, HSPA1A, IPCEF1, IL18R1, NEURL3, AQP9, ASNS, P2RX7, WEE1, KPNA2, TOP1MT, CIART, ARMC6, HK2, EFNA5, MTRR, FANCF, TADA2A, PDE7A, C7orf50, FRMD4B, SPN, TXNRD3, ARL5C, VCPKMT, ADGRE5, WDR59, C16orf74, PIK3IP1, SH2D1A, SLAMF6, GRAMD1A, BAG2, ARHGEF18, BSN, DDC, TRAF1, FRMD6, MXD4, MALT1, TP53INP1, SMPDL3A, KIF21B, SH3BP2, TMEM71, MADD, SLC12A7, CHEK1, LIPT2, TIMM23, ZNF566, RRAGD, FIGNL1, STIM1, NUTF2, CD55, RSAD1, TDRP, FOXO1, CDH13, MTFP1, AARSD1, THRB, SESN2, DUSP1, CXCR4, SLC6A9, POU2F2, SUV39H2, PYCR1, ESYT2, NETO2, S100A1, SELENOP, ZBTB20, GPT2, RGS2, SLC25A15, MYO19, PLEC, U2AF1L4, TLCD1, TUBA1A, CCL5, ACSS2, IKZF2, MYO1F, ST3GAL1, UBE2L6, DTX1, NEB, C19orf57, LRRC32, RNF144A, GATA1, CDKN2D, ALPL, BCL9L, JUNB, LRP8, SGK1, SNRNP35, MMACHC, GGT1, TPST1, FRMD8, MAP2, RAPGEF4, DCLRE1A, PGLYRP1, TGIF2, DOCK11, PRODH, ANXA2, RETREG1, GOLM1, JAKMIP1, DAP, F2RL2, DDIT4, ACY1, IGKC, SGIP1, RACGAP1, DNAJB1, ATP1B1, CD9, FAM69B, FRY, USP2, ZNF428, TRMT5, TECPR2, COL15A1, UPRT, HEXB, EHD3, RABGAP1L, OAS1, PRF1, ENG |
| IMMUNE_ DICTIONARY_ CD8_IL6 | IFIT3, ISG20, RSAD2, IFIT3, WDR90, PRIM1, MX1 |
| IMMUNE_ DICTIONARY_ CD8_IL7 | BCL2, EIF5A, REXO2, PPA1, PHGDH, RAN, BST2, PPP1R14B, ENO1, NME1, CDK6, SOCS1, NCL, SRM, ISG15, PA2G4, SHMT2, RANBP1, LTA, MTHFD2, EIF2S2, HSPD1, NOP56, C1QBP, CCT3, IL7R, JAML, GNL3, IFI35, NARS, TUBB4B, NOP58, MRTO4, SNRPD1, TUBA4A, CYCS, RTP4, EBNA1BP2, DKC1, TCF7, TIMM8A, METTL1, CARS, PIM1, DDX39A, CHCHD1, IGFBP4, BYSL, EIF2S1, KLRD1, PSAT1, RNF213, RRP9, SLC7A5, PLAC8, CISH, MYC, TIMM10, MARS, APEX1, GAR1, NOLC1, FKBP4, STIP1, SLC29A1, RGS10, IFRD2, MYBBP1A, HSPA9, SLC7A1, PHB, IARS, LAP3, PRPF31, RRP15, AIMP2, UCHL3, RRP1B, EPRS, ISG20, AARS, SMC4, NOC2L, ZBP1, EIF4EBP1, NSUN2, GALK1, RUVBL1, EEF1E1, RRS1, ATAD3A, DCTPP1, GARS, CORO2A, CNDP2, CDCA7, PPAN, TIMM9, CCDC86, NDUFAF4, XAF1, GADD45G, IRF7, UNG, ST6GALNAC4, GZMB, NOP16, NIFK, ZNF593, GCSH, WDR12, MAP3K8, BTG2, ODC1, LGALS3BP, MRPS18B, AGPAT3, ATP8A2, UTP18, CD3EAP, HERPUD1, YARS, TSC22D3, YPEL3, FTSJ3, OSM, GRWD1, NOP2, RPF2, SSSCA1, EIF1AX, IFIT1B, IPO5, NAF1, UCK2, ALDH18A1, MOV10, RMI2, PUSL1, LTV1, CLIC4, SOCS3, SIGMAR1, LARS, PNO1, TIMM50, SHMT1, TPI1, SLC19A1, RUVBL2, PUS7, RCL1, IFIT3, FLT3LG, IPO4, MTHFD1, COMTD1, TARS, AEN, UMPS, SMYD5, TMEM238, CD7, CTPS1, FASN, AATF, SETBP1, SCO2, ZNHIT6, DAPL1, NEFH, ARL5C, XPOT, PIK3IP1, USP18, PPAT, CHCHD4, POLR3D, TMEM158, NOC4L, ACTN1, YDJC, PRMT7, TMA16, TRMT61A, HSPA1A, FABP5, SMYD2, PIM2, CLUH, DCTD, TFAP4, TMEM97, MXD4, CD28, WARS, NEURL3, DDIT3, CXCR4, PRELID2, TRIB3, POLD2, RFLNB, TNF, FBXO17, ZDHHC8, MCM2, RRP12, PLA2G12A, USP50, ADAM19, IL2RA, CA12, HIRIP3, CIART, BCL3, ACOT7, WDR36, NOL10, SPATA5, RCC1, POLR1B, SLC30A4, TDRP, JUN, ARMC6, PIM3, CCR9, EPOP, DAP, DYRK4, PISD, C15orf48, TEX2, SLC1A4, MCM5, POLR3H, DHX58, HK2, TSPAN4, AMPD2, SOCS2, TMEM71, NSUN5, ZMYND19, FEN1, EEF1AKMT4, TP53INP1, FOS, IKZF4, SLC12A7, MRM3, GPR174, IFIT3, WEE1, THOP1, SCML4, IKBKE, CASTOR1, LIF, USP31, OAS2, SENP3, FAM46C, OAS1, SYNE1, PCSK4, CD55, ITGAE, ACSS2, TXNRD3, PCMTD2, LDLR, ASNS, LIPT2, SYCE2, CD79A, IFT80, LYPD6B, POP1, C7orf50, TADA2A, GOLM1, GSDME, PHF11, ACTN2, LMNB2, AMZ1, ZER1, GPT2, SLC25A15, RGS2, MTFP1, RAPGEF4, SNTB2, CHAC1, TTC28, SUV39H2, ST3GAL1, DZIP1, OASL, HERC3, RHOB, CHEK1, FBXL20, MYO1F, DTL, FIGNL1, CARD6, HBEGF, SLC26A11, TRIM5, SLC25A40, EMC9, RSAD2, CEP164, SGSH, MFSD2A, SQLE, HLA-DOA, NETO2, HSF4, TLE1, RCN3, CDC14B, SNX8, ABCA1, PCK2, CALCRL, SESN2, SLC28A2, PYCR1, TLR7, UBE3D, UBE2L6, EPHX1, PER2, TOB1, TFDP2, HID1, TLCD1, HSD17B11, BTBD11, RRAGD, MYO19, THRA, MYB, KCNA2, PHLDA3, NUTF2, ART4 |
| IMMUNE_ DICTIONARY_ CD8_IL12 | STAT1, SOCS1, IRF1, GBP6, GBP7, GBP2, PPA1, GADD45G, ZBP1, RTP4, PIM2, IRF8, ST6GALNAC4, IRGM, GBP4, SOCS3, BCL3, AHR, GZMB, CD274, GBP5, GBP6, PRNP, IFNG, FURIN, POP1, IL2RA, HSPA1A, C3orf33, SETBP1, PDP1, CXCR4, TNKS1BP1, PTGER3, FOS, HDAC6 |
| IMMUNE_ DICTIONARY_ CD8_IL27 | ZBP1, GBP2, PLAC8, IRF7, ISG15, IFIT3, XAF1, TSC22D3, FOS, BCL3, IL12RB1, GBP4, OAS1, CYSLTR2, IFIT3, GZMA, GADD45G, GBP6, IL18BP, GZMB, ITGAX, OAS2, RGS2 |
| IMMUNE_ DICTIONARY_ CD8_OSM | LARS2, IGFBP4, DTX1, SOCS3, TRNP1, BCL3, NEB, NME2, HSPA1A, ISG20, FOS, RSAD2, IFIT1B, SKAP2, USP18, JUN, KLF6, IFIT3, IFIT3, GADD45G, SLFN5, DHX58, CHST11, CMPK2, SERPINB1, RHOB, SPATS2, PPP1R15A, PRELID2, PHF11, IGSF23, DYNLT1, DUSP1 |
| IMMUNE_ DICTIONARY_ CD8_IL10 | BCL3, CEBPD |

| GENE SET | GENE SYMBOLS |
|---|---|
| IMMUNE_ DICTIONARY_ CD8_IL22 | FOS, HBB |
| IMMUNE_ DICTIONARY_ CD8_IFNB | ISG15, IFIT3, IFIT1B, ISG20, SLFN5, ZBP1, USP18, PHF11, RNF213, IRF7, BST2, RTP4, IFI16, RSAD2, XAF1, HERC6, GBP7, PPA1, IFI35, PARP14, GBP2, OAS3, DAXX, SAMHD1, JAML, EIF2AK2, IFI16, IFI16, DDX60, IFIT3, MX1, PLAC8, IFI16, TOR3A, TRAFD1, IFI16, PSMB10, OASL, DHX58, IFIT2, IFITM3, CMPK2, PSMB8, IFIH1, LY6E, HLA-E, STAT1, PARP9, GBP6, LGALS3BP, CCND2, SLFN13, GBP6, PHF11, TAPBP, IFIT1, SMCHD1, CXCL10, PML, SOCS1, DDX58, SAMD9L, PHF11, PSME1, OAS2, EPSTI1, CD86, CLIC4, IFI16, HELZ2, NMI, C19orf12, GADD45G, PSME2, OAS1, SELENOW, GZMA, CHMP4B, ATP8A2, TSPO, DTX3L, SP110, PARP12, THY1, STAT2, IFI16, NCOA7, CTSS, CASP8, PSMB9, ZUFSP, CRIP1, SLCO3A1, NAMPT, ASB13, GBP6, LGALS9, GBP5, UBA7, NAA20, USP25, GBP6, UBE2L6, CCRL2, HSPA5, EMP3, TREML2, TAP1, MOV10, TMBIM6, KLF2, TRIM25, S100A10, SDC3, ASCC3, IFI44, ZNFX1, ETNK1, CALHM6, KLRD1, PARP10, PARP11, MITD1, VARS, TOR1AIP1, TRIM5, CD274, CNP, TCF7, TLR7, GIMAP1-GIMAP5, MAX, RNF114, TBRG1, DDX24, DBNL, IRGM, ADAR, CYCS, EOMES, PLA2G16, ST6GALNAC4, MS4A6A, TMEM184B, GBP4, B4GALT5, TOR1AIP2, HSH2D, PCGF5, NLRC5, S1PR1, PHGDH, RASGRP2, PHIP, ZFP36L2, AIDA, TCOF1, HLA-A, ZCCHC2, ZNF365, CTSD, PHF11, TRIM6-TRIM34, SP140, PHC2, OGFRL1, DESI2, TDRD7, KEAP1, TNFSF10, RBL1, ARF4, TRIM21, IL18BP, GZMB, ITPR1, OGFR, GPR18, FAM111A, YPEL3, TAPBPL, LUZP1, TECPR1, TRIM56, TNP2, ACP5, PGD, IGFBP4, ADGRE5, MTHFD2, IFNGR1, IL12RB1, ETV6, LIPG, ACTN1, DUSP28, SLC25A22, CYSLTR2, PIM1, MORC3, AHNAK, SETDB2, FNBP4, RFC3, ATP10A, KIF21B, IRF9, PSME2, ARL5C, NEURL3, UBALD2, GBP6, FLNA, PNPT1, TMEM140, MVB12A, PLSCR3, CARS, RFLNB, ZYX, MXD4, RMI2, BCL3, HDAC7, MYD88, TTC39B, CD5, SLFN12L, SOCS3, PAPD7, MLKL, PDLIM2, CD7, TMEM192, GNG2, SLAMF7, C9orf3, AP1S3, STIM1, JAKMIP1, POU2F2, RBM43, MAP2K1, TLR3, SEPT9, FOS, IRF8, LGALS8, GMPPB, C6orf106, SNX2, C19orf66, TMEM106A, RABEPK, N4BP1, HK1, CISH, GCH1, PMEPA1, GRINA, SLFN13, RAMP1, GM2A, SLC7A1, GOLM1, SHMT2, CD200R1L, CXCR4, AARS, SLC7A5, ISOC1, CEBPB, XDH, HVCN1, ANKFY1, LAMP2, LASP1, IFITM10, EVI2A, MAFK, IARS, VPS54, ZMIZ1, DDIT3, P2RY14, BBC3, MYO1G, CXCR3, ATM, PPP1R15A, PGLYRP1, MAP7, TRIM14, NLRC3, PLCXD2, SGK1, IFI16, RETREG1, EHD4, CITED4, YARS, SIT1, APOBEC1, SSBP3, SORL1, SEMA7A, ANXA5, CPNE3, LIF, MAP3K8, TBXA2R, RCN3, PPM1K, PHLPP1, PEAK1, OSM, UBALD1, COX18, SNTB2, SH3BP2, ACTN2, WARS, PRKCE, STOML1, ARHGAP26, CCNYL1, STK26, OR2V1, VWA5A, ABCA2, CITED2, INSL6, CD72, SGSH, CD55, CCDC107, RACGAP1, UBASH3B, MYO1F, SLC17A9, RGS2, FURIN, SYNE1, TRIM5, CEP164, DHRS7, NBEAL2, CXXC5, HADH, IL10RA, TTC28, CDC42EP3, TPCN1, G0S2, SERPINI1, CARD19, GRN, ELK3, RHOB, SKI, N4BP2, TRIB2, SLC12A6, ILDR1, IZUMO4, BTBD11, MTM1, PLEKHG2, PPCDC, PINK1, SLC26A11, GPSM2, MIER3, GPRIN3, CENPA, MAST3, ITGA6, SYTL2, PIGA, CMTR1, ACSS2, TNS1, RASA4, SFXN3, ITGAE, NMRAL1, DUSP1, CD44, ZFC3H1, ANXA2, GALNT10, MAN2A2, CSTA, CGAS, PRELID2, TMIE, PGM2L1, FCGRT, SESN1, MAPK3, GNG12, SH3GLB2, MILR1, MYB, LITAF, LPIN1, ADAMTS6, CD9, TOB1, PCMTD2, CDC14B, FBXL20, AAED1, CDC25B, ZER1, NUDT14, PKD1, TUBA8, MTURN, CASP4, MAPRE2, CEP97, QRFP, FRY, HID1, PEA15, RNF157, IFNG, RANBP10, LMO2, CXorf21, RNF130, HS3ST3B1, SMAD3, IL15RA, ZFHX2, AGRN, PLEKHA5, ITM2A, GOLIM4, SAMD1, ARHGAP5, PAQR7, TAGAP, SLC25A40, ARMC3, NRM, RTN4RL1, SKAP2, MCEE, CTDSP2, LATS2, DTWD1, RASSF3, IL3RA, ETHE1, TRIB3, ABTB1, PLIN3, ARRB1, HDAC5, LAG3, GSN, STK39, TESK1, PACS2, P2RX4, METTL27, MOSPD3, MXRA8, SLC2A3, IZUMO1R, SPECC1, DHRS1, SLC43A2, VIPR1, SLC28A2, CHKA, FAM168A, TMEM141, IFNGR2, TRAT1, FRAT2, CHIC1, NR4A2, LRRC75B, TNFAIP8L1, KCNJ8, PXYLP1, STRADA, SPICE1, ARHGAP18, HIST4H4, MAP3K3, HIST1H1D, HAUS4, SFMBT2, ALS2, WBP1, FAP, SHISA2, GAS7, CALCRL, ELOVL7, AGFG2, COL23A1, NIPAL3, ACCS, MNS1, CHAC1, UNC5CL, DDC, CASTOR1, SOX4 |
| IMMUNE_ DICTIONARY_ CD8_IFNG | GBP2, GBP6, STAT1, GBP7, ZBP1, IRF1, SAMHD1, RTP4, ISG15, SOCS1, GBP6, BST2, CD274, IFIT3, GBP6, IRGM, TAPBPL, GBP6, RNF213, PARP14, GBP5, IRF7, PARP9, DTX3L, PPA1, IL18BP, PLA2G16, XAF1, GBP4, IRF8, ISG20, IFIT1B, GBP6, NAMPT, OAS3, WARS, USP18, IFIT3, INSL6, TRAFD1, ST6GALNAC4, PARP10, IFI44, STAT2, IFIH1, OAS1, FOS, UBE2L6, PLSCR3, BCL3, NME2, CXCL10, CMPK2, MTUS2 |
| IMMUNE_ DICTIONARY_ CD8_TNFA | IGFBP4, BCL3, CRIP1, NFKBIA, VARS, KLF2, RELB, PPA1, NFKB2, DDIT4, ZBP1, SOCS3, MAPKAPK2, GBP2, GADD45G, CARS, SDHAF1, JUN, ISG15, BIRC3, FKBP5, KLF6, TRNP1, BTG2, GZMB, BATF, AARS, SHMT2, XAF1, HSPA1A, ODC1, FOS, MTHFD2, NFKBIE, IARS, BBC3, MARS, ARID5B, CLIC4, ETV6, SKAP2, ST6GALNAC4, ICAM1, TNFRSF9, YARS, ISG20, ADGRE5, GBP5, FAM46A, MXD4, NEB, INSL6, PRELID2, PLSCR3, C15orf48, PLCG2, CYSLTR2, TRIB3, KSR1, EIF4EBP1, PPP1R15A, SLC7A5, GZMA, RHOB, SERPINB1, RGS2, CD55, CHST11, SLC1A4, SPATS2, SHMT1, CABLES1, MID1IP1, EPOP, ACTN2, SLC28A2, S100A6, CDC25B, CD83, CCL5, IFITM10, EMC9, C3orf33, IL12RB1, FOSB, TOB1, PDCD1LG2, NR4A2, VIPR1, OAS2, MYO10, CIART, ASNS, IL18BP, NOX1, EPHX1, ABCB6, SYT6 |
| IMMUNE_ DICTIONARY_ CD8_TL1A | NFKBIA, RELB, NFKB2, ICAM1, CD82, BCL3, IKBKE, CD83, NFKBIE, BIRC3, TNFRSF9, TNFRSF25, UTF1, MGLL, FOS, LAD1, HSPA1A, GADD45G, ULBP1, PDCD1LG2, LIPT1, EFCAB2, DENND5A, SORCS2, DUSP1, ADAMTSL2, ZBTB46, CMTR2 |
